# Genome-Wide Association Study for Glucocorticoid-Induced Ocular Hypertension

**DOI:** 10.64898/2025.12.31.25343232

**Authors:** Jyoti Lama, Renee Liu, Alicia Huerta-Chagoya, Ashley Li, Katie Huynh, Lynn K. Stanwyck, Samuel Han, Yan Zhao, Weilin Chan, Liyin Chen, Ananya Mukundan, Da Meng, Janine Y. Yang, Gayatri Susarla, Delia Sang, George N. Papaliodis, Lucy Q. Shen, Elizabeth Rossin, Christine Elkins, Irma Benavides, Hussein Wafapoor, Antonio Cutino, Janey L. Wiggs, Alexander M. Eaton, Ayellet V. Segrè, Lucia Sobrin

## Abstract

**Purpose:** To identify genetic variants associated with glucocorticoid (GC)-induced intraocular pressure (IOP) change using genome-wide association study (GWAS) and whole exome sequencing (WES) analyses.

**Methods:** 530 participants from the Fluocinolone Acetonide in Diabetic Macular Edema (FAME) trials were analyzed, with replication performed in an independent cohort of 588 participants from the Mass Eye and Ear/Retina Health Center (MEE/RHC). All participants were exposed to GC, primarily via intravitreal injection. IOP was measured at baseline and serially within 6 months following GC exposure. GWAS and rare variant gene burden analyses were applied, adjusting for covariates.

**Results:** Genetic associations for maximal IOP change within 6 months after GC exposure were evaluated. For the primary outcome across all ancestries in FAME, one variant, rs13425173 within the *UBE2E3* locus reached genome-wide significance (P=2.88 X 10^-8^). In the FAME and MEE/RHC meta-analysis, variant rs1040227, also in the *UBE2E3* locus, was significantly associated at the genome-wide level (P=2.88 X 10^-8^) and showed nominal significance in the MEE/RHC cohort (P=0.02). In the colocalization analyses, the significant FAME GWAS *UBE2E3* locus was linked to expression regulation of this gene in six tissues including artery aorta. In gene-level analysis, *UBE2E3* also demonstrated subthreshold significance (P=6 X 10^-6^). 532 FAME and 586 MEE/RHC participants were included in the WES gene burden analysis. One gene*, MSTO1,* passed false discovery rate correction for the primary outcome in FAME.

**Conclusion:** We have identified genome-wide significant common variants associated with GC-IOP change, as well as genes and rare variants that may influence GC-induced IOP change.

## Introduction

Glucocorticoids (GCs) have an anti-inflammatory effect and are used systemically and locally to treat many ocular disorders. Intraocular pressure (IOP) elevation occurs in a significant proportion of patients treated with GC and can lead to glaucomatous optic nerve damage and vision loss.^1^ Of the delivery modes, intraocular injection of GC is associated with IOP elevation more frequently and severely.^2^ Approximately one third of patients have an elevated IOP of 10 mmHg or more after intraocular GC injection.^3^ Risk factors for IOP elevation include personal or family history of primary open-angle glaucoma (POAG), with up to 90% of POAG patients exhibiting increased IOP after GC exposure.^4–8^ Twin studies demonstrated a heritability index of 0.35 for GC-induced ocular hypertension (OHTN), consistent with polygenic inheritance.^9^ Studies also suggest that patients with close relatives with POAG have a higher risk of GC-induced OHTN compared to those with no primary relatives with POAG.^10–13^

While several studies suggest GC-induced OHTN is heritable, few studies have investigated the genetic factors contributing to GC-induced OHTN risk and there have been no validated genetic susceptibility variants identified.^14–18^ Suggestive evidence of an association between mucin gene *HCG22* and GC-induced IOP elevation was found in a genome-wide association study (GWAS) of 113 patients of primarily European (EUR) ancestry but has not been independently replicated.^17^ The genetic association between *MYOC* and GC-induced OHTN has been studied, with carriers of *MYOC* mutations having more aggressive glaucoma and earlier IOP elevation than POAG patients who do not carry the mutation.^15^ Other genes have been studied, including *NR3C1*, but are limited by small sample sizes that are unable to detect modest effects expected from common variants.^14^ In addition, there are no studies that have examined rare genetic variants. The purpose of this study is to perform genome-wide genotyping of two cohorts of patients treated with GC, primarily through intravitreal GC injection, and investigate common and rare variants associated with GC-induced OHTN.

## Methods

### Participants

Institutional Review Board /Ethics Committee approval was obtained for all cohorts. All patients provided written consent.

### Discovery Cohort

Participant samples for the discovery cohort were derived from Fluocinolone Acetonide in Diabetic Macular Edema (FAME) trials which examined the efficacy of intravitreal inserts releasing 0.2 µg/day or 0.5 µg/day fluocinolone acetonide in patients with diabetic macular edema (DME).^19^ Complete details of the study protocol including inclusion and exclusion criteria and examination methods have been described previously.^20,21^ Briefly, participants had their IOPs measured by Goldmann applanation tonometry in a standardized fashion at baseline, 1 week, 6 weeks, 3 months and every 3 months thereafter. FAME participants were given the option to consent to provide a blood sample for genetic investigations.

### Replication Cohort

Participant samples for the replication cohort were from two clinical sites – the Mass Eye and Ear (MEE) and Retina Health Center (RHC). Participants were identified by review of the medical record. Inclusion criteria included exposure to GC. The GC exposure was intravitreal in the majority of participants, but participants could also be enrolled if they received GC via periocular injection, drops or oral formulations. Demographic and clinical information was extracted from the medical record, including IOPs at baseline, 4-6 weeks, 3 months and 6 months after initial GC exposure. IOPs were measured by a variety of methods but primarily Goldmann applanation and Tono-Pen Handheld tonometry. For a given participant, priority was given to recording IOPs that were measured using the same method across clinical visits.

Exclusion criteria were age < 18 years old, active uveitis during the 6-month IOP monitoring period, history of glaucoma surgery, intraocular surgery within 3 months of GC exposure, hypotony at baseline (IOP < 8 mmHg), endophthalmitis, and ocular trauma.

### Outcome Definitions

The quantitative trait of IOP change was the primary outcome of this study. IOP change was defined as the difference between IOP within a month before GC exposure and the maximum IOP measurement in the first six months after treatment. In the case that the patient had both eyes treated with GC, only the eye with the largest IOP change was analyzed.

Two secondary dichotomous outcomes were also tested: responder vs. non-responder and extreme responder vs. extreme non-responder. A GC responder was an individual with a normal IOP at baseline (8-22 mmHg) and an IOP elevation of ≥ 6 mmHg after GC exposure. A non-responder was defined as an individual with a normal baseline IOP and an IOP that did not increase or increased < 6 mmHg after GC treatment. Extreme GC responders were individuals with a normal baseline IOP and an IOP increase of ≥ 10 mmHg after GC treatment. An extreme non-responder had a normal IOP at baseline and an IOP that did not increase or increased < 3 mmHg after GC treatment. In both the secondary dichotomous outcomes, the treatment period was defined as 6 months following GC exposure.

Since the discovery cohort was only exposed to GC through intravitreal injection, while the replication cohort also contained patients exposed to GC through other means, we performed a sensitivity analysis of the primary outcome using only the subset of replication cohort participants who received intravitreal GC. The rationale for the sensitivity analysis was to harmonize the two cohorts with respect to GC delivery method to increase the power to detect associated variants.

### Covariates Corrected for in GWAS and Gene Burden Testing

The covariates corrected for in the discovery cohort GWAS for common variants included demographic variables [age, sex, top 5 genotype principal components (PCs)], clinical features (GC medication delivery route, GC medication dose, history of glaucoma and/or ocular hypertension, and use of IOP- reducing eye drops during the 6 months after GC exposure).

The covariates corrected for in the rare variant gene burden tests also included the above demographic and clinical variables with an addition of the number of synonymous mutations in whole exome regions per individual. The top 10 genotype PCs were used to correct for population stratification in analyses involving the multi-ancestry (ALL) samples population and top 20 genotype PCs for analyses examining only the European (EUR) population, to ensure correction for subtle population substructures that could have confounding effects in the rare variant analyses. To evaluate population differences that needed to be taken into account in the analyses, we also examined the distribution of maximal IOP change between ancestries (please see Supplemental Methods available at https://www.aaojournal.org).

The covariates corrected for in the replication cohorts are the same as described for discovery cohort except additional clinical features (disease indication for GC and diabetes status) that were only applicable to replication cohort. The covariates corrected for in the gene burden tests are the same as described for discovery cohort except the top 10 genotype PCs were used to correct for population stratification for all analyses.

### Genotyping and Sequencing

Deoxyribonucleic acid (DNA) was extracted from whole blood and was genotyped on the Global Screening Array (GSA) at the Genomics Platform at the Broad Institute (Cambridge, MA).

Whole exome sequencing (WES) was performed on the Illumina platform with a mean sample coverage rate of 96% and a mean of 96.41% of target bases covered at 20X. We performed best practices sample and variant quality control (QC) procedures (please see the Supplemental Methods, Supplemental Tables 1-5 and Supplemental Figure 1). The QCed array genotype calls were imputed to the TOPMed r3 (hg38) reference panel (please see Supplemental Methods). Data generated by this study can be accessed on dbGAP, study ID phs004037.

### GWAS Analysis

Association testing was performed on genotyped and imputed dosage scores following imputation using PLINK 2.0 for both the cohorts on all three outcomes (Primary outcome: Maximum IOP Change, Secondary Outcome: Responders vs Non-responders, Secondary Outcome: Extreme Responders vs. Extreme Non-responders) as listed in Supplemental Table 6. GWAS analyses were performed across all populations [(FAME): EUR, African (AFR), Admixed American (AMR), South Asian (SAS) and (MEE/RHC): EUR, AFR, AMR] and separately only on the EUR population, which had the largest number of participants. Linear or logistic fixed effects regression models were applied to the quantitative or dichotomous outcomes, respectively (using ‘--glm’ from PLINK 2.0), adjusting for the covariates listed above. Sensitivity analysis was performed for primary outcomes using a method similar to mixed effects modelling, REGENIE (version 3.2.6; https://rgcgithub.github.io/regenie/)^22^ that accounts for polygenic effects, population structure, and cryptic relatedness, using a two-step ridge regression framework. Rank-Based Inverse Normal Transformation (RINT) was applied to the quantitative phenotype, Maximum IOP Change, due to its non-normal distribution, to reduce false positives driven by outlier values in GWAS analysis. GWAS was performed on both RINT and non-normalized values to obtain raw beta estimates from the latter, which allows for better biological interpretation on original unit scale (IOP change in mmHg). To identify independent signals per locus we applied linkage disequilibrium (LD) clumping (‘-clump’ from PLINK 2.0) for all nominally significant single nucleotide polymorphisms (SNPs) passing P<10^-5^ using the corresponding study sample set for calculating LD. The genomic inflation factor, lambda, was calculated for each GWAS analysis to assess whether the results were inflated or deflated.

Quantile-quantile (Q-Q) plots and Manhattan plots were generated using the qqman package in R. LocusZoom plots were generated using the LocusZoom tool (https://my.locuszoom.org/#).^23,24^

### Replication in MEE/RHC cohort and GWAS Meta-Analysis of FAME and MEE/RHC

To identify the replicated variants, we tested whether any of the top (P<10^-5^) GWAS loci from the discovery cohort (FAME) were present in the replication cohort (MEE/RHC) and passed replication Bonferroni cutoff (P<0.05/no. of tested loci in FAME) or a nominal significance (P < 0.05). Among these we further focused only on loci that passed the replication Bonferroni cutoff or a subthreshold significance (P<10^-5^) in the meta-analysis of all cohorts and had the same direction of effect in both the cohorts. The replication analysis and the meta-analyses were performed for all three outcomes (Maximum IOP Change, Responders vs. Non-responders, and Extreme Responders vs. Extreme Non-responders) across ALL populations and the EUR population. We used METAL^25^ to perform the fixed-effects inverse-variance weighting meta-analysis.

### Fine-Mapping

We performed statistical fine-mapping of the GWAS summary statistics of genome-wide significant loci using SuSiE^26,27^ from the susieR package (v0.14.2). Mapping was applied for 6,001 variants within the 2Mb region within the genome-wide significant locus from the discovery GWAS and Meta-analysis. We allowed for presence of a maximum of 10 causal signals or 95% credible sets per locus and a cumulative sum of posterior inclusion probabilities (PIP) >0.95 per credible set. LD matrix was derived from the genotype data of the same study population used in association analysis for discovery GWAS and 1000G for Meta-analysis. LD matrix was computed using cor() function in R.

### Colocalization Analysis of Expression and Splicing Quantitative Trait Loci (QTLs) with Associated Genomic Loci

To identify the candidate causal genes and underlying regulatory mechanisms that may be driving the genetic associations with GC-induced OHTN in the significant GWAS loci, we conducted colocalization analysis between the GWAS loci and expression and splicing quantitative trait loci (eQTLs and sQTLs) from GTEx tissues^29^ and eQTLs from retina^28^ (using eCAVIAR.^30^ Colocalization posterior probability above 0.01 was considered significant. Details of the colocalization methods are in the Supplemental Methods.

### Gene and Gene-set Level Association Analysis of GWAS using MAGMA

To identify genes and pathways or gene ontologies associated with GC-induced OHTN, we conducted gene-based and gene-set (pathway) enrichment analyses of genetic associations using MAGMA.^31^ Methodological details of these analyses are in the Supplemental Methods.

### Gene and Pathway Rare variant Burden Test using Whole Exome Sequencing

To identify genes and pathways with a higher frequency of predicted deleterious rare variants in cases compared to controls and vice versa, we performed gene- and gene set-based burden tests for rare variants with different levels of predicted functional effects (Supplemental Table 7). The test was conducted for both discovery and replication cohorts, and finally meta-analysis was performed between the two cohorts. Details about the different levels of predicted functional effects and additional variant QC steps on the final WES analysis sample set are in the Supplemental Methods, Supplemental Figure 2, and Supplemental Table 8. A negative control of nonsynonymous are variants was performed. The replication analysis for rare variants was performed as described above for common variants. Bonferroni correction and the Benjamini-Hochberg false discovery rate (FDR) method were used to correct for multiple hypothesis testing in the gene and gene-set burden tests.

## Results

### GWAS Participant Characteristics

After QC steps were applied, 530 participants from discovery (FAME) cohort and 588 participants from the replication (MEE/RHC) cohort were included in the GWAS analyses (Figure 1). The demographic and clinical characteristics of the GWAS participants are shown in Table 1. The distribution of the maximal IOP change within 6 months of GC treatment is shown in Supplemental Figure 3. The mean maximal change in IOP within the first 6 months after GC exposure for the discovery and replication cohorts were +5.76 mmHg and +5.9 mmHg, respectively (Table 1). The distribution of maximal IOP change by ancestry is shown in Supplemental Figure 4. We did not observe a significant difference between ancestries in the discovery cohort [ANOVA, degrees of freedom (df) =3, P = 0.99], but we observed a significant difference in the distribution of maximal IOP change by ancestry in the replication cohorts (Analysis of variance [ANOVA], df=2, P=0.01), in particular between EUR and AFR ancestries (mean difference= -2.9, Tukey’s honest significant difference, P=0.0099). We corrected for ancestry in the GWAS analyses using the top genotype PCs as described in the Supplemental Materials and Supplemental Figures 5 and 6.

**Figure 1.**
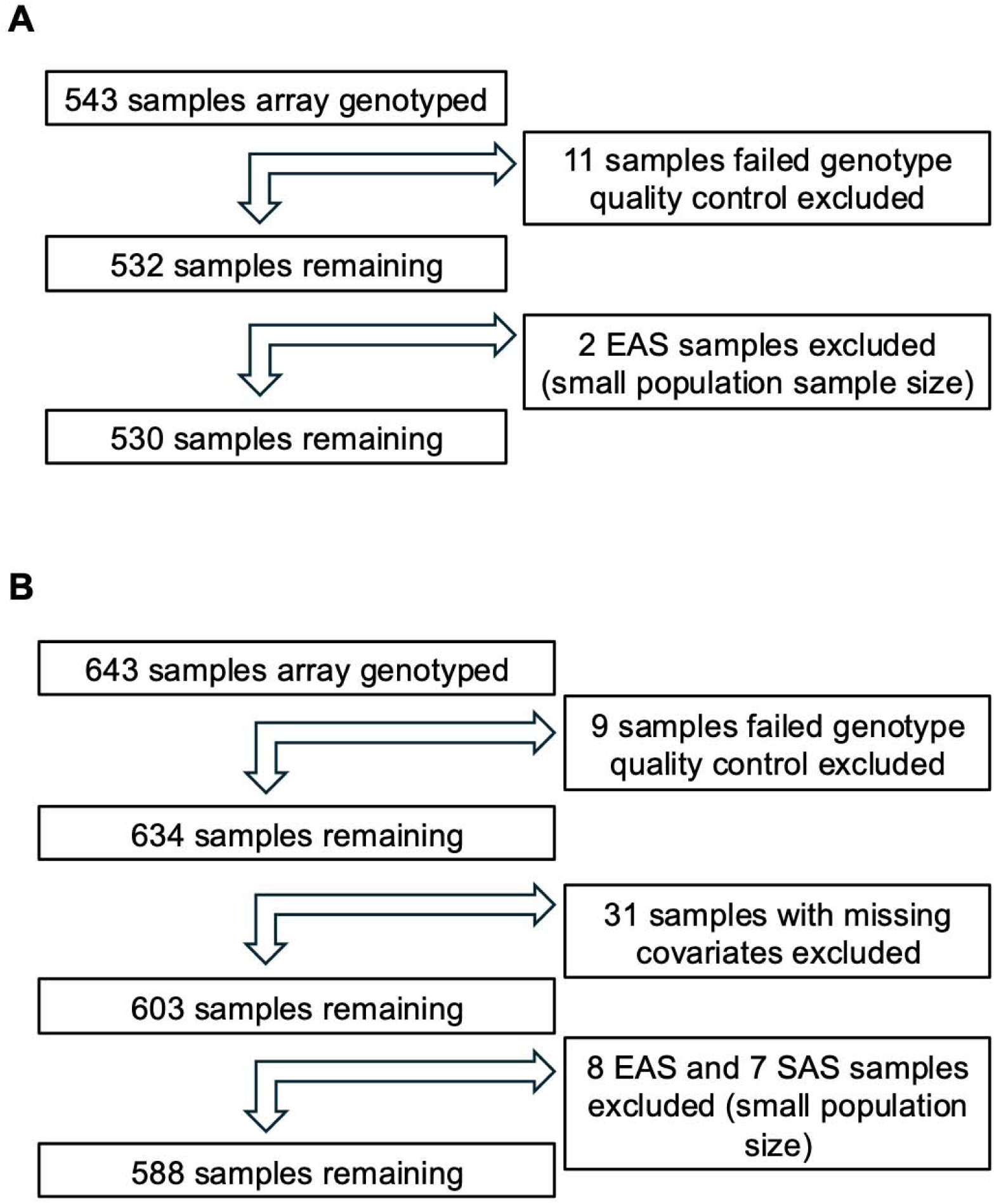
Flow chart of sample exclusion and total number of samples used in the genome-wide association studies (GWAS) for the **(A)** Fluocinolone Acetonide in Diabetic Macular Edema (FAME) samples, and **(B)** Mass Eye and Ear (MEE)/Retina Health Center (RHC) samples. EAS, East Asian; SAS, South Asian.

**Table 1.**
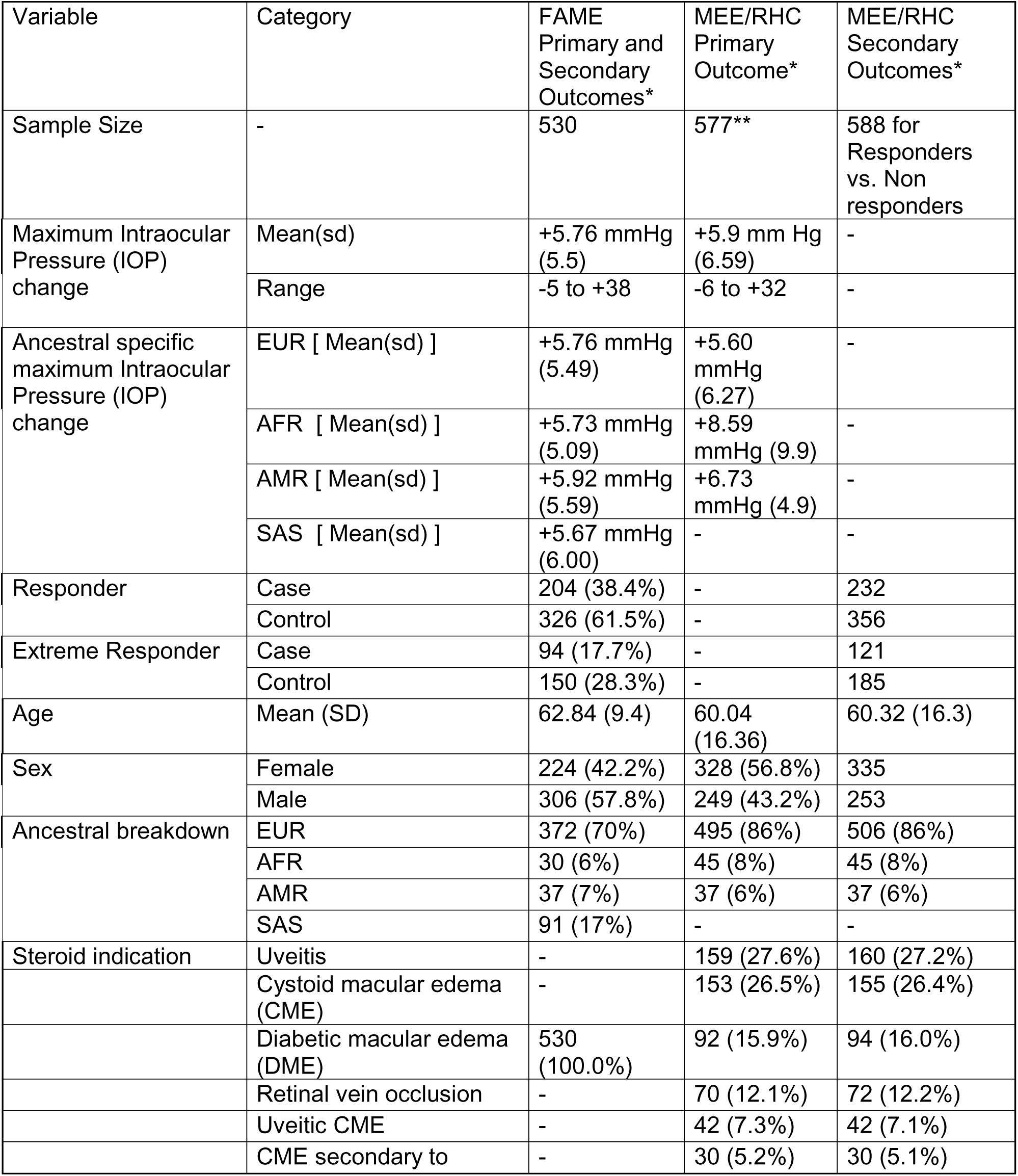
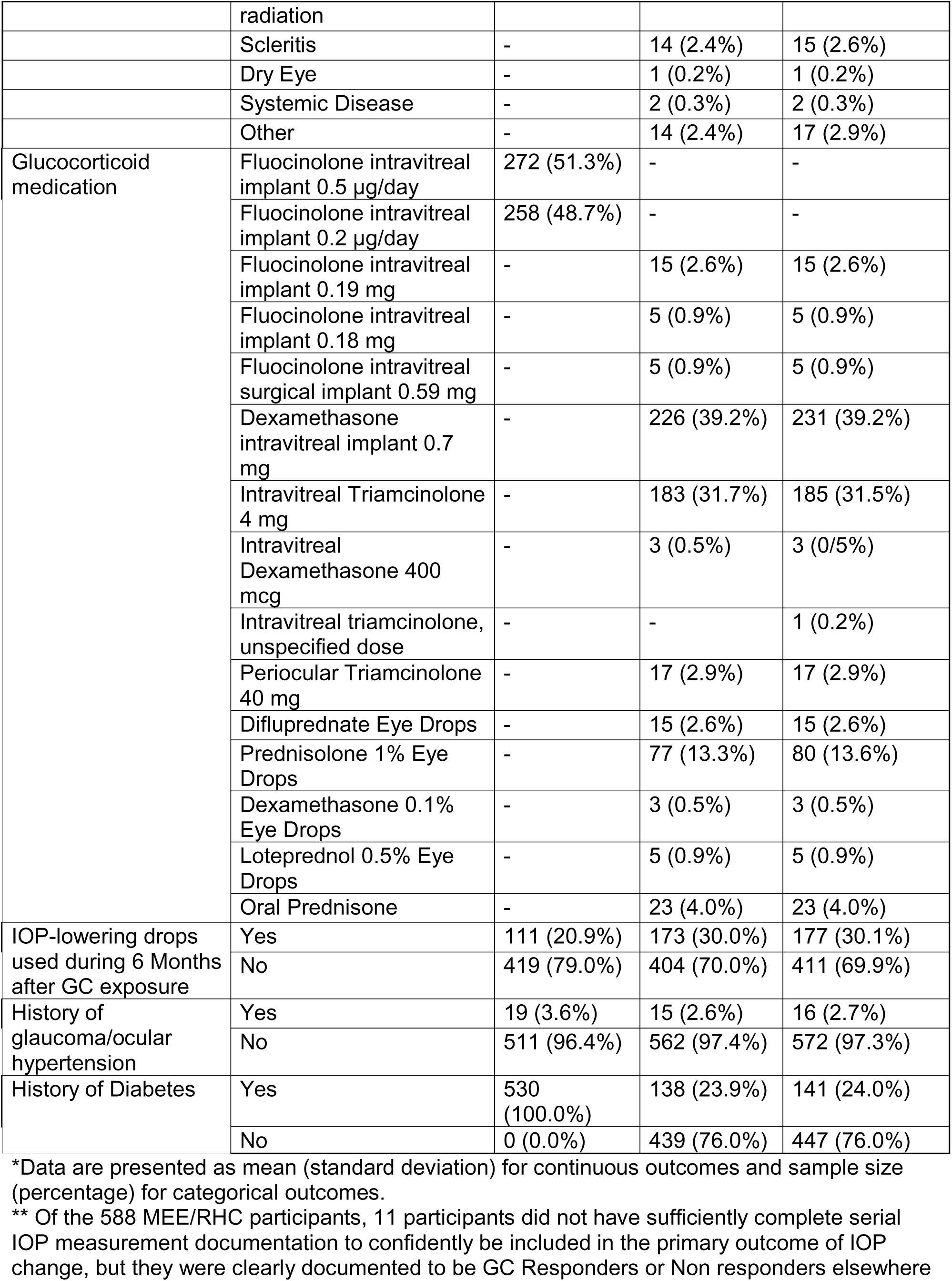
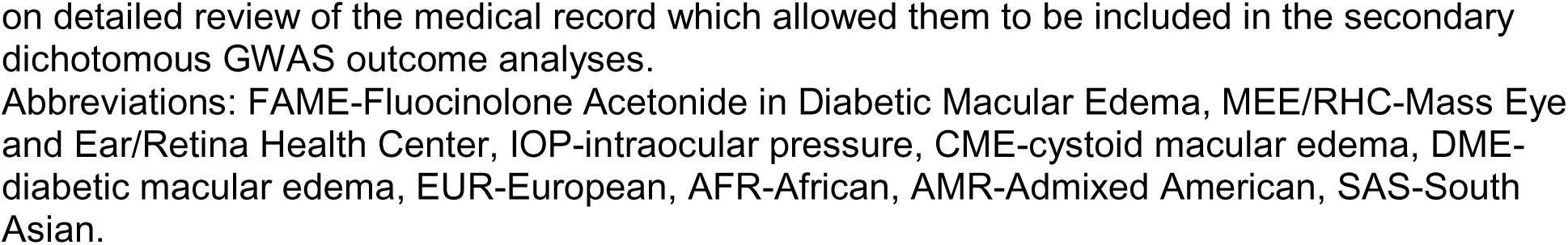
Phenotypic characteristics of the discovery cohort, Fluocinolone Acetonide in Diabetic Macular Edema (FAME) trial, and replication cohort, Mass Eye and Ear (MEE)/Retina Health Center (RHC), included in the genome-wide association studies (GWAS).

The number of genome-wide significant (P < 5x10^-8^) or suggestive (P < 10^-5^) LD-independent loci per discovery or replication GWAS and the meta-analysis GWAS are shown in Supplemental Table 6.

### Common variant GWAS

In the GWAS of common variants one variant, chr2:181004816:G:C (rs13425173), reached genome-wide significance (P = 2.88 X 10^-8^) in the FAME discovery cohort using the primary outcome across ALL ancestry (Figure 2, Table 2). GWAS using the secondary outcomes or only the EUR subset did not identify any genome-wide significant associations in the discovery cohort (Supplemental Figures 7 and 8). Next, we investigated replication in the MEE/RHC cohorts for this variant. Variant chr2:181004816:G:C had the same direction of effect in the MEE/RHC replication cohort but did not meet the threshold for significant association (p= 0.09) and was shy of being genome-wide significant in the FAME-MEE/RHC meta-analysis (P= 1.2 X 10^-7^). However, in the FAME-MEE/RHC meta-analysis, two SNPs in the *UBE2E3* locus [chr2:181021297:G:A (rs1040227) and chr2:181019696:A:C (rs7355651)], and both in high LD (r^2^=0.85 and 0.82) with chr2:181004816:G:C, did reach genome-wide significance (P = 1.23 X 10^-8^ and 1.3 X 10^-8^) (Table 2, Figure 3). These two variants passed subthreshold significance in the meta-analysis (P=2.05 X 10^-7^ and 2.02 X 10^-7^) using REGENIE, a whole-genome regression method that approximates a linear mixed model and better accounts for ancestry-based differences (Supplemental Figures 9-10, Supplemental Table 9).

**Figure 2.**
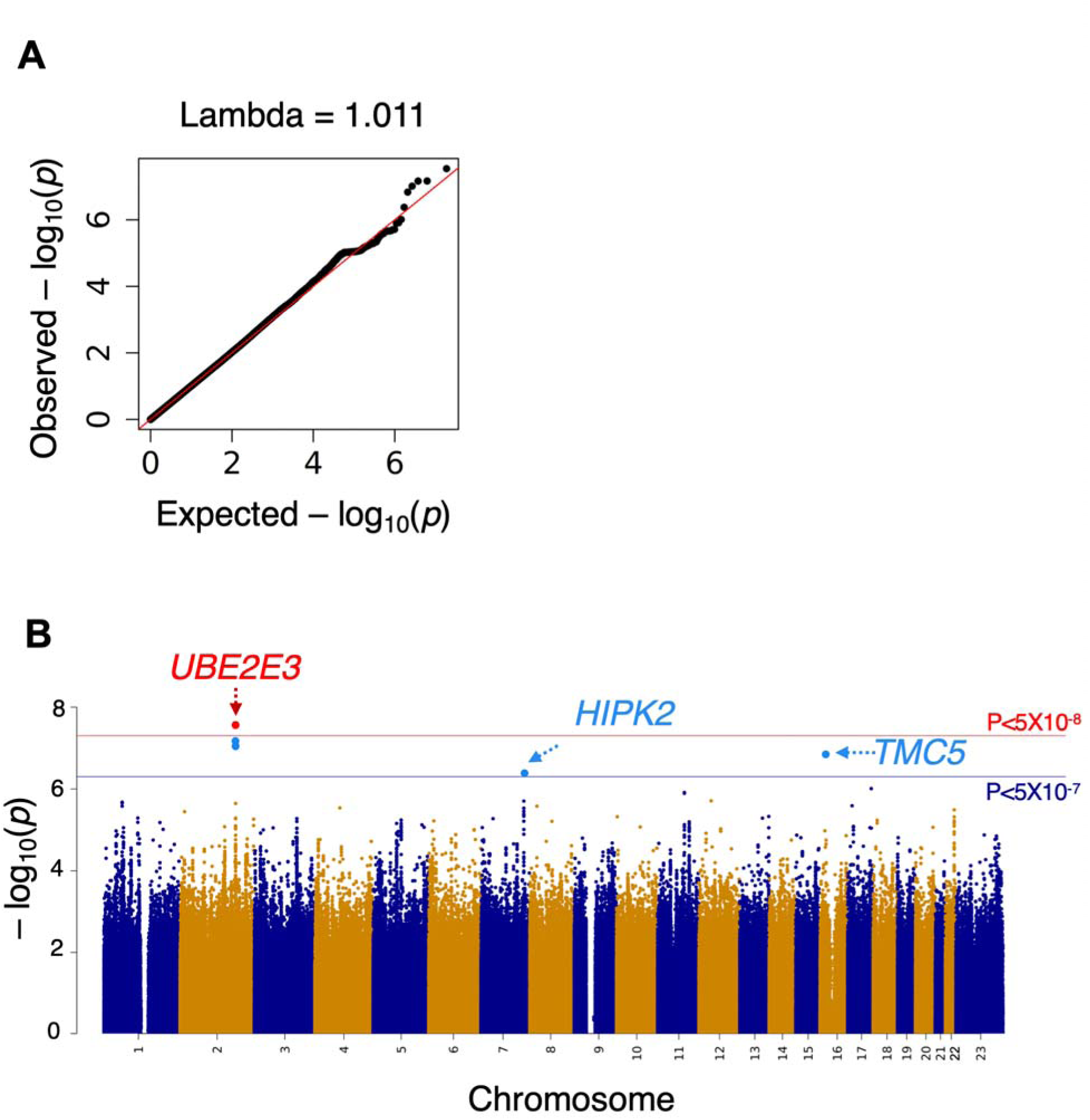
Quantile-quantile (Q-Q) plot (A) and Manhattan plot (B) of maximal intraocular pressure (IOP) change within 6 months in the ALL samples (n=530) genome-wide association study (GWAS) discovery cohort, Fluocinolone Acetonide in Diabetic Macular Edema (FAME) trial. The lambda of 1.011 suggests that there was no significant inflation.

**Figure 3.**
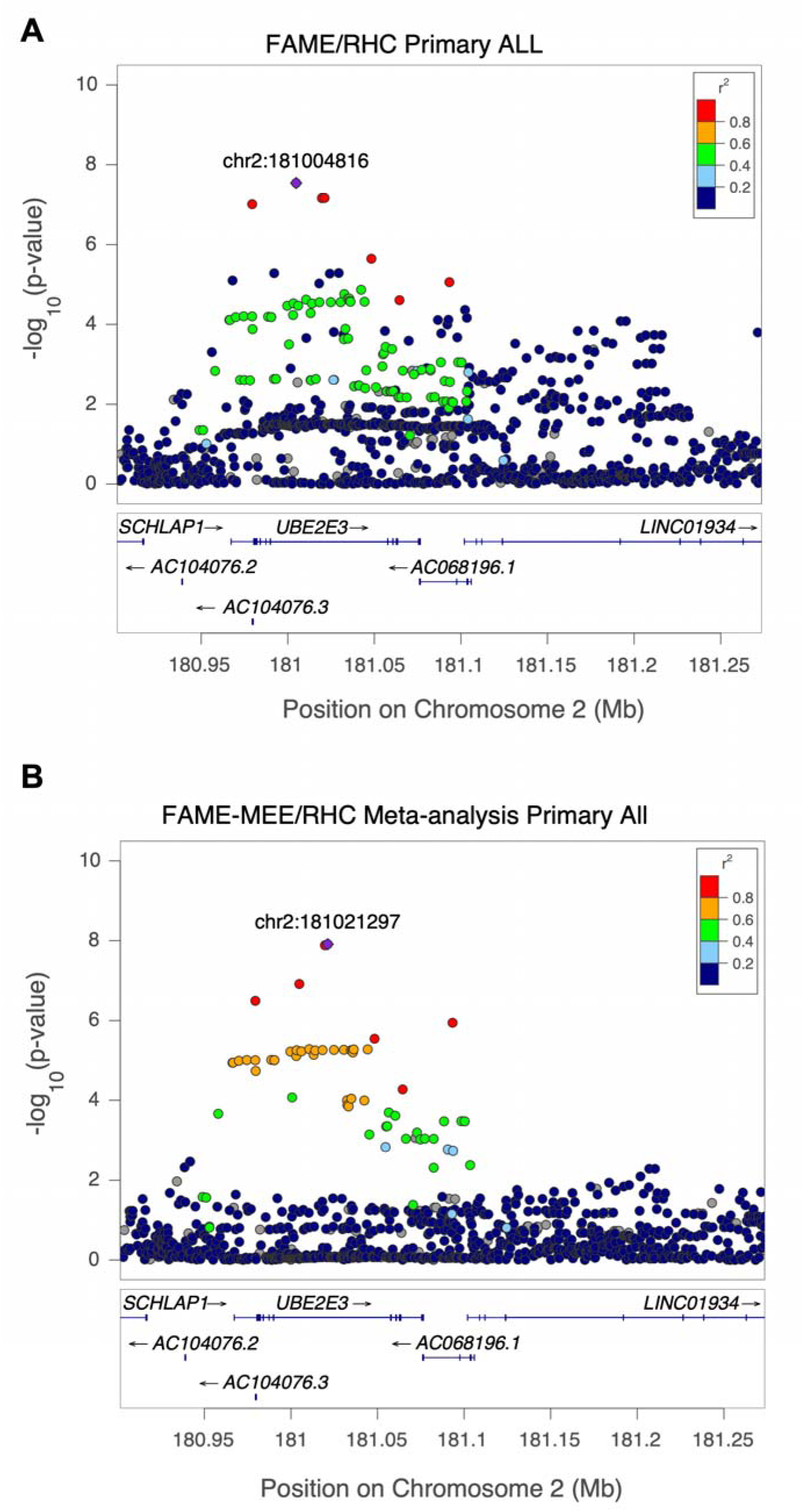
LocusZoom plot for the genome-wide significant locus for maximal intraocular pressure (IOP) change for **(A)** ALL ancestries in Fluocinolone Acetonide Diabetic Macular Edema (FAME) trial (rs13425173, chr2:181004816:G:C) and **(B)** the ALL ancestries in the FAME - Mass Eye and Ear (MEE)/Retina Health Center (RHC) meta-analysis (rs1040227, chr2:181021297:G:A). The y-axis is the GWAS -log_10_ P value and x-axis is the chromosomal position in genome build hg38. Variants are color-coded by the extent of linkage disequilibrium (r^2^) relative to the lead GWAS variant represented by the purple diamond.

**Table 2:**
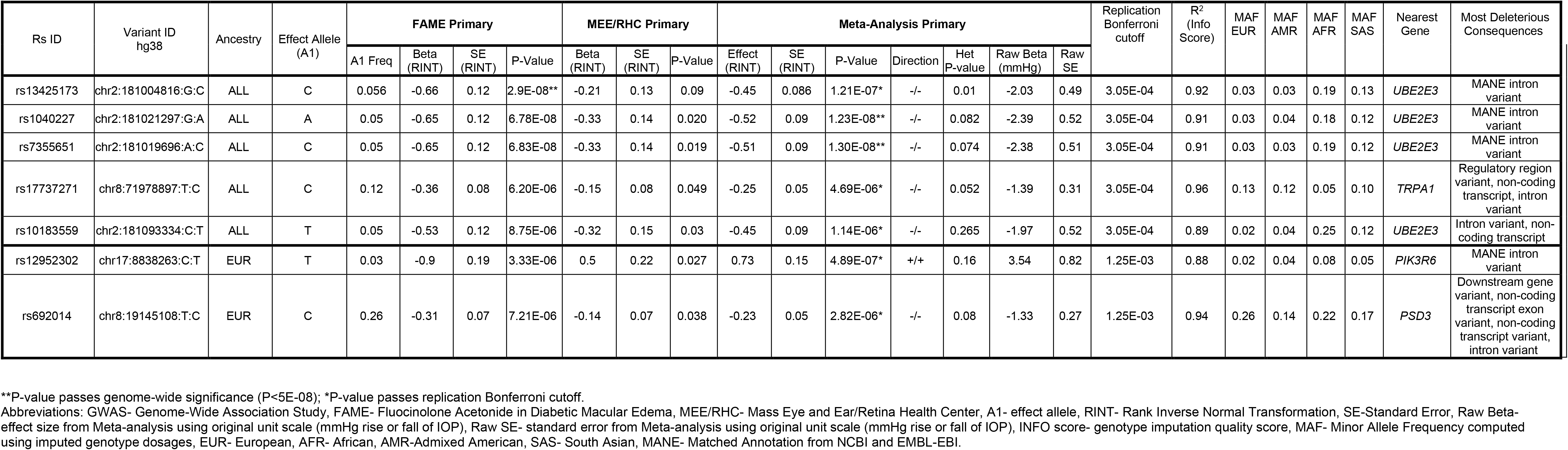
Top variants associated with glucocorticoid-induced intraocular pressure rise (IOP). The table shows all the variants that were genome-wide significant (P < 5×10^-8^) and/or replicated in both cohorts- Fluocinolone Acetonide in Diabetic Macular Edema (FAME) discovery cohort and Mass Eye and Ear/Retina Health Center (MEE/RHC) replication cohort. Listed are the summary statistics for GWAS in discovery FAME cohort, replication MEE/RHC cohort and the FAME-MEE/RHC meta-analysis sorted by their P-values. The Effect Allele (A1), Effect Allele’s frequency (A1 Freq), P-values, direction of effect, heterozygosity P-value, replication Bonferroni cutoff and genotype imputation quality (R^2^, INFO score) are provided. The table also shows RINT’ed and raw effect size (Beta/Effect) and standard error (SE) from GWAS and/or Meta-analysis using rank inverse normal transformed (RINT) maximal IOP rise and pre-transformed maximal IOP rise in original unit scale (mmHg), respectively. Minor allele frequency (MAF) of the variant within each ancestry was calculated using the study population in which the variant was genome-wide significant or passed replication Bonferroni cutoff. The nearest gene and most deleterious consequences were annotated using Ensembl’s Variant Effect Predictor (VEP). All the variants within the *UBE2E3* locus listed below were in high linkage disequilibrium (LD) with each other.

Inspecting the effect of this variant across ancestral groups and cohorts revealed a consistent direction of effect on IOP change across all ancestries and cohorts (Figure 4, Supplemental Figure 11). All three of these chromosome 2 variants are within the intronic region of *UBE2E3*, ubiquitin conjugating enzyme E2 E3 (Figure 3). The C allele for chr2:181004816:G:C is more common in the AFR and SAS ancestries (minor allele frequency [MAF] of 0.19 and 0.13, respectively) than the EUR or AMR ancestries (MAF∼0.03). Each copy of the C allele (minor allele) was associated with 2.41 mmHg *lesser* rise in IOP, on average, within 6 months of GC treatment (Table 2). Similarly, the A allele (minor allele) for chr2:181021297:G:A is more common in AFR and SAS ancestries. Each copy of the A allele was associated with 2.34 mmHg *lesser* rise in IOP, on average, within 6 months of GC treatment (Table 2).

**Figure 4.**
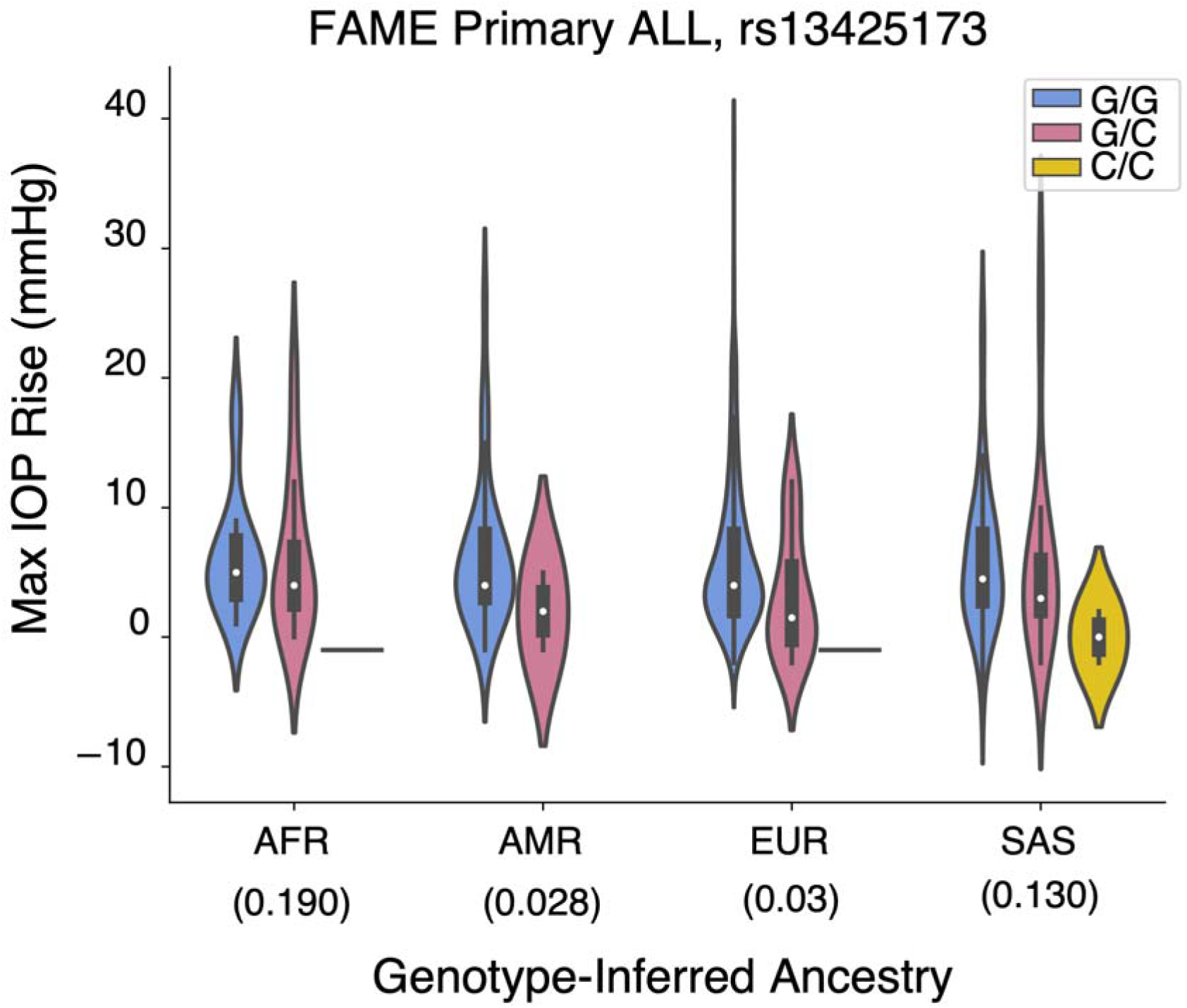
Violin plot showing distribution of maximal intraocular pressure (IOP) change within 6 months after glucocorticoid treatment stratified by genotype and ancestry for a genome-wide significant association (rs13425173, chr2:181004816:G:C) with the primary outcome across ALL ancestries in the Fluocinolone Acetonide in Diabetic Macular Edema (FAME) trial. AFR, African; AMR, Admixed American; EUR, European; SAS, South Asian

Full results for the LD-independent variants at P < 5X10^-7^ in the discovery cohort GWAS of the primary outcome are shown in Supplemental Table 10. LocusZoom plots for variants at P < 5 X 10^-7^ are shown in Supplemental Figure 12.

In addition to the *UBE2E3* locus, three independent loci (one in the ALL ancestry and two in the EUR ancestry) for the primary outcome in the discovery cohort showed nominal evidence of replication (P<0.05) in the MEE-RHC cohort and concordant direction of effect (Table 2). These three loci did not achieve genome-wide significance in the FAME-MEE/RHC meta-analysis, however they did reach the Bonferroni replication threshold (Table 2, Supplemental Figure 13). The first of these three loci, chr8:71978897:T:C (rs17737271), is near the protein-coding gene *TRPA1,* Transient receptor potential (TRP) ankyrin 1, although it resides within the non-coding gene *MSC-AS1* (see LocusZoom plot in Supplemental Figure 14). The second locus at chr17:8838263:C:T (rs12952302) lies within the gene *PIK3R6,* Phosphoinositide-3-kinase regulatory subunit 6. This variant passed genome-wide significance in the REGENIE GWAS of primary outcome across ALL ancestries (Supplemental Table 9). The third locus at chr8:19145108:T:C (rs692014), is near the protein-coding gene *PSD3,* Pleckstrin and Sec7 domain containing 3, but resides within the non-coding gene *RP11-1080G15.1.* The distribution of IOPs stratified by genotype and ancestry for these variants reveal consistent direction of effect across ancestries (Supplemental Figures 11, 15). There were no genome-wide significant variants in the meta-analyses for the secondary outcomes or injection-only sensitivity analyses.

The results of all LD-independent variant associations at P<5X10^-7^ in the FAME-MEE/RHC meta-analysis for all outcomes and ancestries are shown in Supplemental Table 11. The results of the LD-independent variant associations at P<10^-5^ for the MEE/RHC replication cohort generated for the meta-analyses are provided in the Supplemental Materials, Supplemental Figure 16, and Supplemental Table 12 (available at https://www.aaojournal.org).

### Fine mapping of the *UBE2E3* locus

To narrow down the most likely causal variants within the genome-wide significant loci, we performed fine-mapping on the discovery GWAS and Meta-analysis using SuSiE.^26^ For both the studies, we identified one 95% credible set with posterior probability >0.95 that included four variants in very high LD mapping to the intronic region of the *UBE2E3* gene (Supplemental Figure 17). Of these three variants, chr2:181004816:G:C, chr2:181019696:A:C, and chr2:181021297:G:A are the same genome-wide significant variants described above - one from the discovery cohort and two from the FAME-MEE/RHC meta-analysis all mapping to the intronic region (MANE) of *UBE2E3*. The fourth variant, chr2:180979454:T:C (FAME beta=-0.59, SE.=0.11, P=9.7X10^-8^) maps both to an upstream (MANE) and intronic region of *UBE2E3*.

### Colocalization Analyses of GWAS Loci (Common Variants)

To prioritize putative causal regulatory mechanisms and genes that may underlie the genetic associations with IOP change within 6 months of GC treatment, we tested if our top candidate variants colocalized with expression quantitative trait loci (eQTLs) in retina or eQTLs and splicing QTLs (sQTLs) in 49 non-ocular tissues in Genotype-Tissue Expression (GTEx) v8, many of which contain relevant cell types. For this analysis, we considered the loci around the genome-wide significant GWAS variant and replicated variants listed in Table 2. Three variants, two on chromosome 2 (*UBE2E3*) and one on chromosome 17 significantly colocalized with eQTLs or sQTLs (Table 3, Supplemental Tables 13-14, and Supplemental Figure 18). The significant GWAS variant from the discovery primary analysis (chr2:181004816:G:C) that falls in *UBE2E3* was associated with the expression of this gene in six different tissues including aorta (Figure 5 and Supplemental Figure 18). Increase of *UBE2E3* expression is suggested to be associated with decrease in IOP change following GC treatment (Table 3). The other *UBE2E3* variant, chr2:181021297:G:A, colocalized with a tibial nerve eQTL that targets *SSFA2*, sperm specific antigen 2 (also known as *ITPRID2*; Figure 30C,D). Increased *SSFA2* expression is suggested to increase IOP following GC exposure (Table 3). Similarly, the replicated locus, chr17:8838263:C:T, colocalized with three different genes including *GUCY2D* in retina.

**Figure 5.**
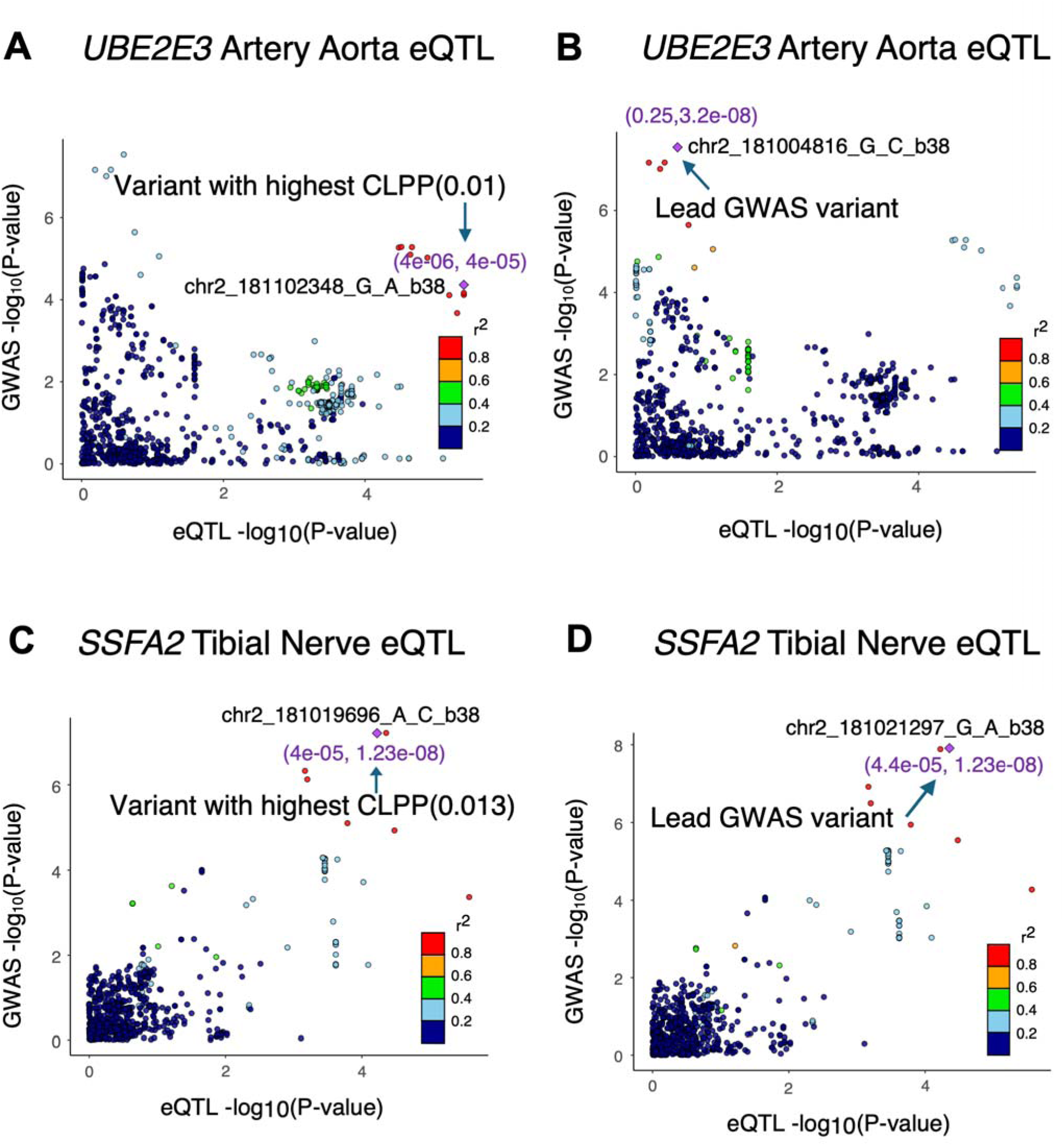
LocusCompare plots for two genome-wide significant loci associated with GC-induced intraocular pressure change that significantly colocalized with expression quantitative trait loci (eQTLs). -log_10_[genome-wide association study (GWAS} P value] is plotted as a function of - log_10_(eQTL P value) for **(A,B)** the Fluocinolone Acetonide Diabetic Macular Edema (FAME) trial primary GWAS across ALL ancestries in locus chr2:181004816:G:C versus the *UBE2E3* artery aorta eQTL, and **(C,D)** the FAME - Mass Eye and Ear/Retina Health Center (MEE/RHC) meta-analysis for the primary outcome across ALL ancestries versus the *SSFA2* Nerve Tibial eQTL. The eQTLs are from GTEx v8. The points represent the different variants in the GWAS locus [linkage disequilibrium (LD) interval, r^2^>0.01 and ±50kb] and are color-coded based on their LD (r^2^) relative to the eQTL variant with the most significant (highest) eCAVIAR colocalization posterior probability **(A,C)** or the lead GWAS variant **(B,D)**, represented by the purple diamond. The values inside the brackets in purple represent the eQTL and GWAS P values for the representative variant, respectively.

**Table 3.** GWAS loci that significantly colocalized with expression or splicing quantitative trait loci in retina and other non-ocular tissues. This table contains the significant eCAVIAR colocalization results for genome-wide significant GWAS variants in discovery cohort and variants that were replicated in both cohort by their trait/analysis, QTL type, target gene and tissue type tested across 49 GTEx tissues and peripheral retina. Significance was determined at a colocalization posterior probability (CLPP) > 0.01. The GWAS variant tested for colocalization is provided in column ‘Lead GWAS variant (hg38)’. The colocalizing e/sQTL variant with the highest CLPP is provided in column ‘e/sQTL variant (hg38)’. To eliminate potential false positives, only cases where the GWAS p-value of colocalization e/sVariant was below or equal to 0.05 or whose e/sQTL p-values was below or equal to 10^-4^ and/or did passed FDR below 0.05 were reported. All e/sQTL variants within a linkage disequilibrium (LD) interval (r^2^>0.1 plus 50 kb on either side) around each lead GWAS variant were tested in the colocalization analysis.

| Trait | Lead GWAS variant (hg38) | Replicated GWAS loci in both cohorts | GWAS P-value | e/sQTL variant (hg38) | CLPP | Direction of effect | Colocalizing e/sQTL target genes | e/sQTL tissue |
| --- | --- | --- | --- | --- | --- | --- | --- | --- |
| Genome-wide Significant GWAS Loci (P<5x10<sup>-8</sup>) |  |  |  |  |  |  |  |  |
| FAME Primary ALL | chr2:181004816:G:C | No | 2.88E-08 | chr2:181029486:T:A | 0.1 | Opposite | *UBE2E3* (e) | Minor Salivary gland, Artery Aorta, Breast Mammary Tissue, Pancreas, Nerve Tibial, Esophagus Mucosa |
| FAME-MEE/RHC Meta-analysis Primary ALL | chr2:181021297:G:A | Yes | 1.23E-08 | chr2:181021297:G:A | 0.01 | Same | *SSFA2 (ITPRID2)* (e) | Nerve Tibial |
| Replicated GWAS Loci |  |  |  |  |  |  |  |  |
| FAME-MEE Meta-analysis Primary EUR | chr17:8838263:C:T | Yes | 4.89E-07 | chr17:8080839:A:C | 0.04 | Opposite | *GUCY2D* (e) | Retina |
| FAME-MEE Meta-analysis Primary EUR | chr17:8838263:C:T | Yes | 4.89E-07 | chr17:8778482:G:A | 0.02 | Opposite | *MFSD6L* (e) | Pituitary |
| FAME-MEE Meta-analysis Primary EUR | chr17:8838263:C:T | Yes | 4.89E-07 | chr17:8851364:T:C | 0.02 | Same | *PIK3R6* (e) | Testis |
| FAME-MEE Meta-analysis Primary EUR | chr17:8838263:C:T | Yes | 4.89E-07 | chr17:8851364:T:C | 0.02 | Opposite | *PIK3R6* (s) | Testis |

Abbreviations: eQTL- expression quantitative trait loci, sQTL- splicing quantitative trait loci, □CLPP-colocalization posterior probability computed with eCAVIAR, (e)- eQTL, (s)- sQTL. Retina eQTLs are from EyeGEx and non-ocular tissue eQTLs and sQTLs are from GTEx v8.

Increased expression of *GUCY2D,* Guanylate Cyclase 2D, in retina is proposed to decrease GC-induced IOP in the combined FAME-MEE/RHC cohort. Additional colocalization results for subthreshold discovery GWAS and meta-analysis GWAS loci are detailed in the Supplemental Materials, Supplemental Table 13-14.

### Common variant GWAS Gene-Level and Pathway Analysis

In the gene-level association analysis using MAGMA, three genes were associated with GC-induced IOP change at subthreshold significance at P < 5 X 10^-5^ in the discovery cohort (Table 4), including *UBE2E3*. In the gene set-level analysis, the most notable and significantly associated pathways were the postsynaptic modulation of chemical synaptic transmission (P=2.2 X 10^-8^; Supplemental Table 15), protein deglutamylation (P=2.3 X 10^-7^), NLRP3 inflammasome complex assembly (P=4.8 X 10^-6^), and synthesis of prostaglandins and thromboxanes (P=2.6 X 10^-5^).

**Table 4.** Gene-based association analysis using MAGMA: Top genes (P<5 X 10^-5^) for Fluocinolone Acetonide in Diabetic Macular Edema (FAME). Listed are the top genes passing gene-based association analysis with their genomic position, the number of variants mapped to the gene (Number of variants), no. of samples used in GWAS (N), no. of parameters used in the model (NPARAM), Z-score (Z-statistic) and P-values. The column “Genome-wide association study significant variant” lists the variants within the gene that was genome-wide significant in the GWAS analysis and “Significant expression or splicing quantitative trait loci colocalization” highlights genes that also showed significant colocalization with expression or splicing quantitative trait loci.

| Trait | Population | Gene Entrez ID | Symbol | Chr | Start | Stop | Number of variants | NPARAM | N | Z-statistic | P-value | Genome-wide association study significant variant | Significant expression or splicing quantitative trait loci colocalization |
| --- | --- | --- | --- | --- | --- | --- | --- | --- | --- | --- | --- | --- | --- |
| FAME Primary | ALL | 10477 | UBE2E3 | 2 | 180880385 | 181163427 | 1002 | 67 | 530 | 4.4 | 6.6E-06 | 2:181021297:G:A | Yes |
| FAME Primary | ALL | 728276 | CLEC19A | 16 | 19185755 | 19424537 | 1020 | 108 | 530 | 4.1 | 2.2E-05 | 16:19391115:C:T | No |
| FAME Primary | ALL | 79838 | TMC5 | 16 | 19310522 | 19599113 | 1256 | 85 | 530 | 4.0 | 3.17E-05 | 16:19391115:C:T | No |

Abbreviations: MAGMA- Multi-marker Analysis of GenoMic Annotation, FAME=Fluocinolone Acetonide in Diabetic Macular Edema, NPARAM=number of principal components retained after pruning in the model; N=GWAS sample size. Bonferroni-corrected significance at P < 2.5 x 10<sup>-6</sup>.

### Single-cell gene expression in the anterior segment of the eye (Common Variants)

To further investigate the functional relevance of genome-wide significant and subthreshold loci, we looked at the expression levels of the associated genes in these loci in a publicly available single cell RNA-seq dataset from the anterior segment of the eye^32^ (Figure 6 and Supplemental Figure 19 ). Of all genes (coding and non-coding) within the LD interval of the genome-wide significant locus, chr2:181021297:G:A, only *UBE2E3* is highly expressed in the tissues associated with IOP regulation, such as Schlemm’s canal endothelium, fibroblasts (from limbus), uveal melanocytes, vascular endothelium, and pericytes [derived from trabecular meshwork (TM), ciliary body (CB), and corneoscleral wedge] (Figure 6).

**Figure 6.**
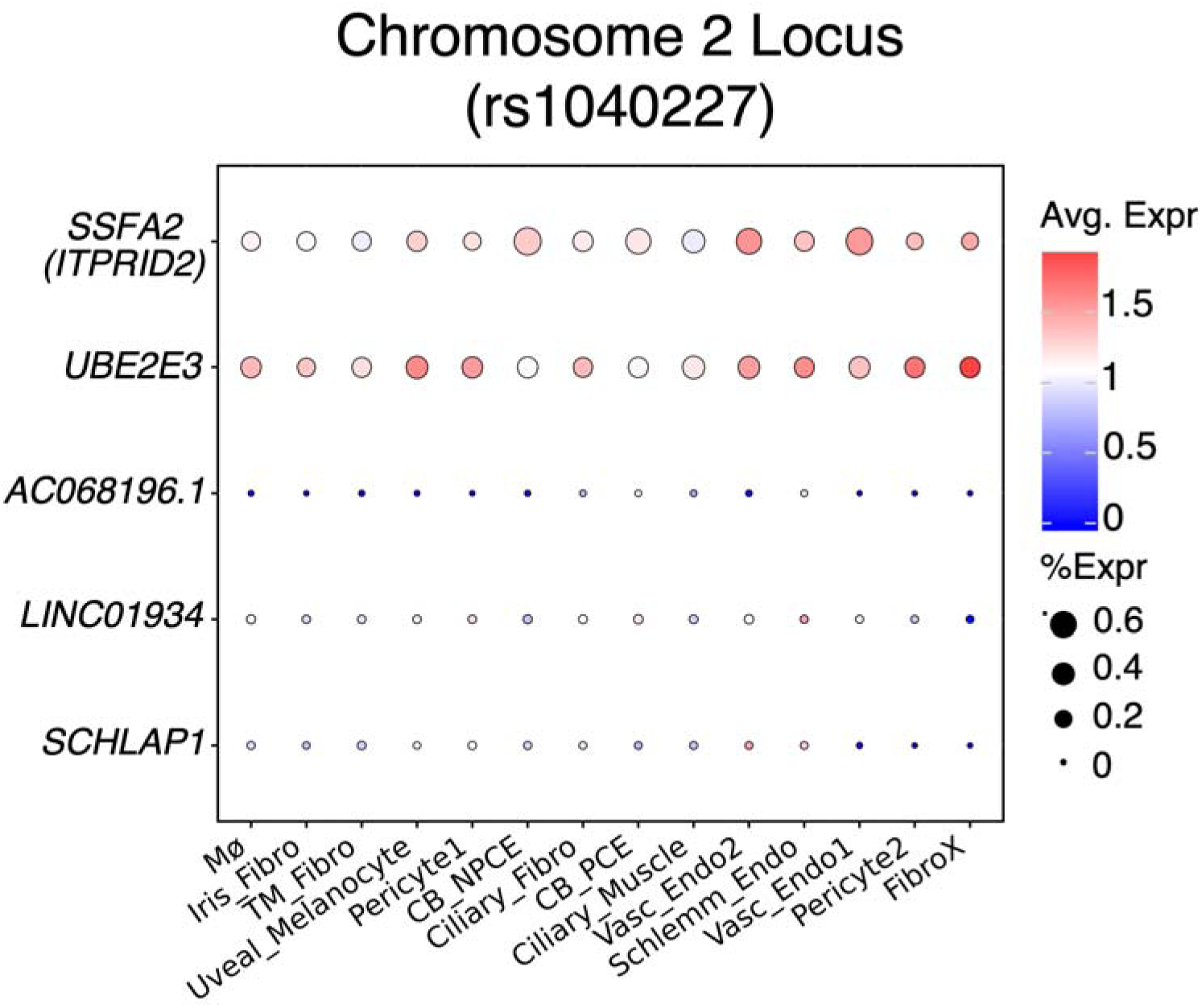
Bubble plot showing average single cell expression in anterior segment tissues previously associated with IOP of all (protein coding and non-coding) genes within the LD interval of the genome-wide significant locus, chr2:181004816:G:C (rs13425173) and *SSFA2* whose nerve tibial eQTL colocalized with the GWAS locus. *SCHLAP1* and *LINC01934* are non-coding genes present in the single cell data set that are within the LD interval of the chr2:181021297:G:A locus (as seen in the LocusZoom plot in Figure 3). The size of each circle is proportional to the percentage of nuclei within a cluster expressing the gene, and the color intensity represents the average normalized transcript count (log10(TPK+1)) in the expressing cells. Abbreviations: TPK=transcript per kilobase; Mø- marcrophage; Iris_Fibro- iris fibroblast; TM_Fibro - trabecular meshwork fibroblast; Uveal_Melanocyte- uveal melanocyte; Pericyte1-pericyte derived from trabecular meshwork and ciliary body; CB_NPCE- ciliary body nonpigmented ciliary epithelium; Ciliary_Fibro - ciliary fibroblast; CB_PCE - ciliary body pigmented ciliary epithelium; Ciliary_Muscle - ciliary muscle; Vasc_Endo2 - vascular endothelium 2; Schlemm_Endo – Schlemm’s Canal endothelium; Vasc_Endo1- vascular endothelium 1; Pericyte2- pericyte derived from corneoscleral wedge; FibroX- fibroblast from limbus.

### Whole Exome Sequencing Participant Characteristics

After sample and variant QC steps were applied, 532 discovery participants and 586 replication participants with high-quality whole exome sequences were included for the rare variant analyses (Supplemental Figure 2). The distributions of the GC-induced maximal IOP change values used for the gene burden testing are shown in Supplemental Figure 20. RINT transformation achieved normal distribution of IOP change. The genotype variance explained and ancestral distributions of the participants in the WES analyses are detailed in the Supplemental Materials and Supplemental Figures 21 and 22.

### Rare Variant Whole Exome Sequencing Analysis

The Q-Q plots for the primary and secondary outcomes for the WES analyses are shown in Supplemental Figure 23. The lambda values for the gene burden tests are shown in Supplemental Table 16. Of the gene burden and SKAT-O analyses, some analyses had lambda values between >1.1 or <0.9. These analyses were not considered to be reliable due to the small sample size or insufficiently correcting for population heterogeneity. In the discovery burden test using primary outcome, one gene, *MSTO1* (lambda =0.99) passed Benjamini-Hochberg FDR cutoff (FDR<0.15) in ALL ancestry in both gene burden and SKAT-O tests however none passed in the EUR subgroup (Supplemental Table 19). The different WES analyses performed as well as the summary table for the significant genes are shown in Supplemental Tables 17-19 and discussed in the Supplemental Materials.

We investigated whether the top genes (P<1 X 10^-3^) from discovery FAME cohort could be replicated in MEE/RHC cohort. Table 5 shows two candidates that were replicated in MEE/RHC cohort: *ZNF248, and LDHAP5*. *ZNF248* passed the replication Bonferroni cutoff in the replication cohort, while *LDHAP5* passed the nominal significance threshold. Both genes passed replication Bonferroni cutoff in the combined meta-analysis (P < 2.9 X 10^-3^and 2.2E X 10^-3^) with concordant direction of effect. In the meta-analysis three additional genes (*SPAG4, MYO10, and SLC52A3)* passed both Bonferroni and FDR cutoff (Supplemental Table 19).

**Table 5.**
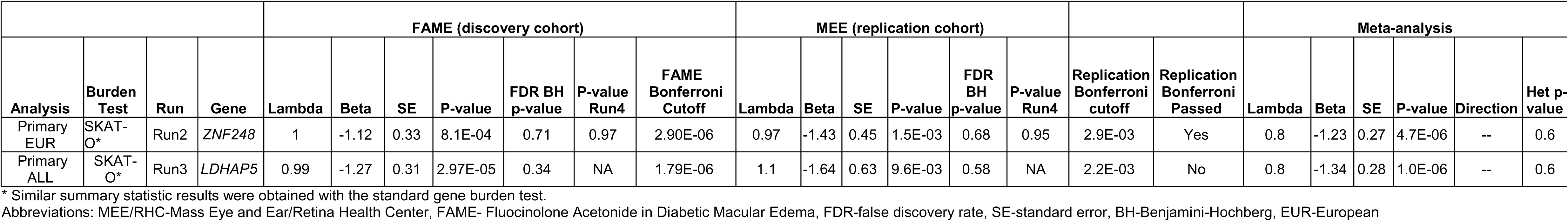
Exome data replication results: Top genes (P<10^-3^) from discovery burden analysis replicated in Mass Eye and Ear (MEE)/Retina Health Center samples. The table shows all the top genes from Fluocinolone Acetonide in Diabetic Macular Edema (FAME) discovery cohort that either passed replication Bonferroni or nominal significance in Mass Eye and Ear/Retina Health Center (MEE/RHC) replication cohort by analysis/trait, type of burden test performed and Run (grouping of variants based on their functional impact to the gene). Listed are the summary statistics for discovery FAME cohort, replication MEE/RHC cohort and the FAME-MEE/RHC meta-analysis. The lambda values, effect size (Beta), standard error (SE), P-value, false discovery rate, Bonferroni cutoff within each study/cohort are provided. Also listed are the direction of effect and Heterozygosity P-value for the meta-analysis and replication Bonferroni cutoff. Column “P-value Run4” shows the P-value in the negative control group Run4 (group of synonymous variants).

### Overlap with genes previously associated with IOP

We examined our top GWAS and WES associations for overlap with genes previously associated with IOP or related traits,^33–39^ and/or that have been reported to be differentially expressed in response to GC in eye tissues.^40,41^ The Supplemental Materials and Supplemental Tables 22 and 23 show the overlapping findings from the GWAS and WES gene burden testing analyses, respectively. Three subthreshold variants from discovery GWAS and eight from the meta-analysis resided in or near genes previously associated with POAG or IOP. These genes include *SDK1*, *ZNF516, TRPA1, PIEZO2, CCDC85A, ACTR5, FOXQ1, GAS7,* and *CALD1*.

From the discovery and meta-analysis, three genes with the lowest P-values were - *CCDC85A, PIEZO2,* and *SDK1.* The variant, rs6462562, mapping to *SDK1* gene was previously associated with POAG in African population.^37^ The *SDK1* variant is subthreshold in the FAME primary EUR GWAS (chr7:4108958:C:G, P=4.5 X 10^-7^) and is not in strong LD with the rs6462562 variant (r^2^= 0.02). *PIEZO2* encodes for a mechanosensitive channel protein previously associated with POAG.^36^ The *PIEZO2* locus with subthreshold association in the primary outcome meta-analysis (chr18:11313797:C:A, P=4.1 X 10^-7^) is not in LD with the POAG-associated variant (rs112014893, r^2^=0). *CCDC85A* (Coiled-Coil Domain Containing 85A) is a gene that has been consistently associated with POAG and IOP in several studies. The *CCDC85A* variant rs112926739 associated with GC-induced OHTN with a P value of 1.3 X 10^-6^ in the meta-analysis (Supplemental Table 11) is not in strong LD with the variants rs4672075 and rs7572523 associated with IOP previously (r^2^= 0.0036 and 0.004, respectively).^33,35^ When we examined genes known to be differentially expressed in human TM cells after GC treatment,^40,41^ we found two genes, *HIPK2* and *TMC5,* that have nearby variants (chr7:139570283:G:A and chr16:19391115:C:T, respectively) associated with the FAME Primary outcomes in ALL ancestry in our GWAS at a P-value of 4.2 X 10^-7^ and 1.5 X 10^-7^, respectively (Supplemental Table 22). *HIPK2* encodes Homeodomain Interacting Protein Kinase 2, a serine/threonine nuclear kinase. *TMC5* encodes Transmembrane Channel Like 5, which enables mechanosensitive monoatomic ion channel activity.

Among the WES results, a frameshift variant in *MYO10*, was significantly associated with Extreme Responder outcome from the meta-analysis in the EUR subgroup (Supplemental Table 23). This gene is known to play a role in IOP via its regulation of actin-based filopodia and tunneling nanotubes in TM and extracellular matrix turnover.^42^

## Discussion

Our GWAS analysis identified one genome-wide significant locus associated with GC-induced IOP change. Two variants in high LD within this locus (*UBE2E3)* reached the threshold for genome-wide significance – one in the discovery GWAS (chr2:181004816:G:C) and another in the meta-analysis across the two cohorts, FAME and MEE/RHC (chr2:181021297:G:A). Three additional loci near the genes *TRPA1, PIK3R6* and *PSD3* were also statistically associated after meta-analysis with the replication cohort. In the gene-level association analyses, the same gene *UBE2E3* passed sub-threshold significance. Colocalization analysis with eQTLs and sQTLs in retina and GTEx tissues identified two target genes in this locus, *UBE2E3* and *SSFA2,* whose expression change might be associated with GC-induced OHTN.

*UBE2E3* encodes ubiquitin conjugating enzyme E2 E3. The modification of proteins with ubiquitins is an important cellular mechanism for targeting abnormal or short-lived proteins for degradation. *UBE2E3* also demonstrated nominal association with GC-induced OHTN in the gene-based MAGMA analysis (P=6.6 X 10^-6^). *UBE2E3* is also enriched in tissues relevant to IOP in the single-cell expression dataset from the anterior segment of the eye.^32^ These multiple lines of evidence suggest that this gene may play a role in GC-induced OHTN. There is experimental data that ubiquitin plays a central role in the turnover of proteins in the TM and that altered turnover of these proteins is key in the pathogenesis of GC-induced OHTN.^43^

There were three other loci which were replicated at a nominal significance level. The first of these replicated loci (chr8:71978897:T:C; rs17737271) is near the gene *TRPA1*. *TRPA1* is a member of the transient receptor protein (TRP) family of ion channels functioning in sensation and transduction of mechanical stress and inflammation, and has been implicated in maintaining IOP.^44, 45^ Both common and rare variants (rs2587561, rs61758122) within this gene have been associated with glaucoma.^36^ However, these two variants were not in LD with our replicated variant, chr8:71978897:T:C. The second variant, chr17:8838263:C:T, lies within the gene *PIK3R6*, or phosphoinositide-3-kinase regulatory subunit 6; phosphoinositide 3-kinase gamma is a lipid kinase. This variant colocalized with an eQTL in retina associated with the expression of *GUCY2D*, a retina-specific guanylate cyclase, and with eQTLs regulating *PIK3R6 and MFSD6L* in gland tissues. The third variant, chr8:19145108:T:C, is near the gene, *PSD3,* or Pleckstrin And Sec7 Domain Containing 3, which is expressed in the TM cell-extracellular matrix complex involved in modulating cell junctions and is known to be induced by TGF-beta in TM.^46,47^

The WES rare variant gene burden testing found a different set of genes compared to the common variant GWAS analyses. When applying the multiple testing FDR correction, one gene, *MSTO1,* a gene involved in the regulation of mitochondrial morphology,^48^ was significantly enriched for rare variants with missense and modifier effects associated with GC-induced OHTN in the discovery cohort, however this gene association did not replicate in the MEE/RHC cohorts. Two other genes (*ZNF248* and *LDHAP5*) did demonstrate association across both FAME and MEE/RHC. *ZNF248,* which passed the replication Bonferroni cutoff, is a member of the zinc finger protein family functioning as a regulatory transcription factor. *LDHAP5* passed nominal significance and encodes lactase dehydrogenase A pseudogene 5. Its function is not well understood. However, due to the modest cohort size the gene burden analysis was underpowered and might have missed some biologically relevant associations.

The existence of several other POAG/IOP associated genes among the top findings of our GWAS and WES analyses implicates a shared genetic vulnerability to both non-GC-related and GC-related IOP change. The variants for GC-related IOP change are often necessarily the same variants for non-GC-related IOP in the same gene. The possibility that different variants in the same genes lead to differential susceptibility to GC-induced vs. primary IOP variation needs to be explored in larger datasets.

To the best of our knowledge, this is the largest GWAS and the first WES study to examine GC-induced OHTN. Its strengths include the well-phenotyped FAME cohort, which had rigorous clinical trial collection of data, and the comprehensive nature of the genetic analyses performed. Despite being the largest GWAS to date, the sample size is relatively small for GWAS and thus there is limited power to detect variants of modest effects. There was heterogeneity of GC delivery methods and IOP measurement methods in the MEE/RHC cohort, which could also have diminished the power of the study. We did try to mitigate the heterogeneity of the GC delivery methods in part by doing a sensitivity analysis examining only the participants who received intravitreal GC in the MEE/RHC cohort. We also tried to mitigate the variability in IOP measurement methods by taking measurements from the same method for any given participant. While we adjusted for many of the covariates relevant to this phenotype, including GC dose/delivery mode and IOP-lowering drop use during the first six months, there is still the potential for unmeasured or incompletely captured confounding effects. For example, it is possible the maximal IOP was affected by IOP-lowering drop use to varying degrees and adjusting for IOP-lowering drug use as a dichotomous covariate did not capture all the variability this medical intervention could introduce. All the genome-wide significant findings were identified in the primary outcome analysis of maximal IOP Change. This is in keeping with the larger power to detect genetic variants with continuous versus dichotomous traits. Finally, we performed several GWAS analyses of different phenotype definitions or ancestral groups, which could increase the risk of false positive results from increased multiple hypothesis testing.

However, the different GWAS outcomes tested within each cohort are highly correlated and thus further Bonferroni correction (beyond what is already applied for the establishment of the genome-wide significance level of P<5X10^-8^) would be overly conservative. We tried to mitigate this multiple hypothesis correction risk by obtaining additional evidence of potential functional impact of our significant GWAS findings through colocalization analyses with eQTLs and sQTLs. Another limitation of our study relates to linkage disequilibrium (LD) reference panels used in colocalization analysis and fine mapping. Given the diverse ancestry composition in our cohort, we used multi-ethnic reference panels to compute LD, which may reduce power or introduce biases.

In summary, we provide several lines of evidence that support a role for common variation in *UBE2E3* in GC-induced IOP change. This genetic risk may be mediated through alteration of protein turnover in the TM and ciliary body. We also identified other common and rare variants that may influence GC-induced IOP change, which require further validation in larger datasets.

## Data Availability

Data generated by this study can be accessed on dbGAP, study ID phs004037.

## Acknowledgements

We gratefully acknowledge the physicians of the Mass Eye and Ear Glaucoma and Retina Services for facilitating the enrollment of patients from their practices in this study.

