## Supplementary Figures for "Genome-Wide Association Study for Glucocorticoid-Induced Ocular Hypertension"

### Slide 1
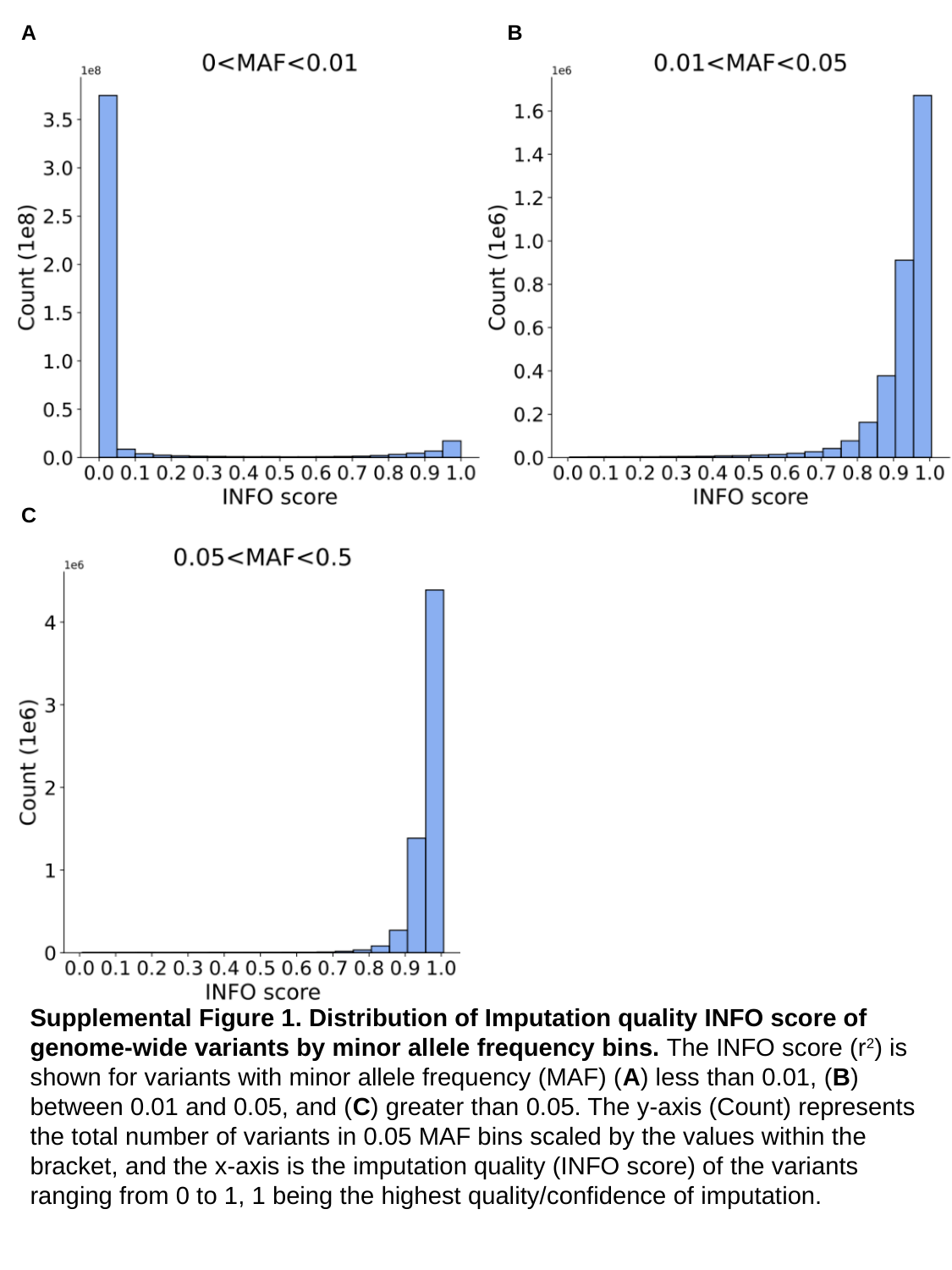

A
B
C
Supplemental Figure 1. Distribution of Imputation quality INFO score of genome-wide variants by minor allele frequency bins. The INFO score (r^2^) is shown for variants with minor allele frequency (MAF) (A) less than 0.01, (B) between 0.01 and 0.05, and (C) greater than 0.05. The y-axis (Count) represents the total number of variants in 0.05 MAF bins scaled by the values within the bracket, and the x-axis is the imputation quality (INFO score) of the variants ranging from 0 to 1, 1 being the highest quality/confidence of imputation.

### Slide 2
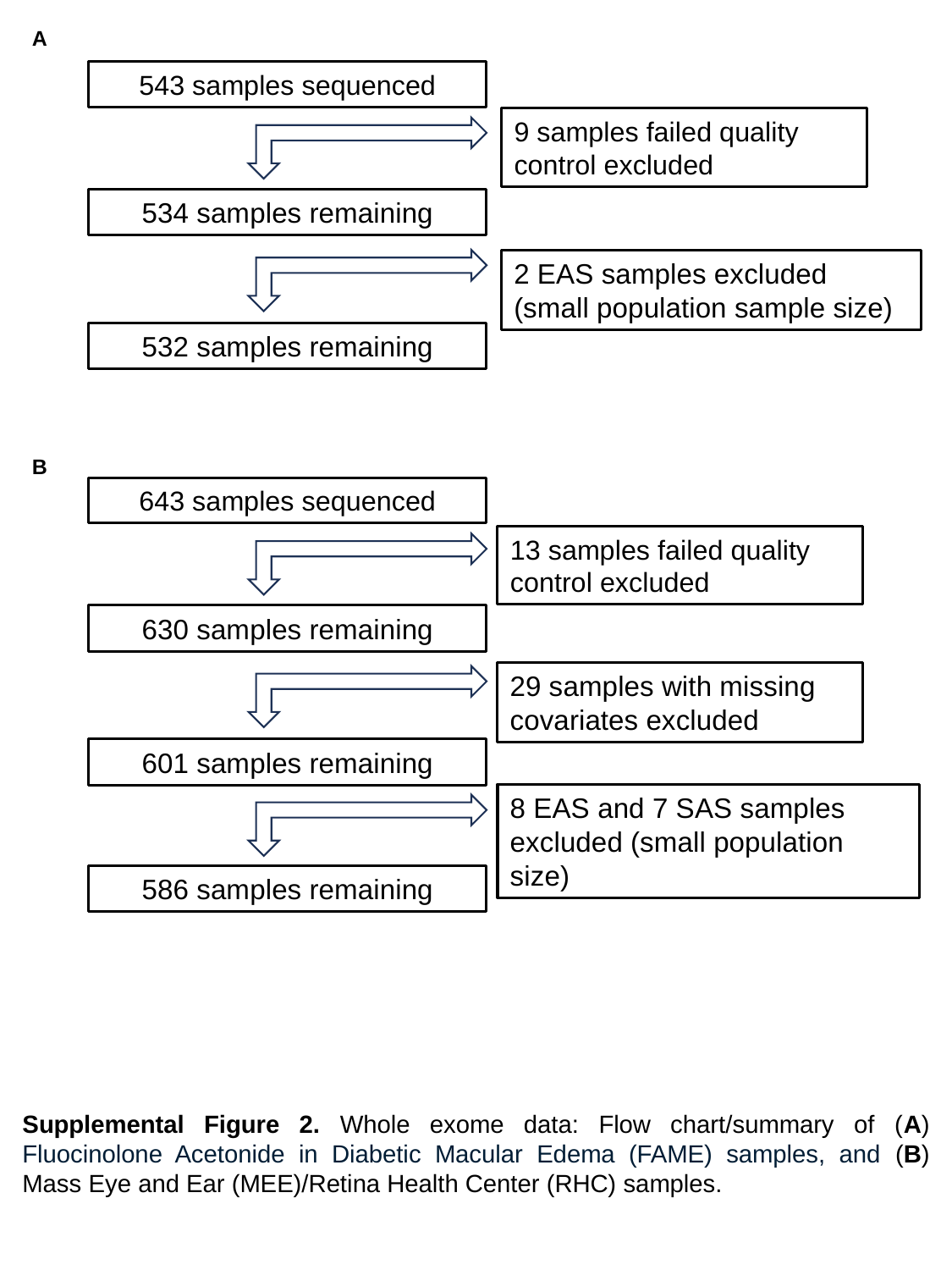

A
543 samples sequenced
9 samples failed quality control excluded
534 samples remaining
2 EAS samples excluded (small population sample size)
532 samples remaining
B
643 samples sequenced
13 samples failed quality control excluded
630 samples remaining
29 samples with missing covariates excluded
601 samples remaining
8 EAS and 7 SAS samples excluded (small population size)
586 samples remaining
Supplemental Figure 2. Whole exome data: Flow chart/summary of (A) Fluocinolone Acetonide in Diabetic Macular Edema (FAME) samples, and (B) Mass Eye and Ear (MEE)/Retina Health Center (RHC) samples.

### Slide 3
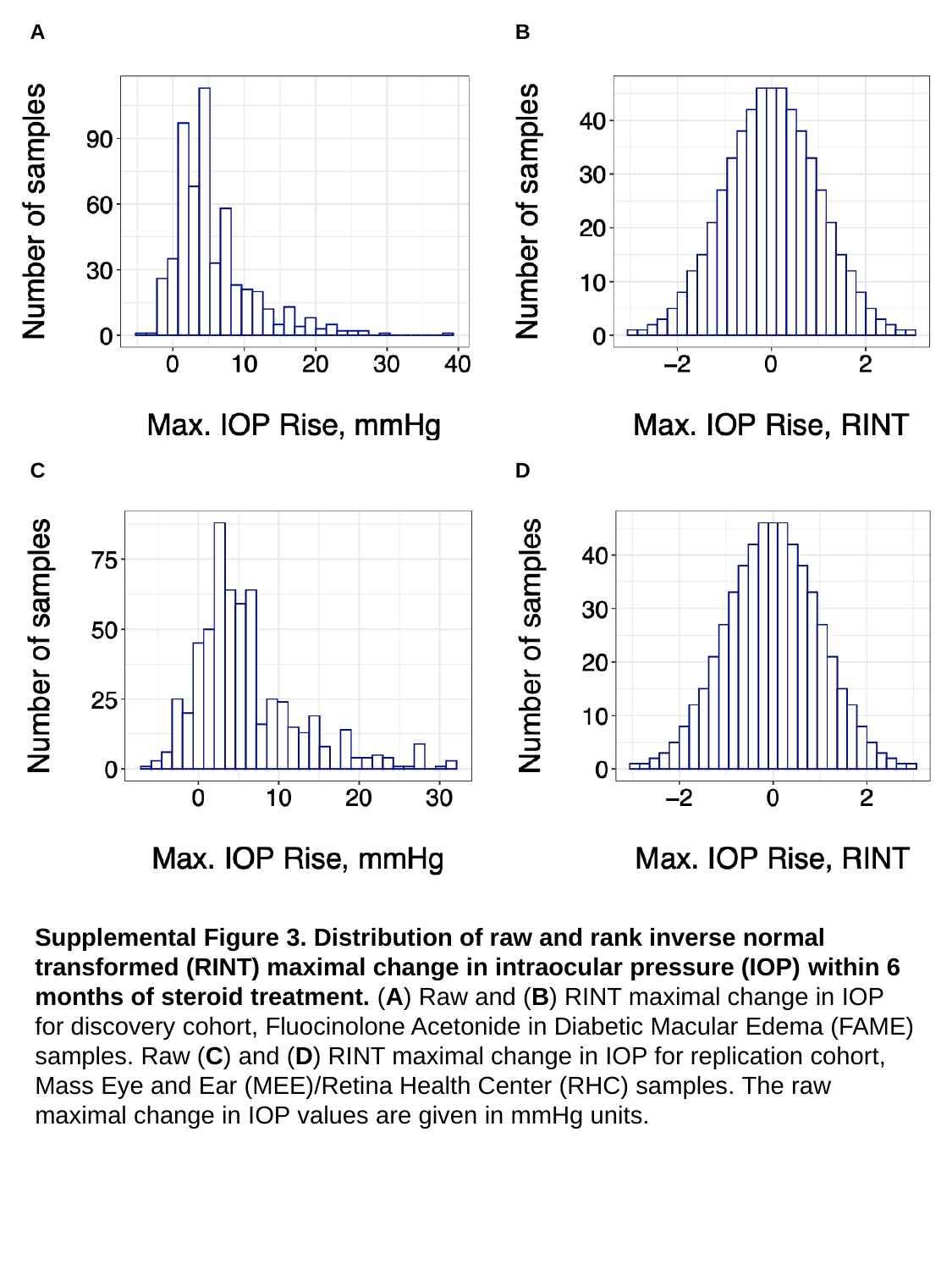

A
B
C
D
Supplemental Figure 3. Distribution of raw and rank inverse normal transformed (RINT) maximal change in intraocular pressure (IOP) within 6 months of steroid treatment. (A) Raw and (B) RINT maximal change in IOP for discovery cohort, Fluocinolone Acetonide in Diabetic Macular Edema (FAME) samples. Raw (C) and (D) RINT maximal change in IOP for replication cohort, Mass Eye and Ear (MEE)/Retina Health Center (RHC) samples. The raw maximal change in IOP values are given in mmHg units.

### Slide 4
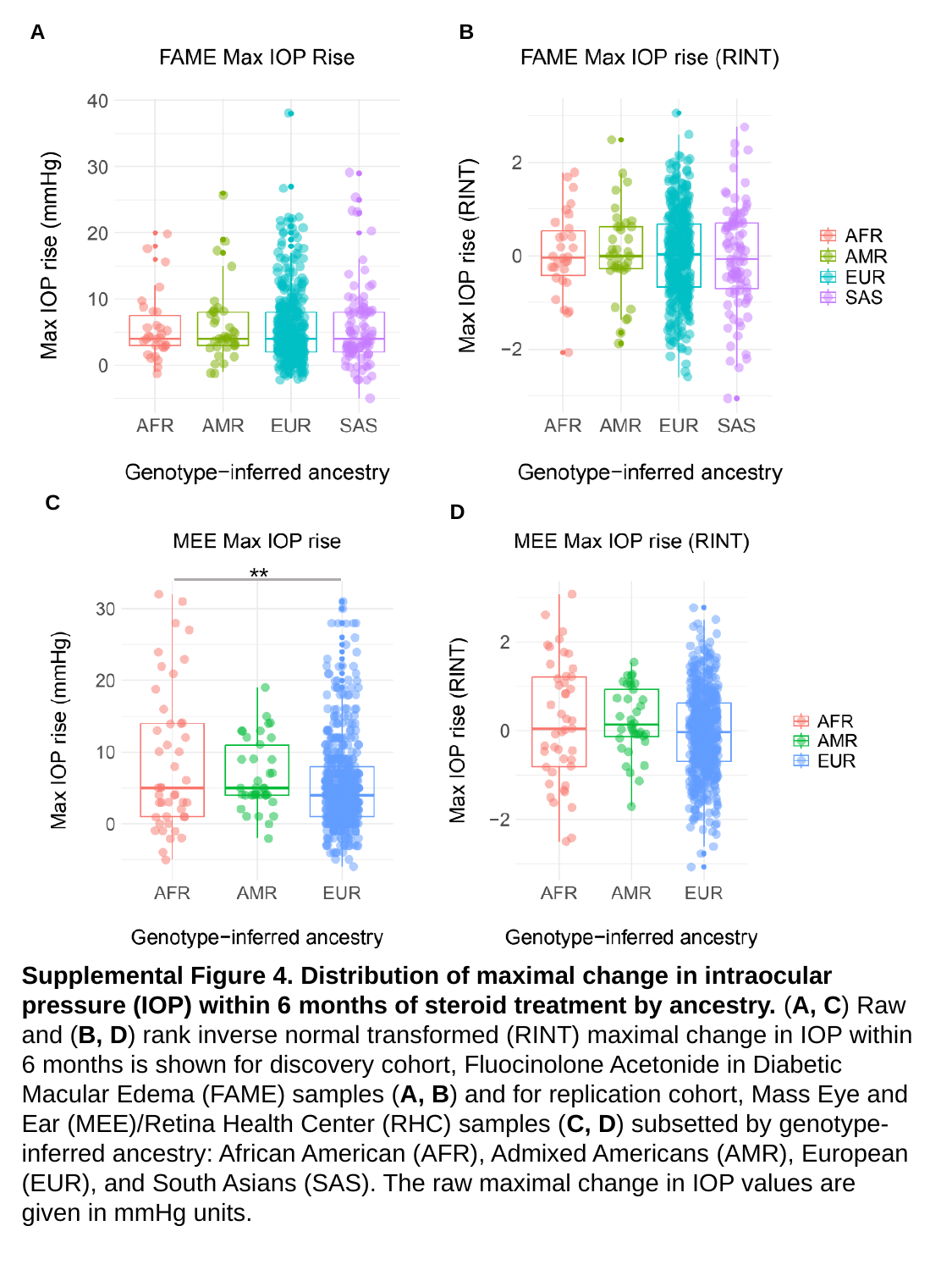

A
B
C
D
**
Supplemental Figure 4. Distribution of maximal change in intraocular pressure (IOP) within 6 months of steroid treatment by ancestry. (A, C) Raw and (B, D) rank inverse normal transformed (RINT) maximal change in IOP within 6 months is shown for discovery cohort, Fluocinolone Acetonide in Diabetic Macular Edema (FAME) samples (A, B) and for replication cohort, Mass Eye and Ear (MEE)/Retina Health Center (RHC) samples (C, D) subsetted by genotype-inferred ancestry: African American (AFR), Admixed Americans (AMR), European (EUR), and South Asians (SAS). The raw maximal change in IOP values are given in mmHg units.

### Slide 5
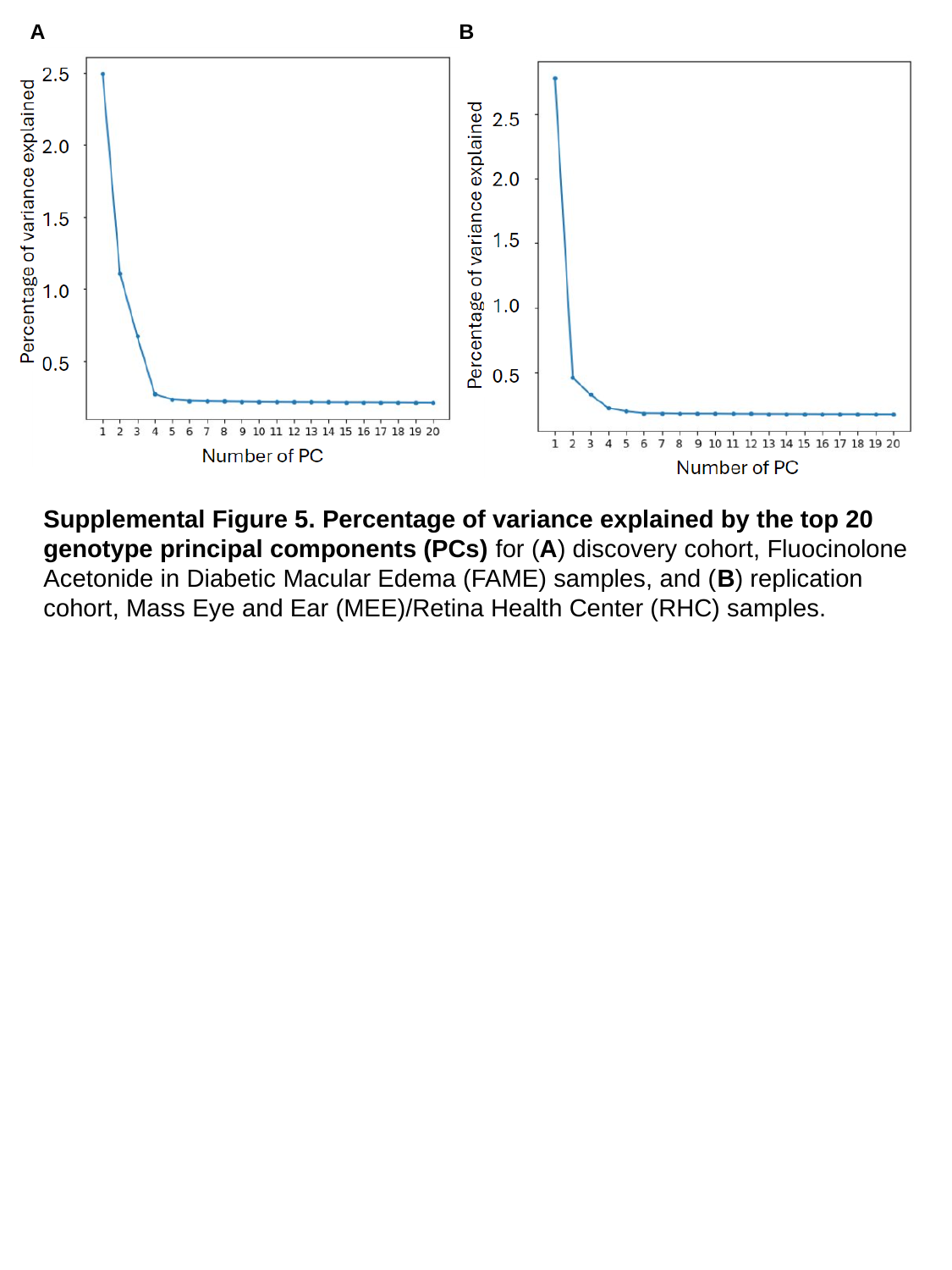

A
B
Supplemental Figure 5. Percentage of variance explained by the top 20 genotype principal components (PCs) for (A) discovery cohort, Fluocinolone Acetonide in Diabetic Macular Edema (FAME) samples, and (B) replication cohort, Mass Eye and Ear (MEE)/Retina Health Center (RHC) samples.

### Slide 6
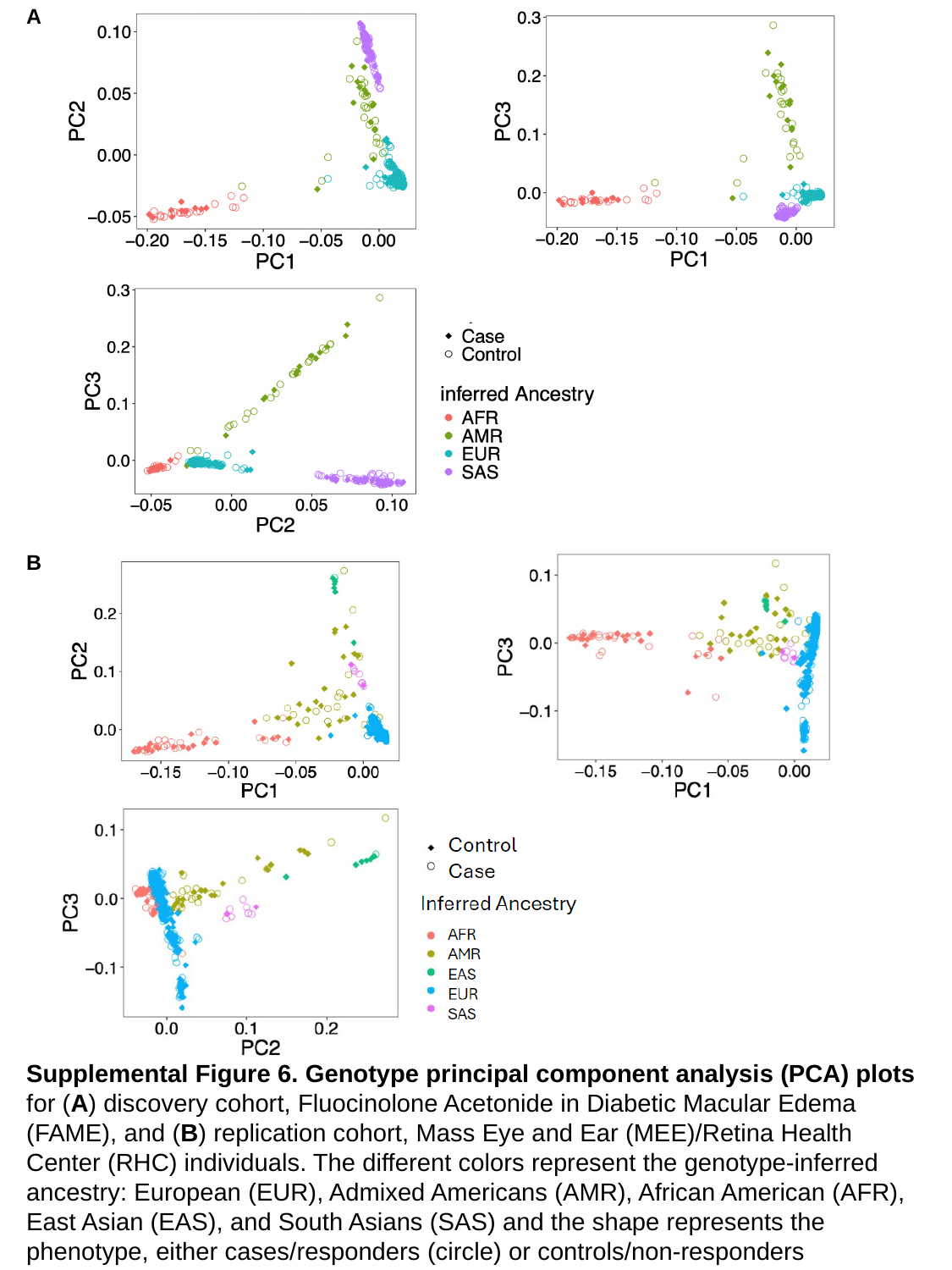

A
B
Supplemental Figure 6. Genotype principal component analysis (PCA) plots for (A) discovery cohort, Fluocinolone Acetonide in Diabetic Macular Edema (FAME), and (B) replication cohort, Mass Eye and Ear (MEE)/Retina Health Center (RHC) individuals. The different colors represent the genotype-inferred ancestry: European (EUR), Admixed Americans (AMR), African American (AFR), East Asian (EAS), and South Asians (SAS) and the shape represents the phenotype, either cases/responders (circle) or controls/non-responders (diamond).

### Slide 7
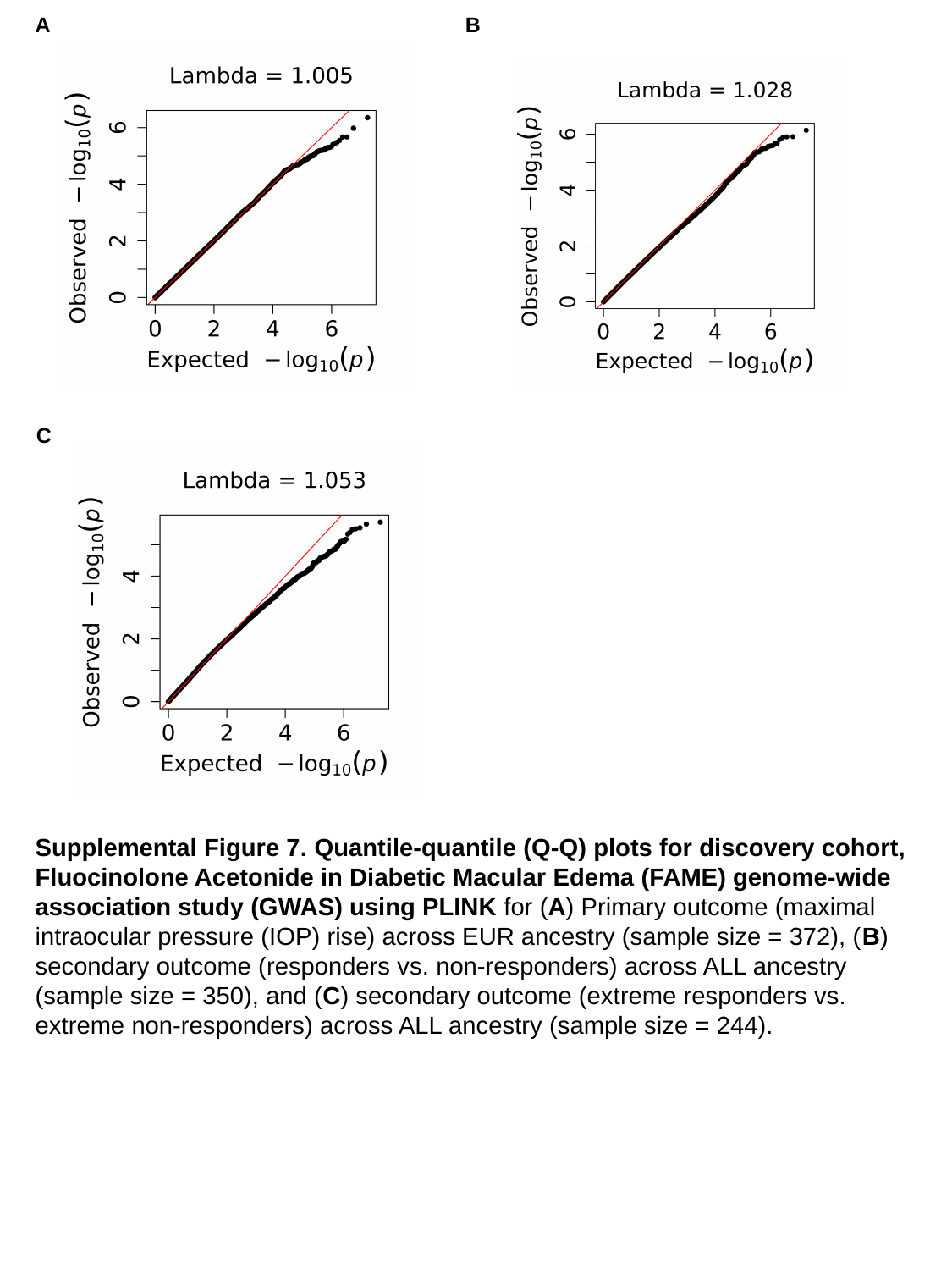

A
B
C
Supplemental Figure 7. Quantile-quantile (Q-Q) plots for discovery cohort, Fluocinolone Acetonide in Diabetic Macular Edema (FAME) genome-wide association study (GWAS) using PLINK for (A) Primary outcome (maximal intraocular pressure (IOP) rise) across EUR ancestry (sample size = 372), (B) secondary outcome (responders vs. non-responders) across ALL ancestry (sample size = 350), and (C) secondary outcome (extreme responders vs. extreme non-responders) across ALL ancestry (sample size = 244).

### Slide 8
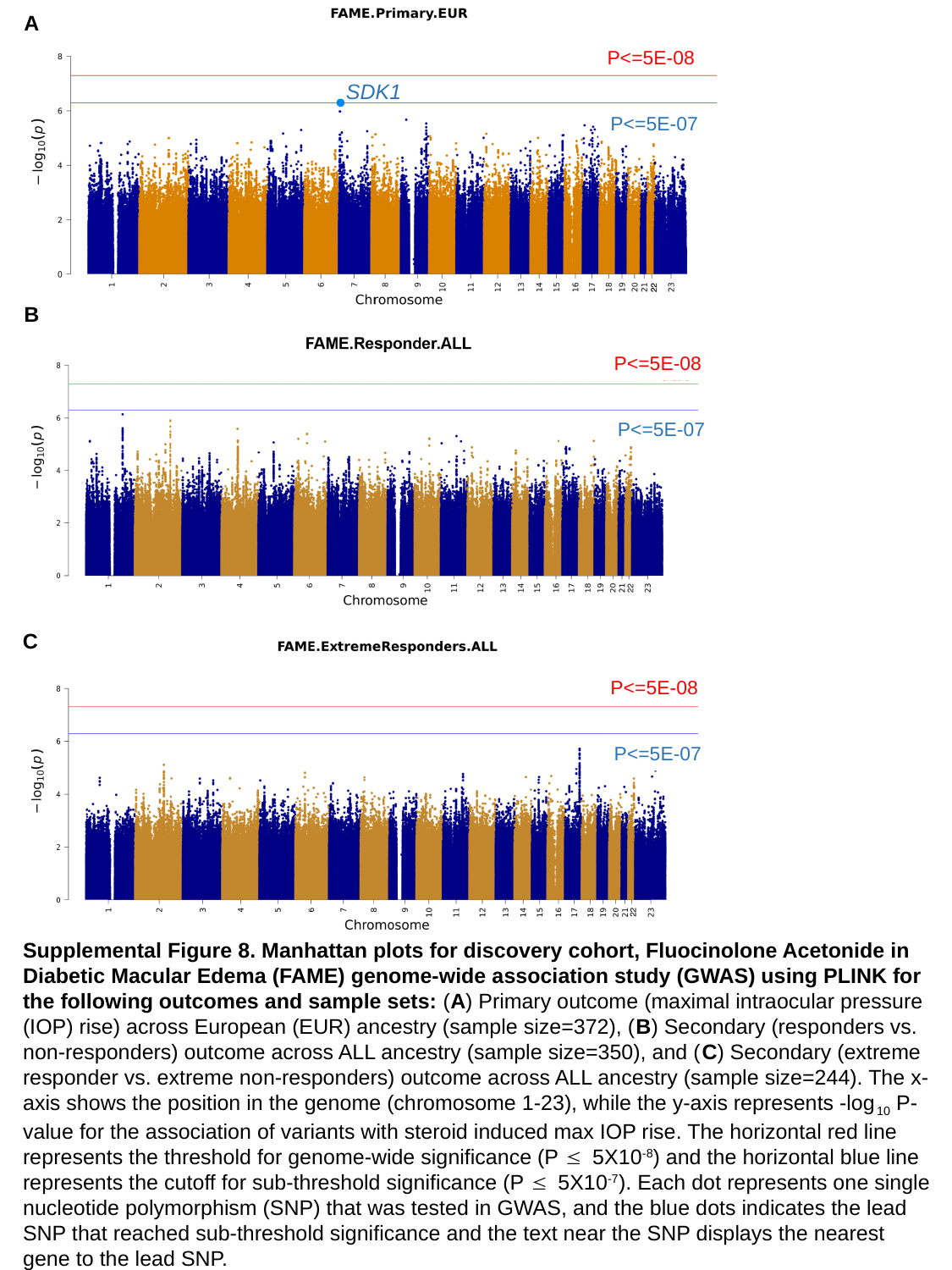

A
P<=5E-08
SDK1
P<=5E-07
B
P<=5E-08
P<=5E-07
C
P<=5E-08
P<=5E-07
Supplemental Figure 8. Manhattan plots for discovery cohort, Fluocinolone Acetonide in Diabetic Macular Edema (FAME) genome-wide association study (GWAS) using PLINK for the following outcomes and sample sets: (A) Primary outcome (maximal intraocular pressure (IOP) rise) across European (EUR) ancestry (sample size=372), (B) Secondary (responders vs. non-responders) outcome across ALL ancestry (sample size=350), and (C) Secondary (extreme responder vs. extreme non-responders) outcome across ALL ancestry (sample size=244). The x-axis shows the position in the genome (chromosome 1-23), while the y-axis represents -log_10_ P-value for the association of variants with steroid induced max IOP rise. The horizontal red line represents the threshold for genome-wide significance (P ≤ 5X10^-8^) and the horizontal blue line represents the cutoff for sub-threshold significance (P ≤ 5X10^-7^). Each dot represents one single nucleotide polymorphism (SNP) that was tested in GWAS, and the blue dots indicates the lead SNP that reached sub-threshold significance and the text near the SNP displays the nearest gene to the lead SNP.

### Slide 9
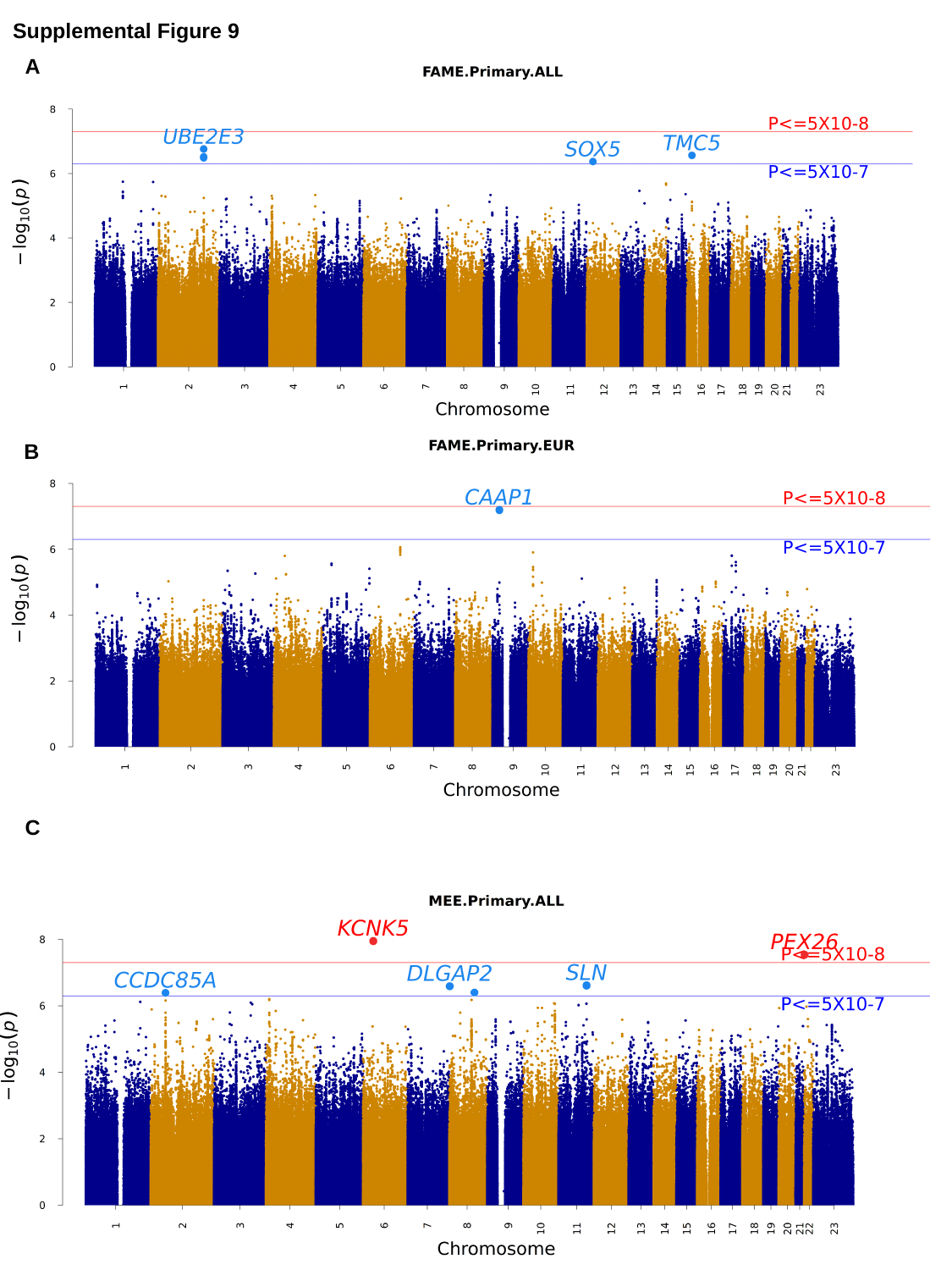

Supplemental Figure 9
A
B
C

### Slide 10
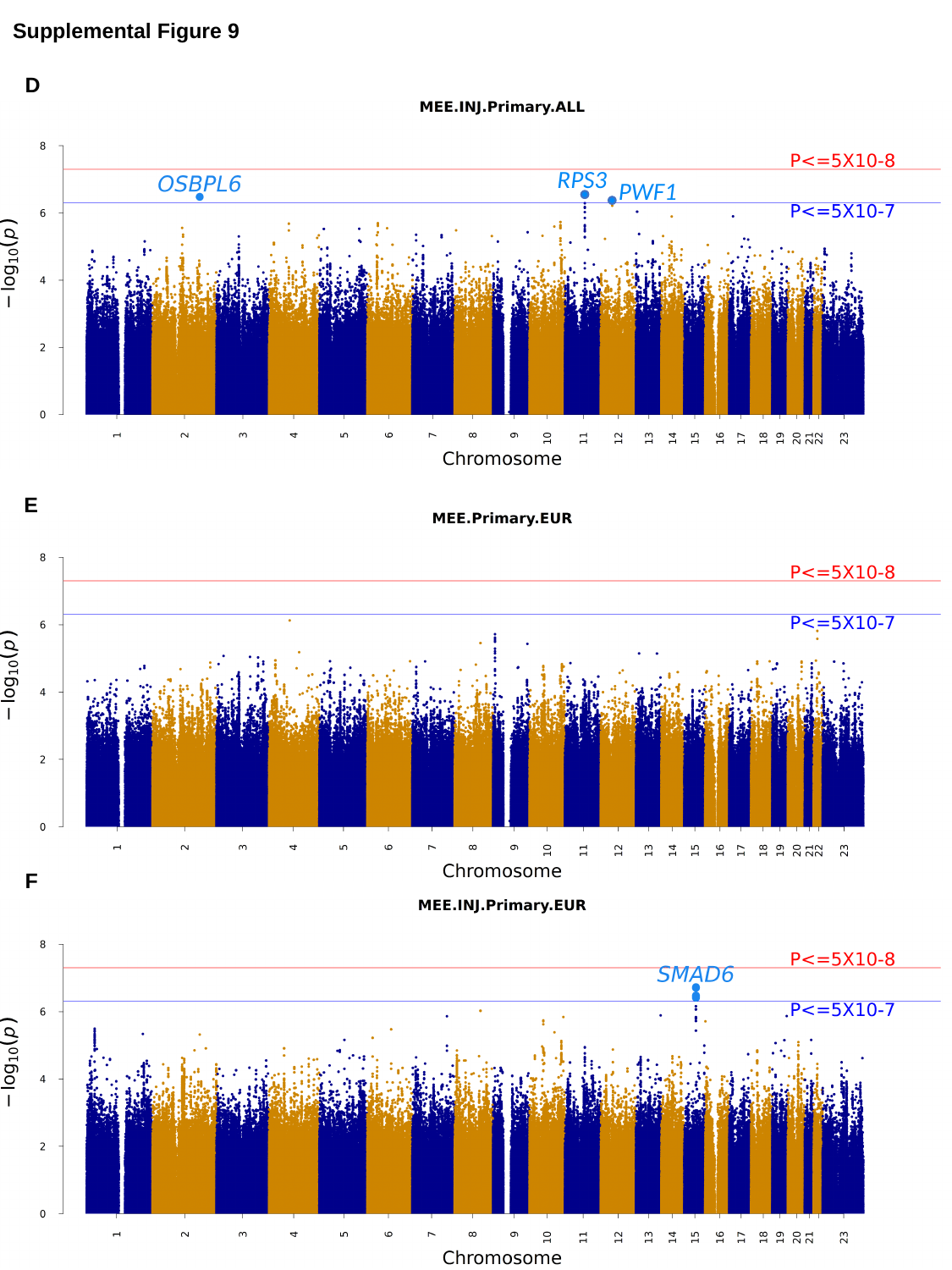

Supplemental Figure 9
D
RPS3
PWF1
E
F

### Slide 11
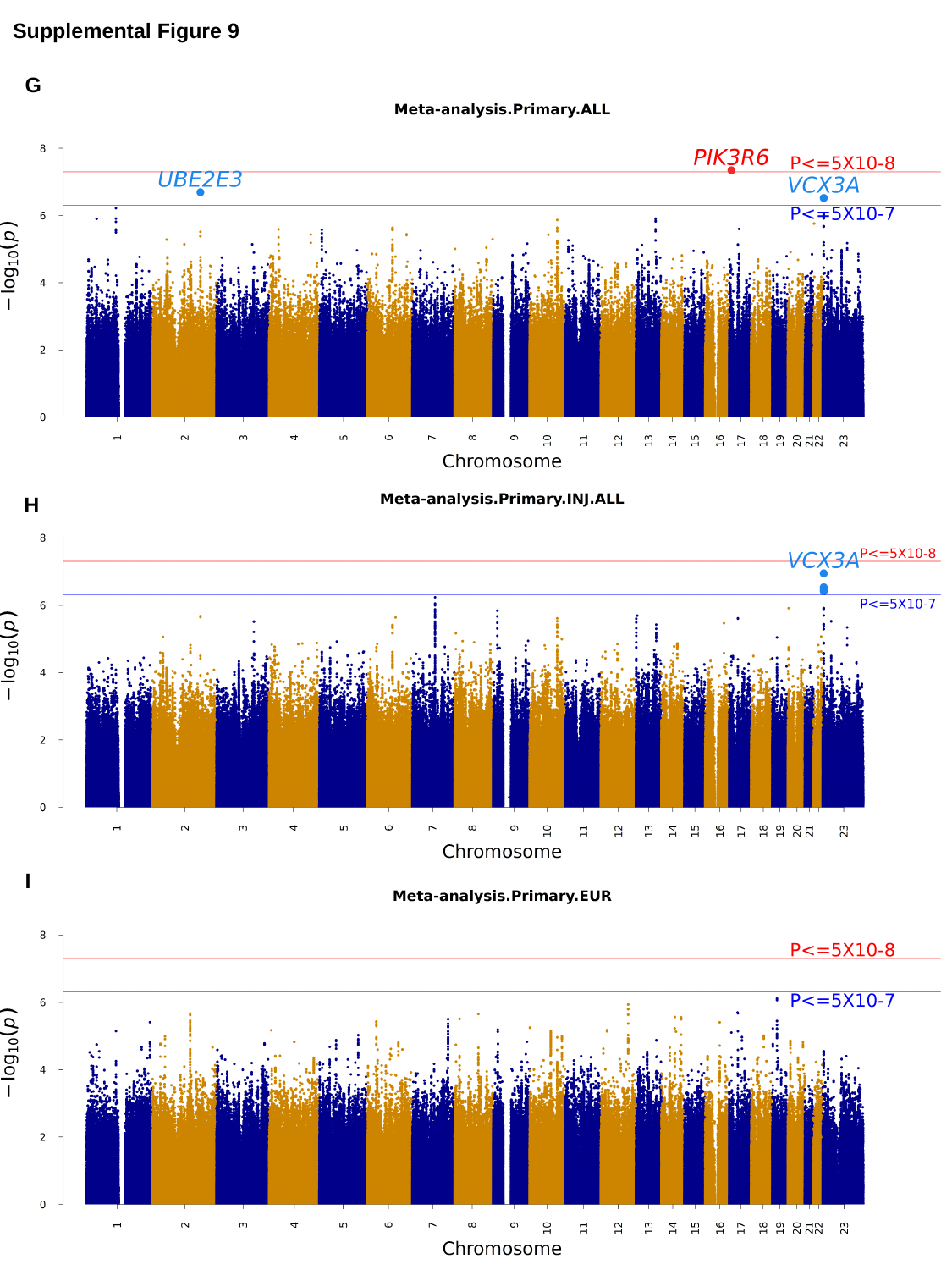

Supplemental Figure 9
G
H
I

### Slide 12
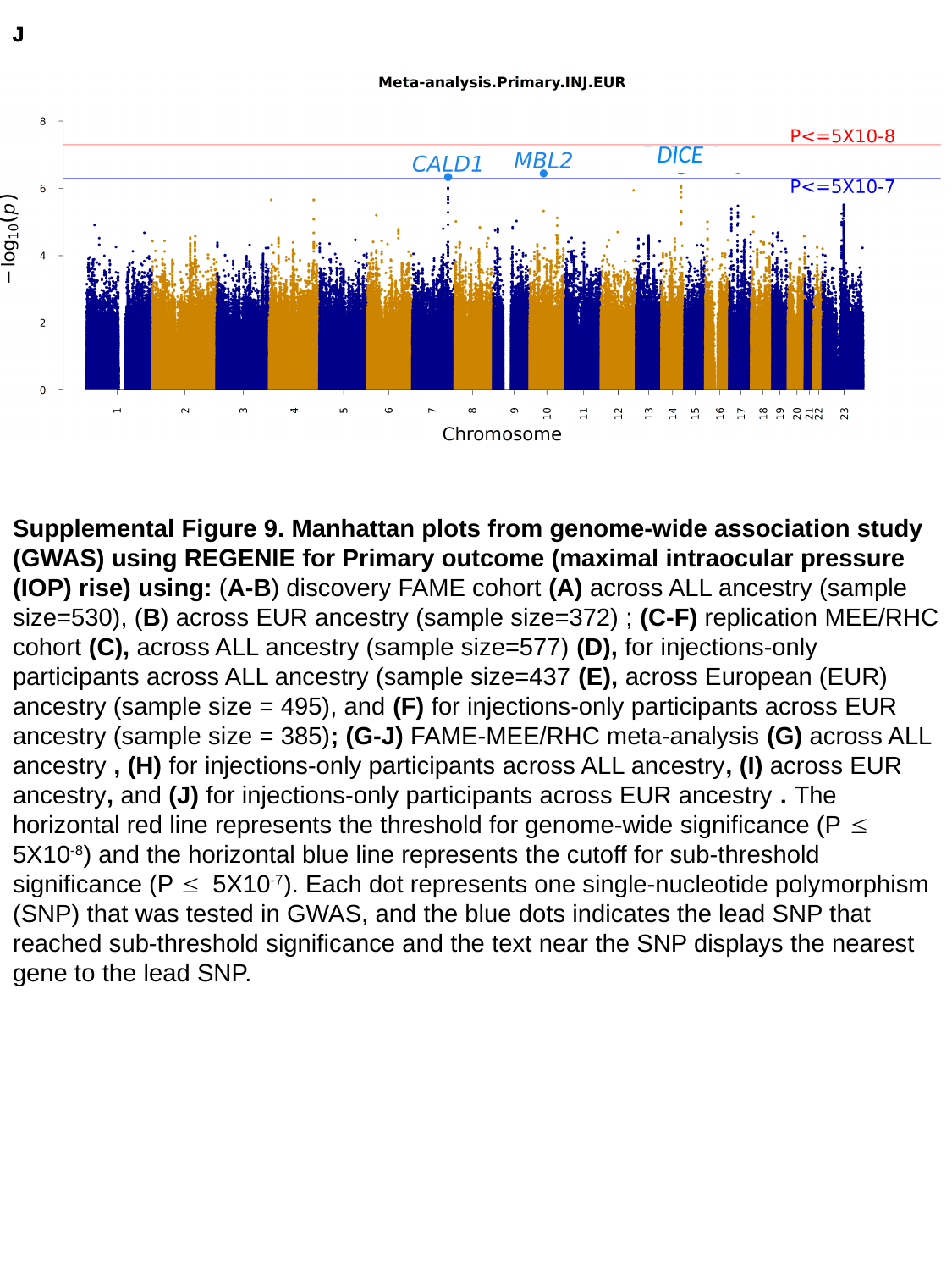

J
J
DICE
DICE
Supplemental Figure 9. Manhattan plots from genome-wide association study (GWAS) using REGENIE for Primary outcome (maximal intraocular pressure (IOP) rise) using: (A-B) discovery FAME cohort (A) across ALL ancestry (sample size=530), (B) across EUR ancestry (sample size=372) ; (C-F) replication MEE/RHC cohort (C), across ALL ancestry (sample size=577) (D), for injections-only participants across ALL ancestry (sample size=437 (E), across European (EUR) ancestry (sample size = 495), and (F) for injections-only participants across EUR ancestry (sample size = 385); (G-J) FAME-MEE/RHC meta-analysis (G) across ALL ancestry , (H) for injections-only participants across ALL ancestry, (I) across EUR ancestry, and (J) for injections-only participants across EUR ancestry . The horizontal red line represents the threshold for genome-wide significance (P ≤ 5X10^-8^) and the horizontal blue line represents the cutoff for sub-threshold significance (P ≤ 5X10^-7^). Each dot represents one single-nucleotide polymorphism (SNP) that was tested in GWAS, and the blue dots indicates the lead SNP that reached sub-threshold significance and the text near the SNP displays the nearest gene to the lead SNP.

### Slide 13
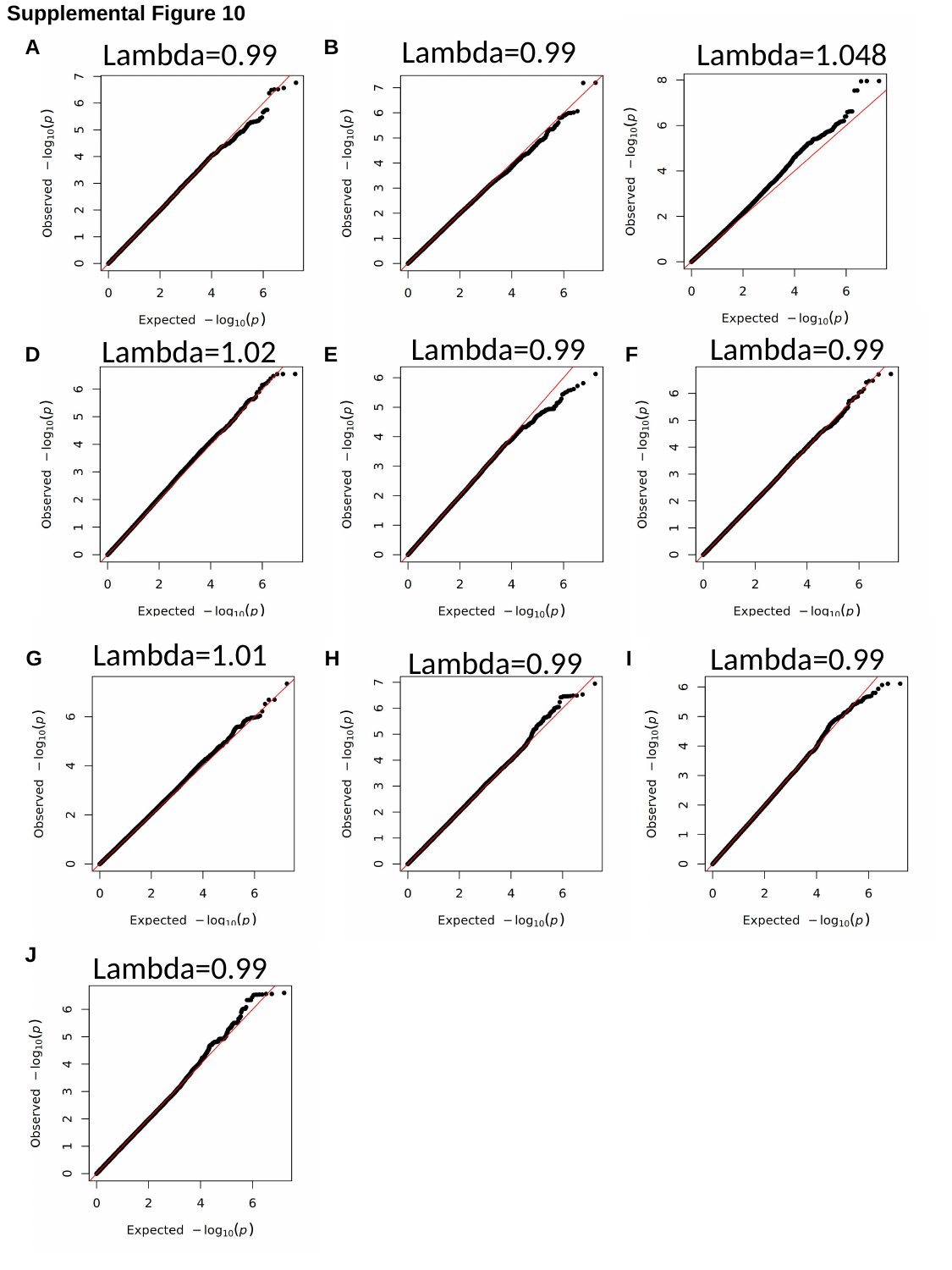

Supplemental Figure 10
Lambda=0.99
A
Lambda=0.99
B
C
Lambda=1.048
Lambda=0.99
Lambda=0.99
Lambda=1.02
D
E
F
Lambda=1.01
Lambda=0.99
Lambda=0.99
G
H
I
J
Lambda=0.99

### Slide 14
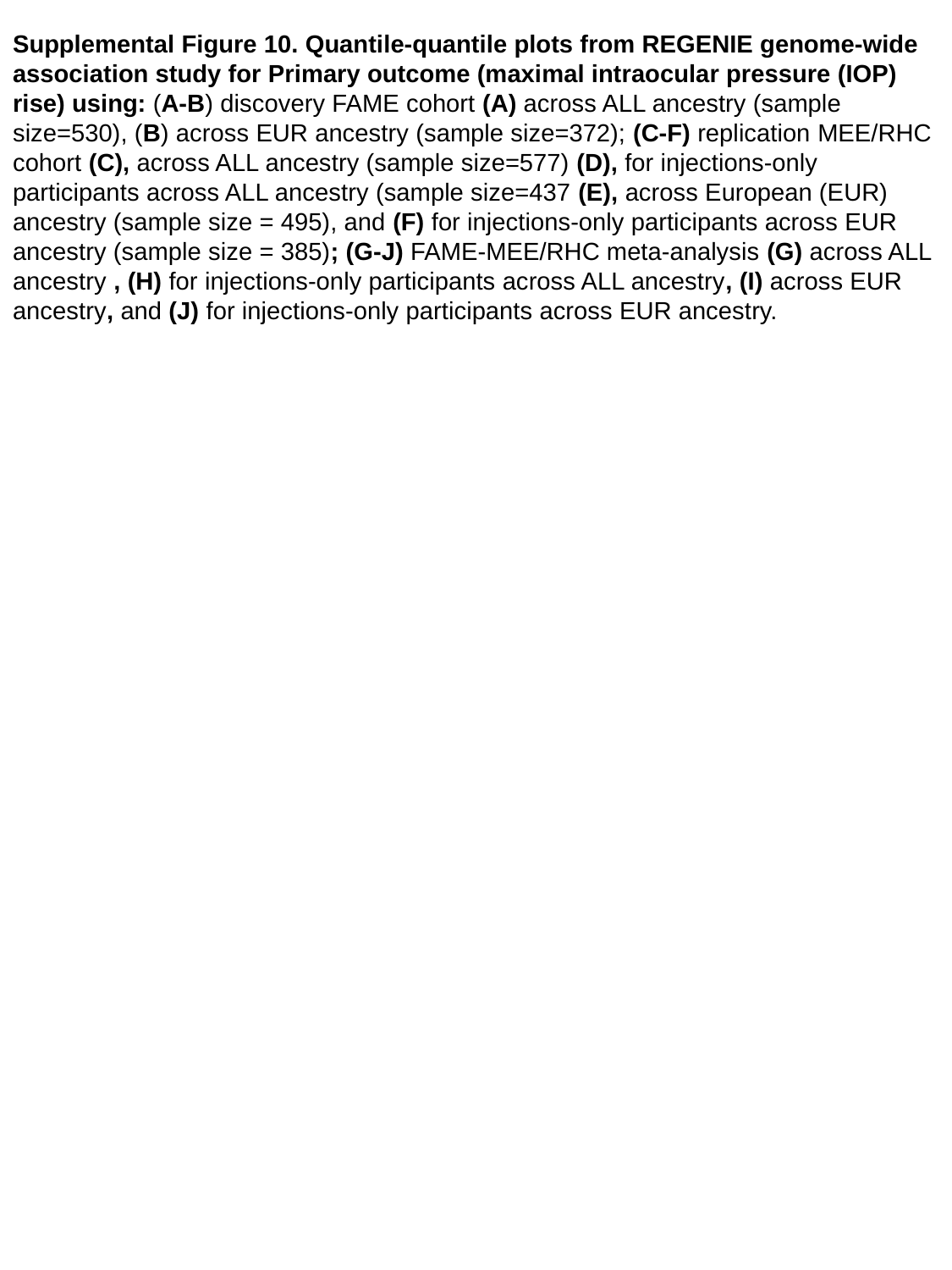

Supplemental Figure 10. Quantile-quantile plots from REGENIE genome-wide association study for Primary outcome (maximal intraocular pressure (IOP) rise) using: (A-B) discovery FAME cohort (A) across ALL ancestry (sample size=530), (B) across EUR ancestry (sample size=372); (C-F) replication MEE/RHC cohort (C), across ALL ancestry (sample size=577) (D), for injections-only participants across ALL ancestry (sample size=437 (E), across European (EUR) ancestry (sample size = 495), and (F) for injections-only participants across EUR ancestry (sample size = 385); (G-J) FAME-MEE/RHC meta-analysis (G) across ALL ancestry , (H) for injections-only participants across ALL ancestry, (I) across EUR ancestry, and (J) for injections-only participants across EUR ancestry.

### Slide 15
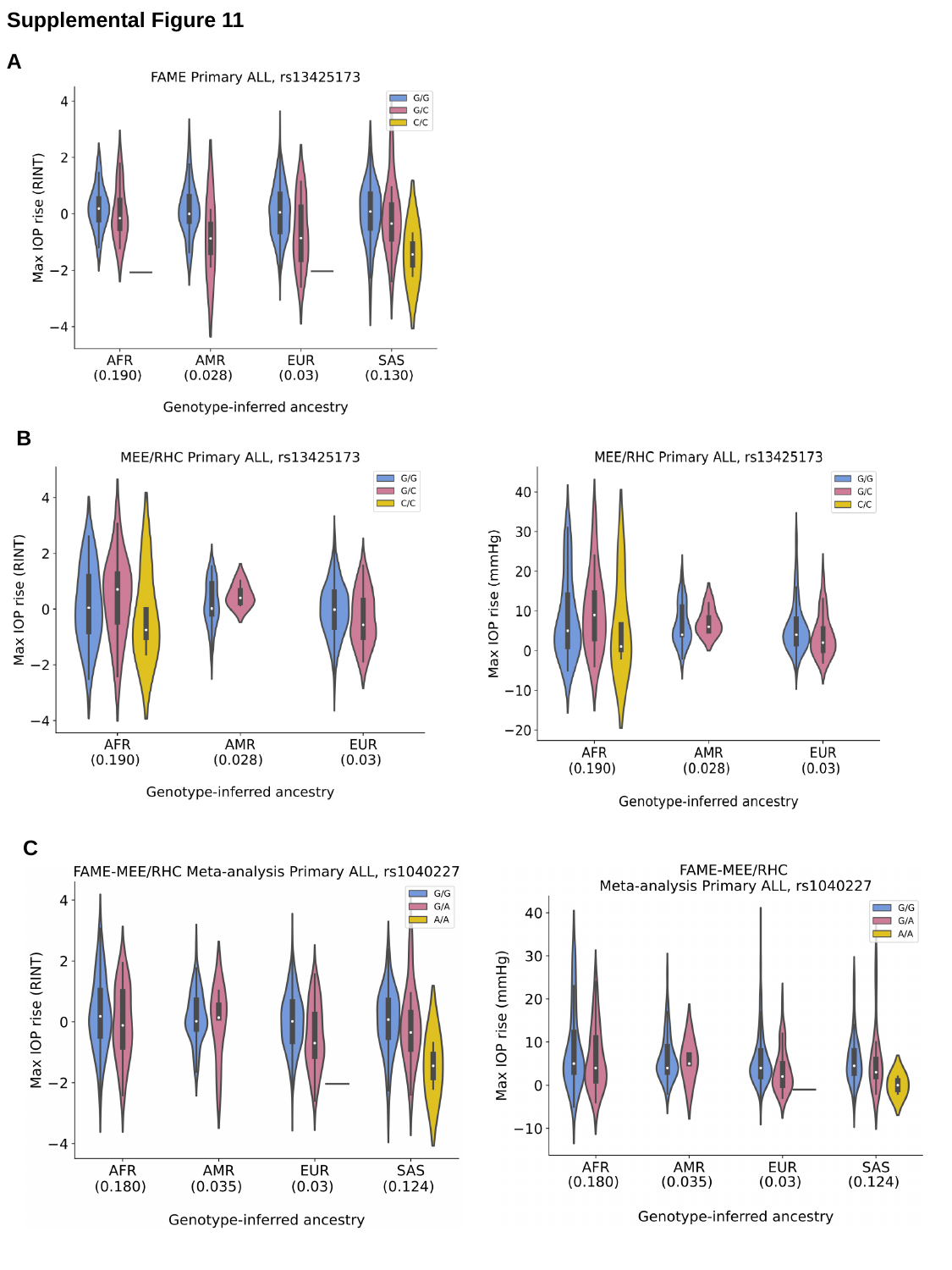

Supplemental Figure 11
A
B
C

### Slide 16
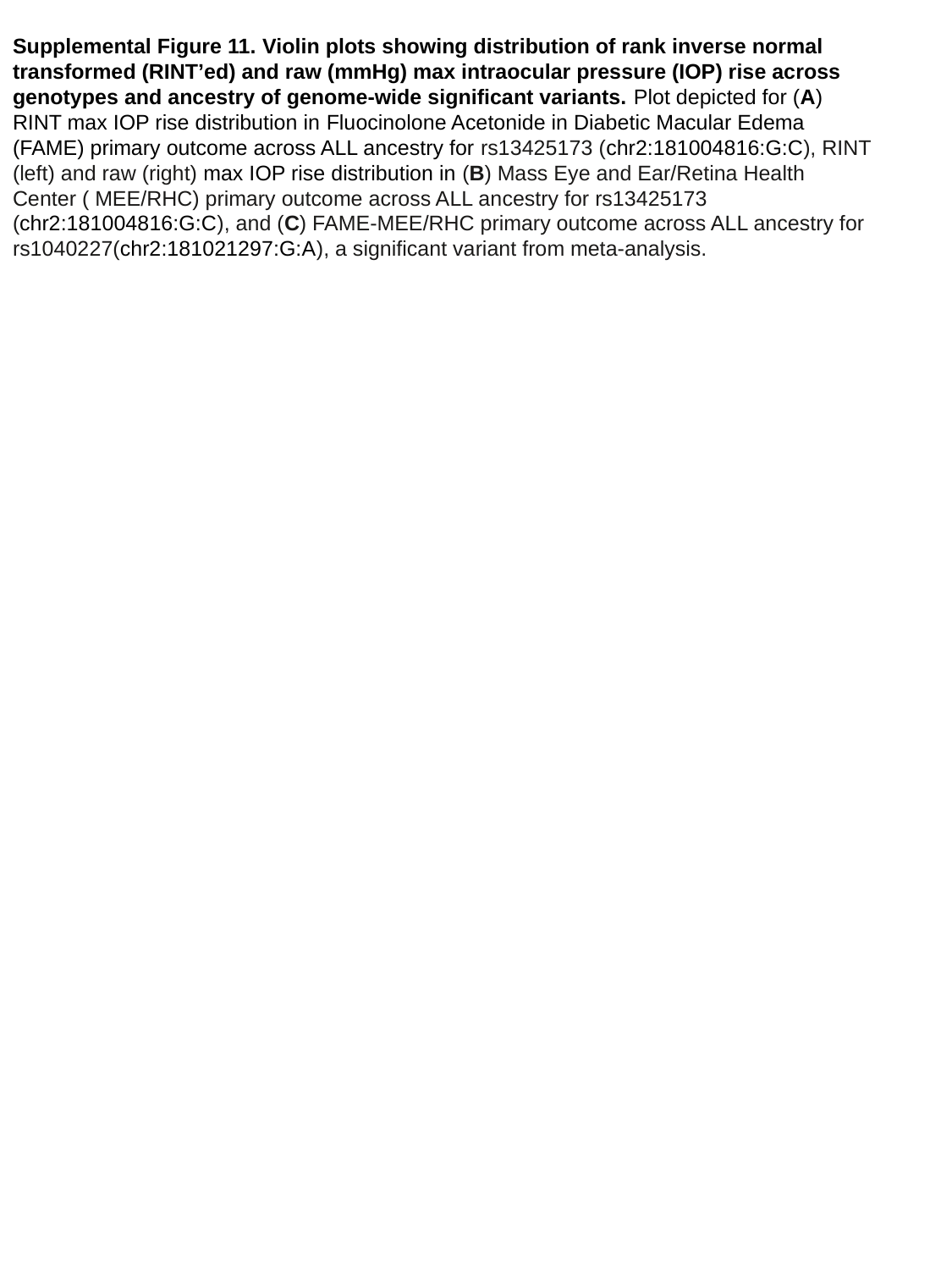

Supplemental Figure 11. Violin plots showing distribution of rank inverse normal transformed (RINT’ed) and raw (mmHg) max intraocular pressure (IOP) rise across genotypes and ancestry of genome-wide significant variants. Plot depicted for (A) RINT max IOP rise distribution in Fluocinolone Acetonide in Diabetic Macular Edema (FAME) primary outcome across ALL ancestry for rs13425173 (chr2:181004816:G:C), RINT (left) and raw (right) max IOP rise distribution in (B) Mass Eye and Ear/Retina Health Center ( MEE/RHC) primary outcome across ALL ancestry for rs13425173 (chr2:181004816:G:C), and (C) FAME-MEE/RHC primary outcome across ALL ancestry for rs1040227(chr2:181021297:G:A), a significant variant from meta-analysis.

### Slide 17
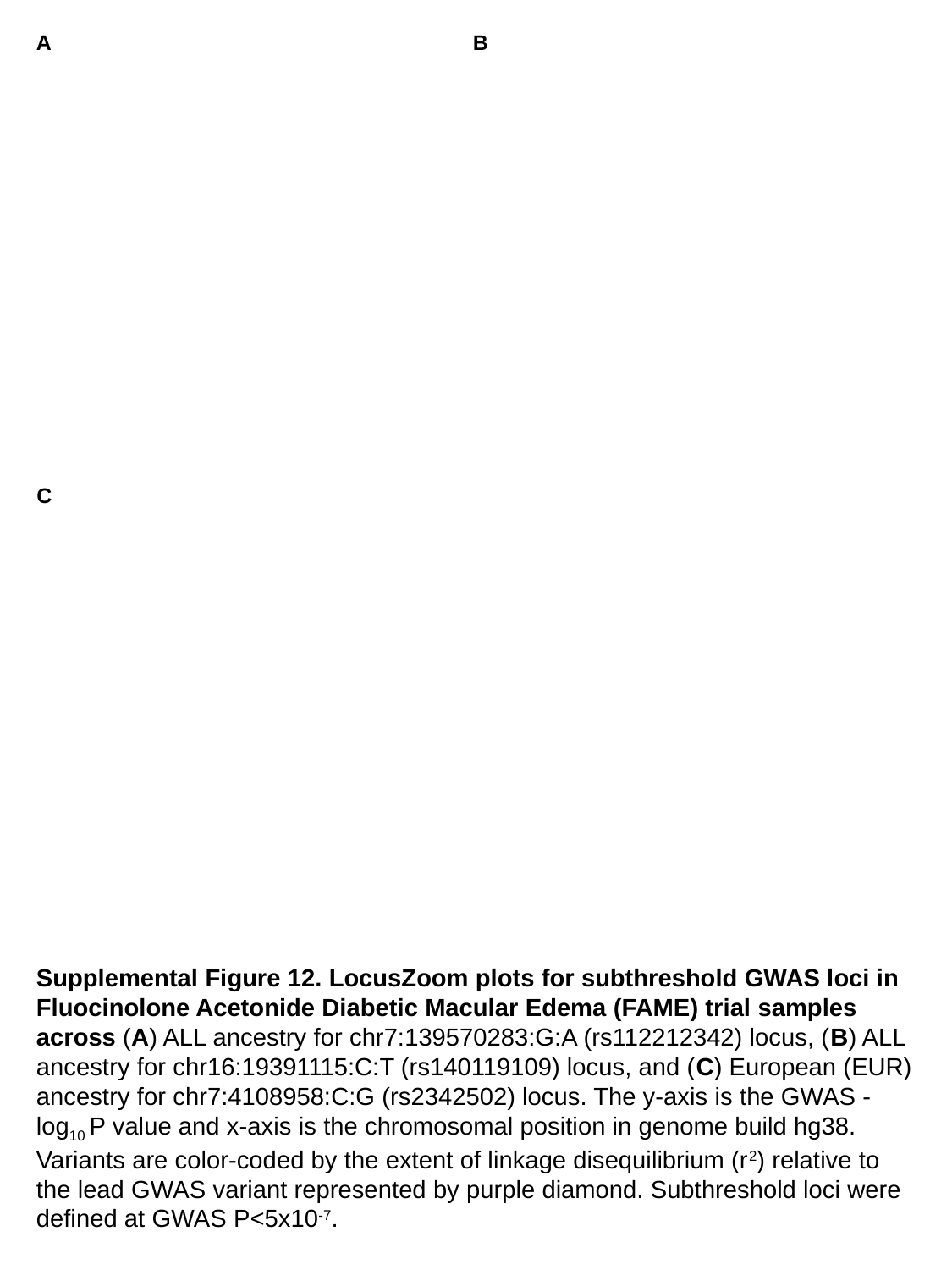

A
B
C
Supplemental Figure 12. LocusZoom plots for subthreshold GWAS loci in Fluocinolone Acetonide Diabetic Macular Edema (FAME) trial samples across (A) ALL ancestry for chr7:139570283:G:A (rs112212342) locus, (B) ALL ancestry for chr16:19391115:C:T (rs140119109) locus, and (C) European (EUR) ancestry for chr7:4108958:C:G (rs2342502) locus. The y-axis is the GWAS -log_10_ P value and x-axis is the chromosomal position in genome build hg38. Variants are color-coded by the extent of linkage disequilibrium (r^2^) relative to the lead GWAS variant represented by purple diamond. Subthreshold loci were defined at GWAS P<5x10^-7^.

### Slide 18
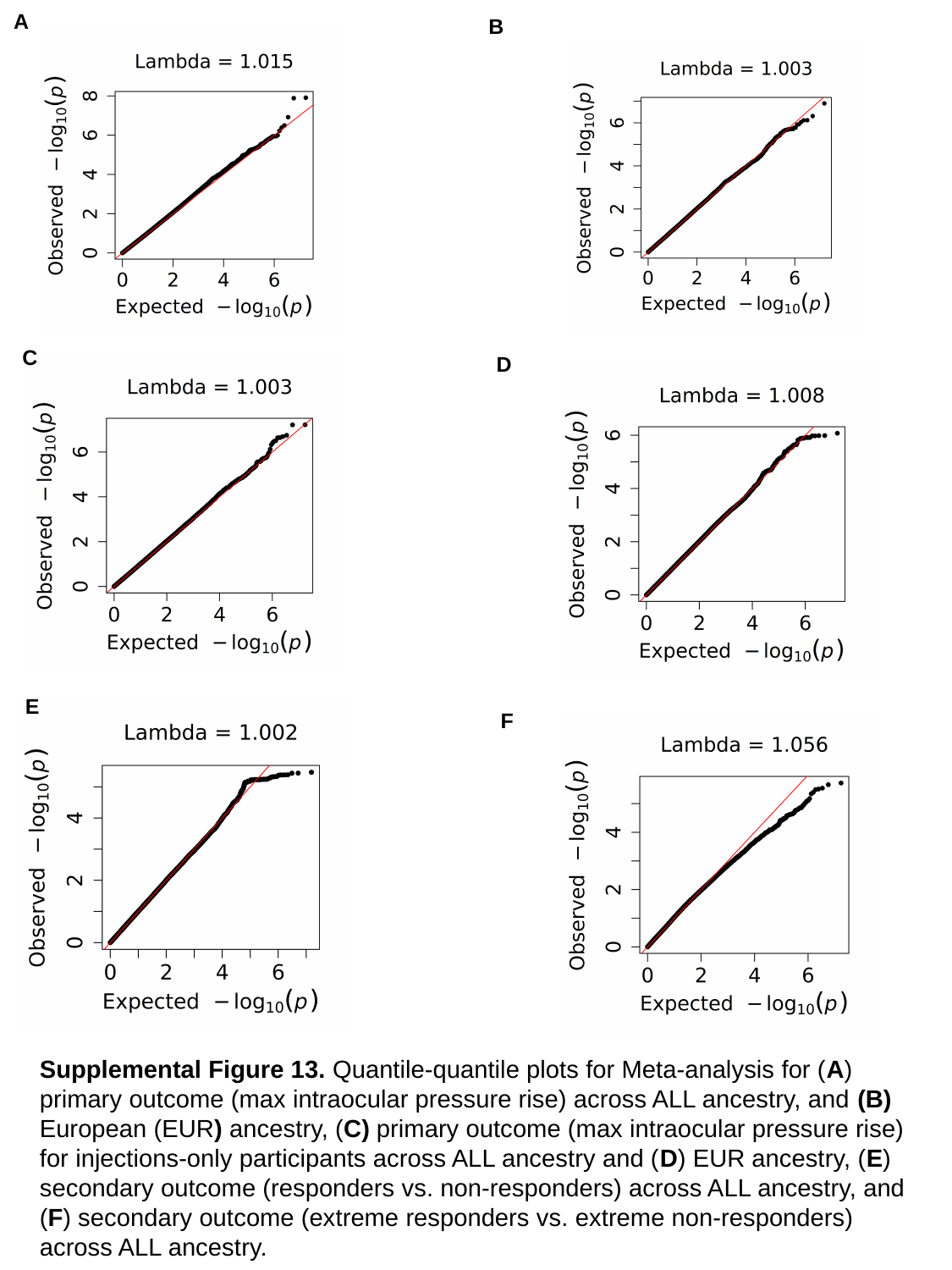

A
B
C
D
E
F
Supplemental Figure 13. Quantile-quantile plots for Meta-analysis for (A) primary outcome (max intraocular pressure rise) across ALL ancestry, and (B) European (EUR) ancestry, (C) primary outcome (max intraocular pressure rise) for injections-only participants across ALL ancestry and (D) EUR ancestry, (E) secondary outcome (responders vs. non-responders) across ALL ancestry, and (F) secondary outcome (extreme responders vs. extreme non-responders) across ALL ancestry.

### Slide 19
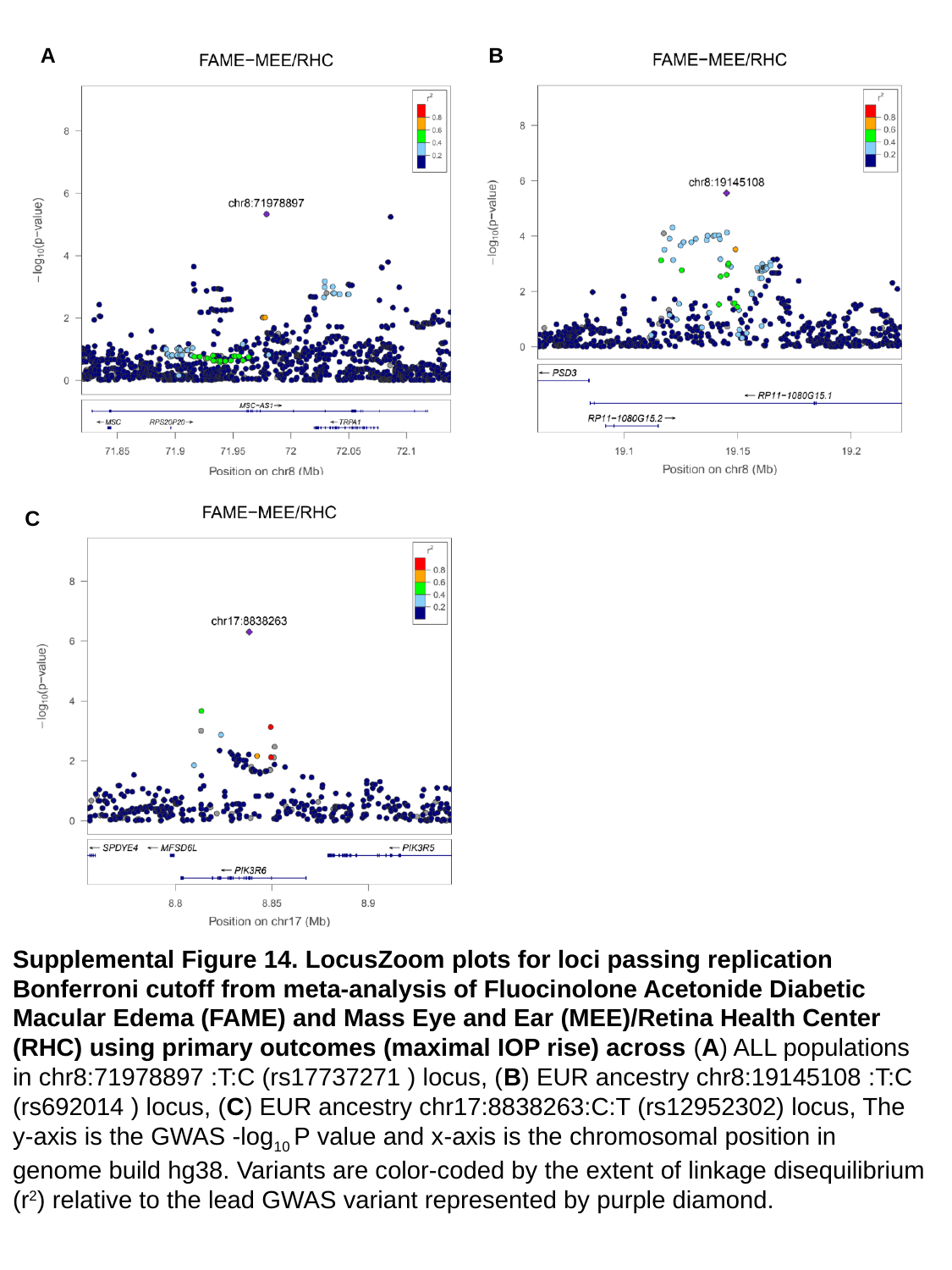

A
B
C
Supplemental Figure 14. LocusZoom plots for loci passing replication Bonferroni cutoff from meta-analysis of Fluocinolone Acetonide Diabetic Macular Edema (FAME) and Mass Eye and Ear (MEE)/Retina Health Center (RHC) using primary outcomes (maximal IOP rise) across (A) ALL populations in chr8:71978897 :T:C (rs17737271 ) locus, (B) EUR ancestry chr8:19145108 :T:C (rs692014 ) locus, (C) EUR ancestry chr17:8838263:C:T (rs12952302) locus, The y-axis is the GWAS -log_10_ P value and x-axis is the chromosomal position in genome build hg38. Variants are color-coded by the extent of linkage disequilibrium (r^2^) relative to the lead GWAS variant represented by purple diamond.

### Slide 20
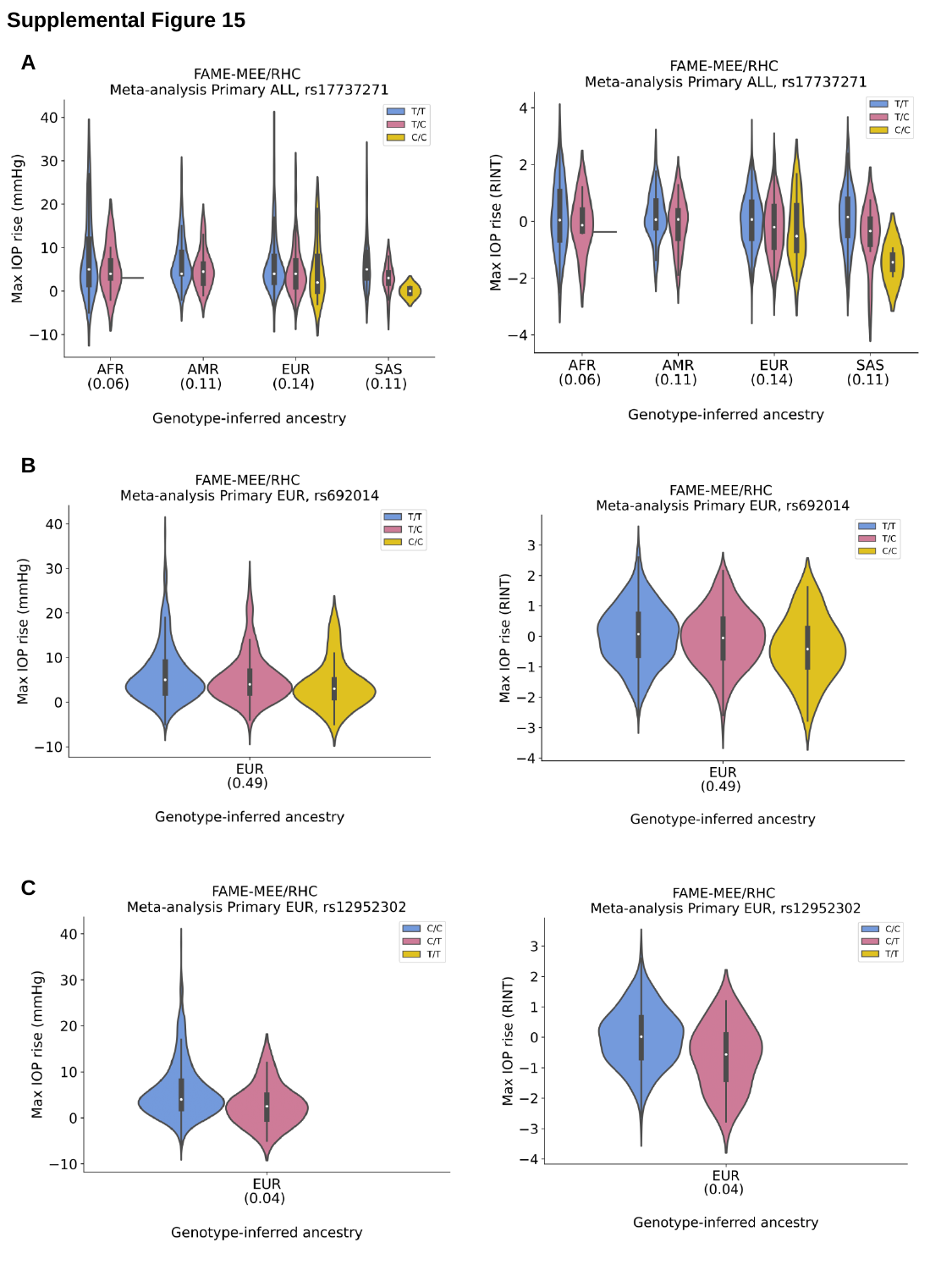

Supplemental Figure 15
A
B
C

### Slide 21
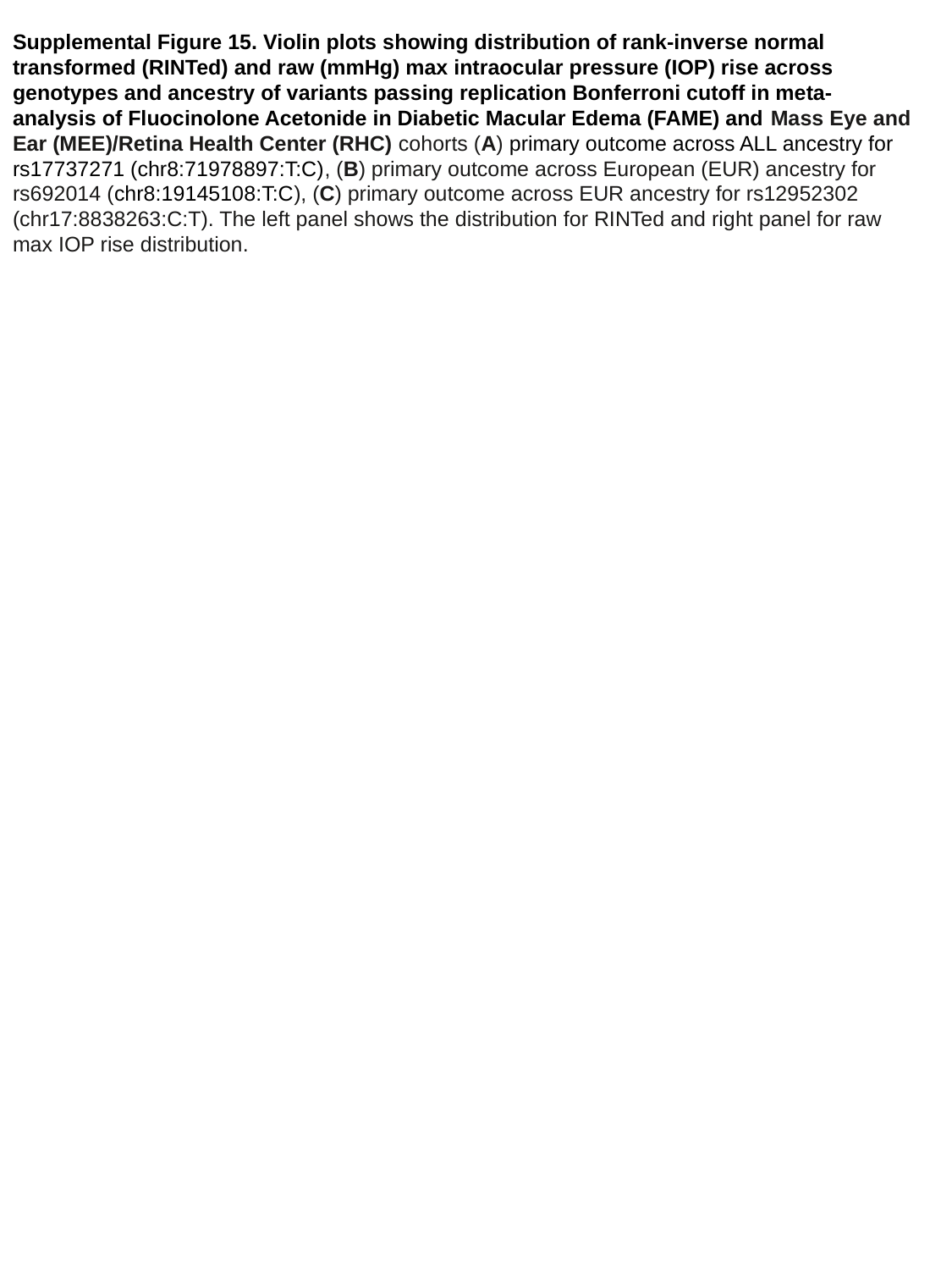

Supplemental Figure 15. Violin plots showing distribution of rank-inverse normal transformed (RINTed) and raw (mmHg) max intraocular pressure (IOP) rise across genotypes and ancestry of variants passing replication Bonferroni cutoff in meta-analysis of Fluocinolone Acetonide in Diabetic Macular Edema (FAME) and Mass Eye and Ear (MEE)/Retina Health Center (RHC) cohorts (A) primary outcome across ALL ancestry for rs17737271 (chr8:71978897:T:C), (B) primary outcome across European (EUR) ancestry for rs692014 (chr8:19145108:T:C), (C) primary outcome across EUR ancestry for rs12952302 (chr17:8838263:C:T). The left panel shows the distribution for RINTed and right panel for raw max IOP rise distribution.

### Slide 22
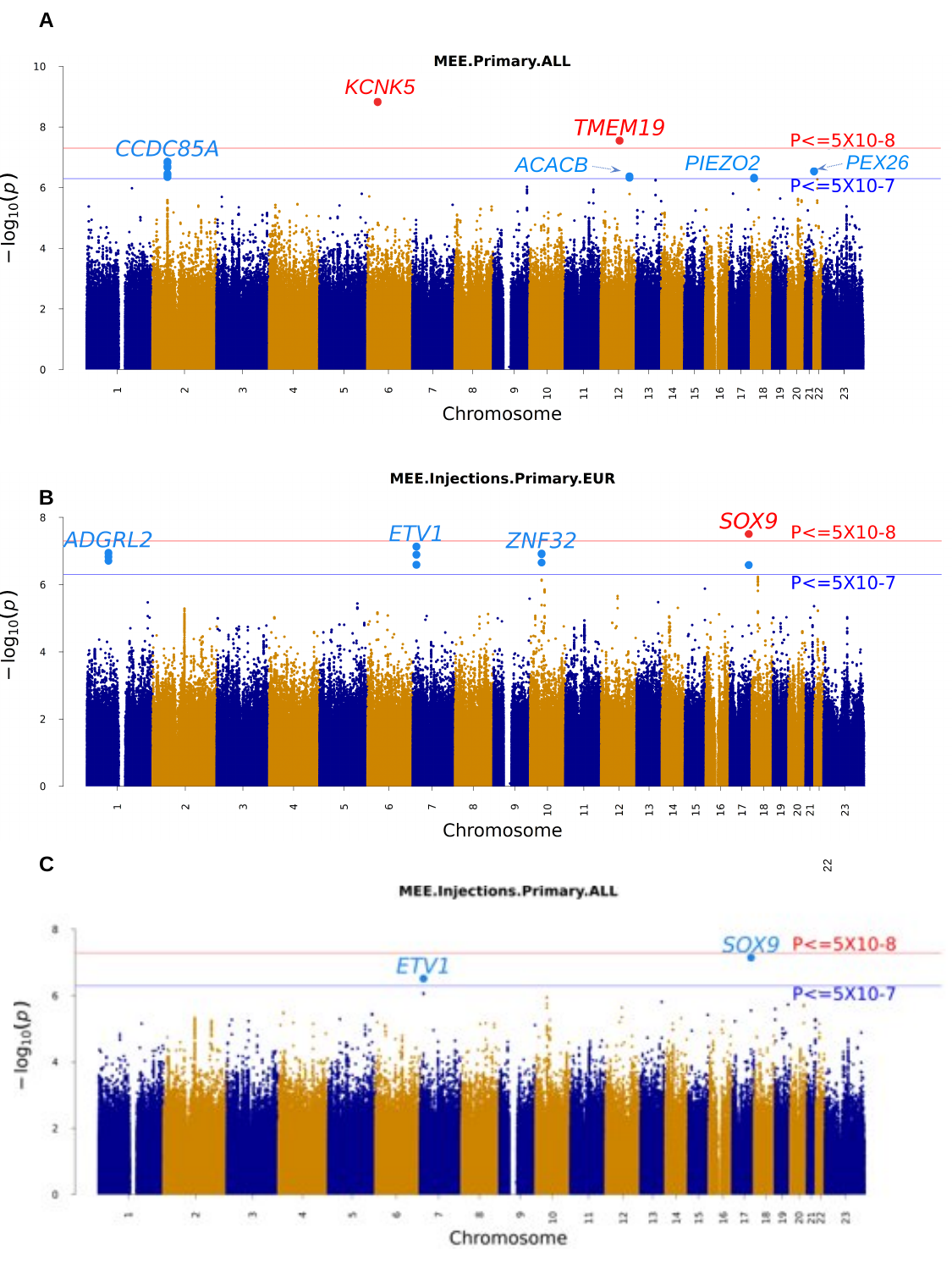

A
KCNK5
PIEZO2
PEX26
ACACB
B
C
22

### Slide 23
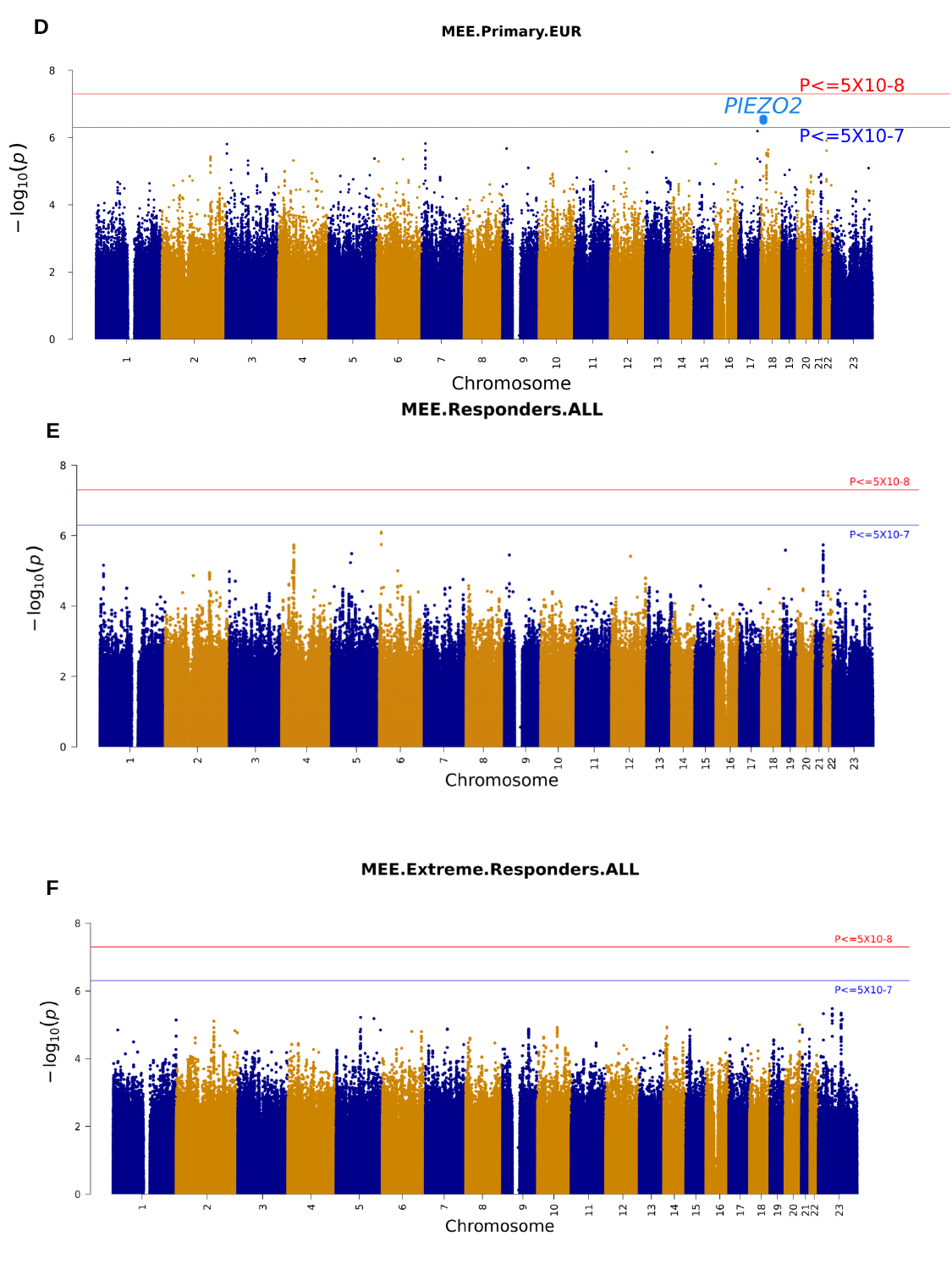

D
E
A
B
F
P<=5E-08
P<=5E-07

### Slide 24
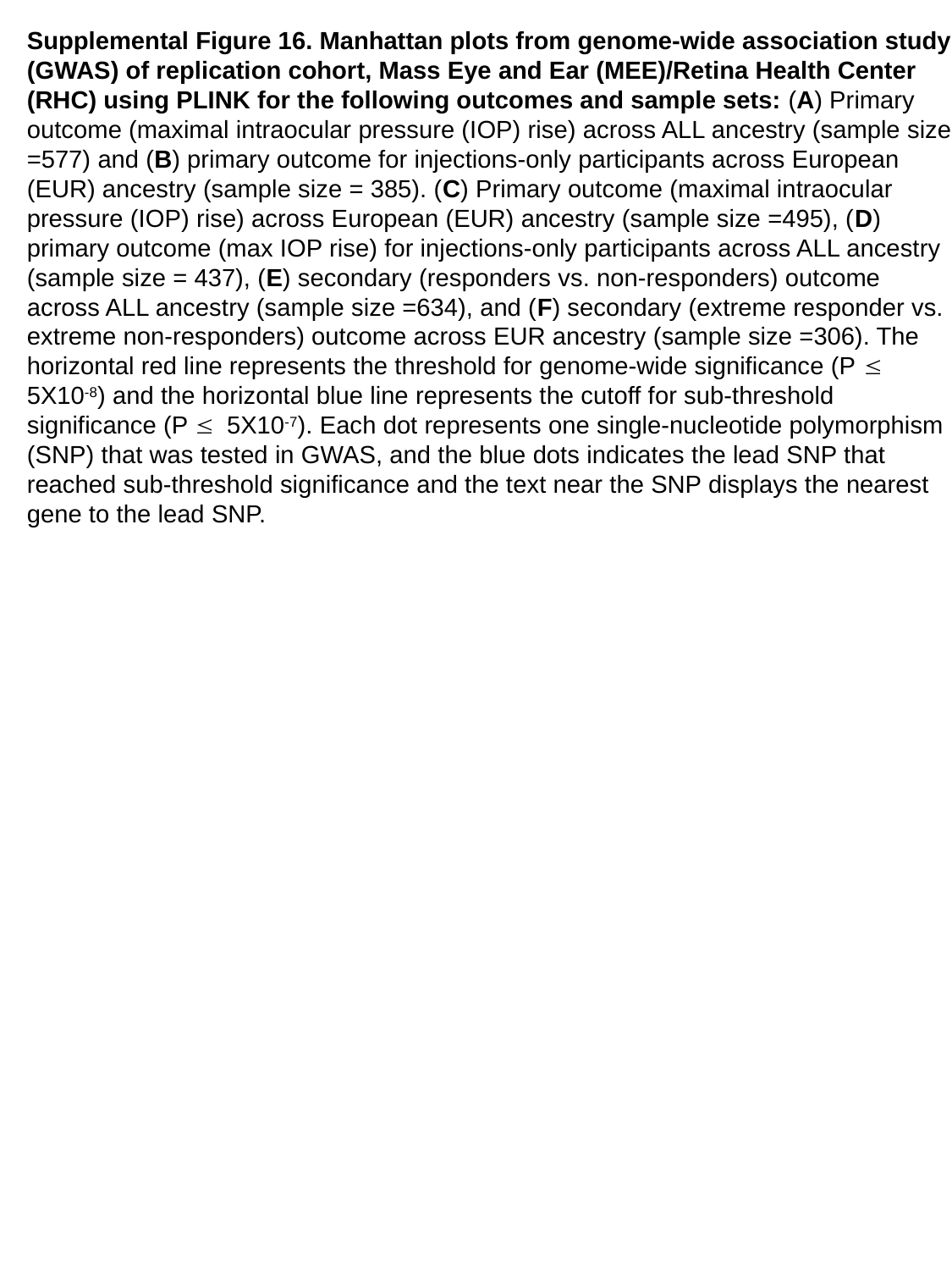

Supplemental Figure 16. Manhattan plots from genome-wide association study (GWAS) of replication cohort, Mass Eye and Ear (MEE)/Retina Health Center (RHC) using PLINK for the following outcomes and sample sets: (A) Primary outcome (maximal intraocular pressure (IOP) rise) across ALL ancestry (sample size =577) and (B) primary outcome for injections-only participants across European (EUR) ancestry (sample size = 385). (C) Primary outcome (maximal intraocular pressure (IOP) rise) across European (EUR) ancestry (sample size =495), (D) primary outcome (max IOP rise) for injections-only participants across ALL ancestry (sample size = 437), (E) secondary (responders vs. non-responders) outcome across ALL ancestry (sample size =634), and (F) secondary (extreme responder vs. extreme non-responders) outcome across EUR ancestry (sample size =306). The horizontal red line represents the threshold for genome-wide significance (P ≤ 5X10^-8^) and the horizontal blue line represents the cutoff for sub-threshold significance (P ≤ 5X10^-7^). Each dot represents one single-nucleotide polymorphism (SNP) that was tested in GWAS, and the blue dots indicates the lead SNP that reached sub-threshold significance and the text near the SNP displays the nearest gene to the lead SNP.

### Slide 25
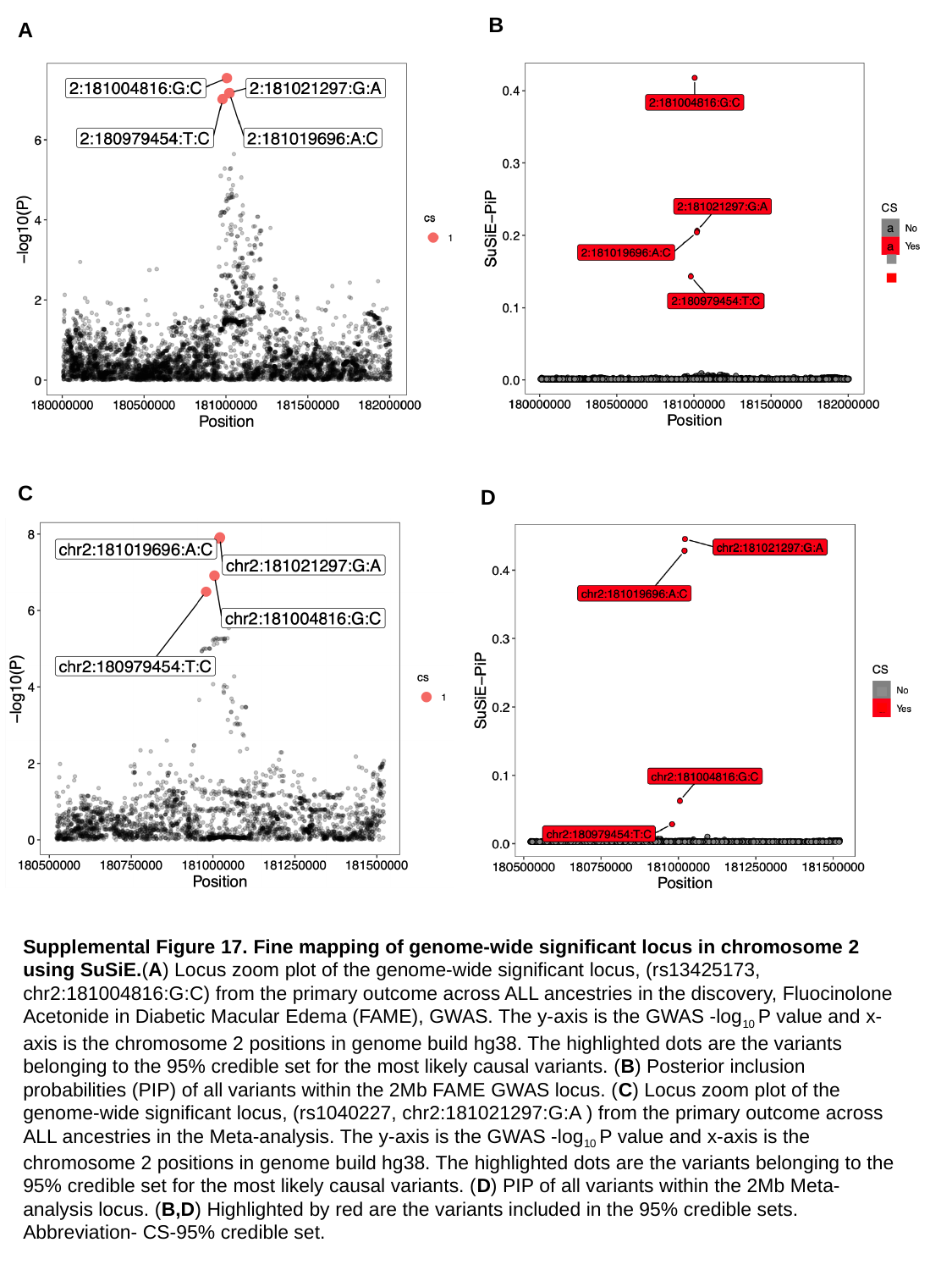

B
A
C
D
Supplemental Figure 17. Fine mapping of genome-wide significant locus in chromosome 2 using SuSiE.(A) Locus zoom plot of the genome-wide significant locus, (rs13425173, chr2:181004816:G:C) from the primary outcome across ALL ancestries in the discovery, Fluocinolone Acetonide in Diabetic Macular Edema (FAME), GWAS. The y-axis is the GWAS -log_10_ P value and x-axis is the chromosome 2 positions in genome build hg38. The highlighted dots are the variants belonging to the 95% credible set for the most likely causal variants. (B) Posterior inclusion probabilities (PIP) of all variants within the 2Mb FAME GWAS locus. (C) Locus zoom plot of the genome-wide significant locus, (rs1040227, chr2:181021297:G:A ) from the primary outcome across ALL ancestries in the Meta-analysis. The y-axis is the GWAS -log_10_ P value and x-axis is the chromosome 2 positions in genome build hg38. The highlighted dots are the variants belonging to the 95% credible set for the most likely causal variants. (D) PIP of all variants within the 2Mb Meta-analysis locus. (B,D) Highlighted by red are the variants included in the 95% credible sets.
Abbreviation- CS-95% credible set.

### Slide 26
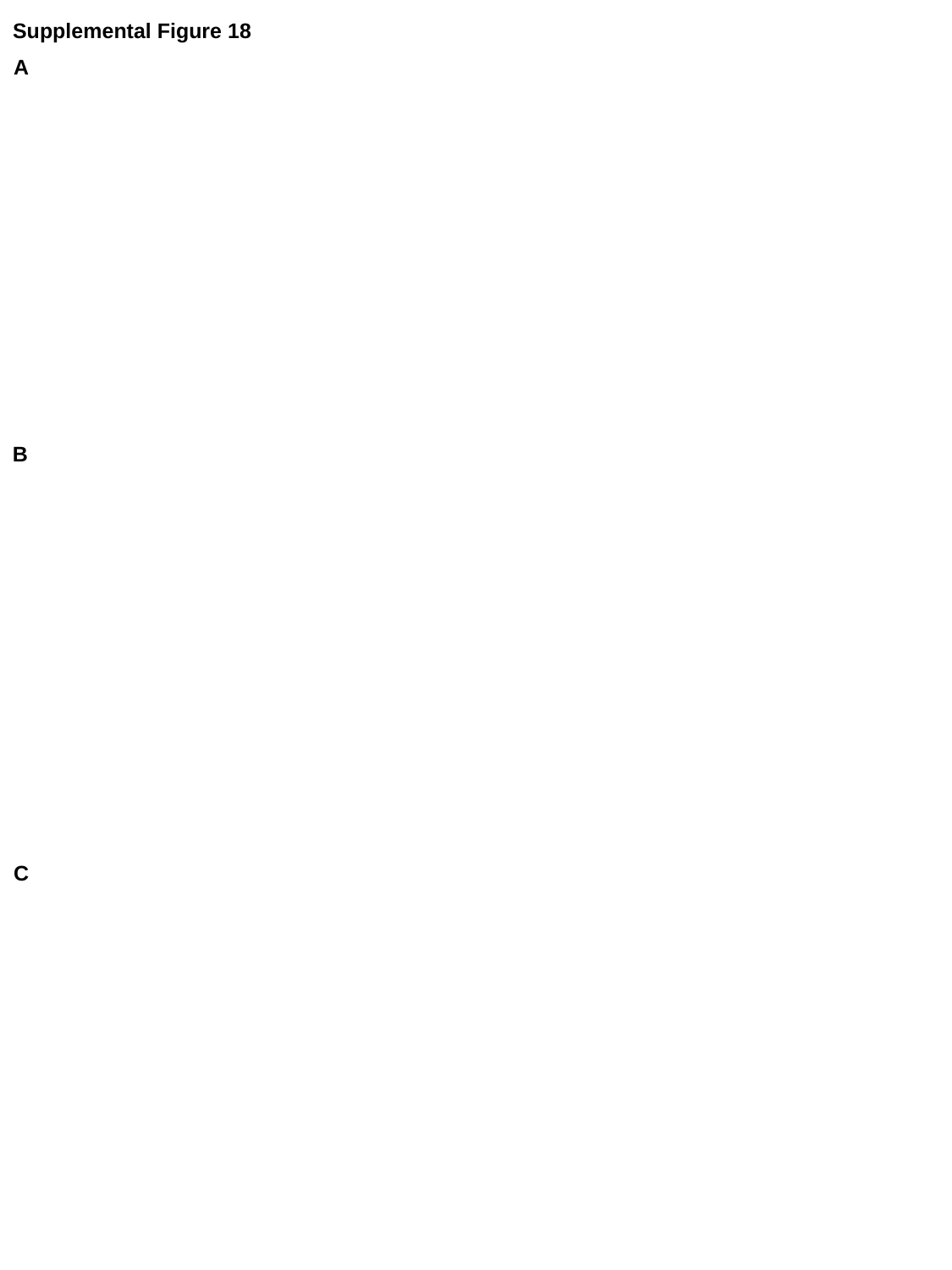

Supplemental Figure 18
A
B
C

### Slide 27
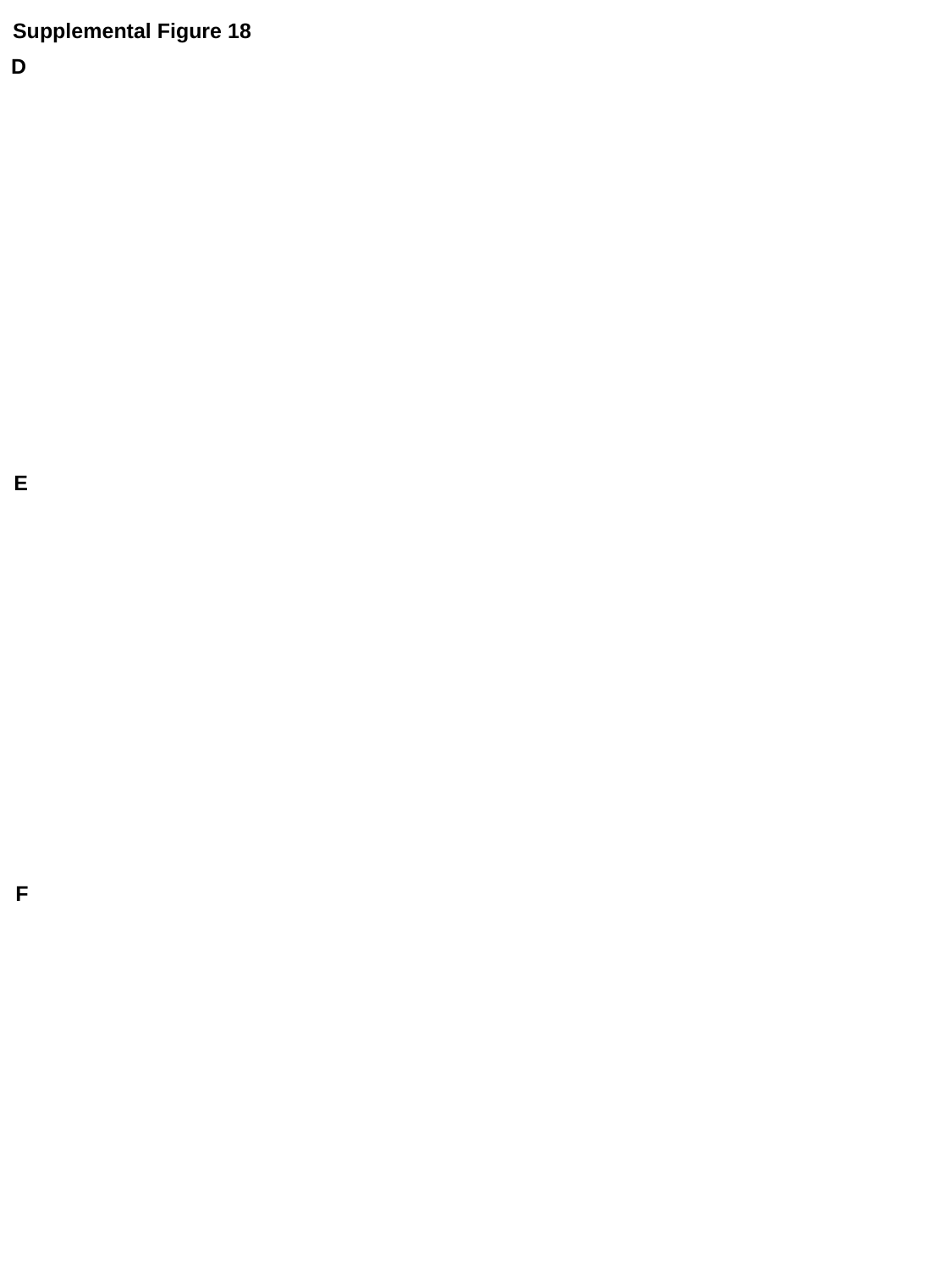

Supplemental Figure 18
D
E
F

### Slide 28
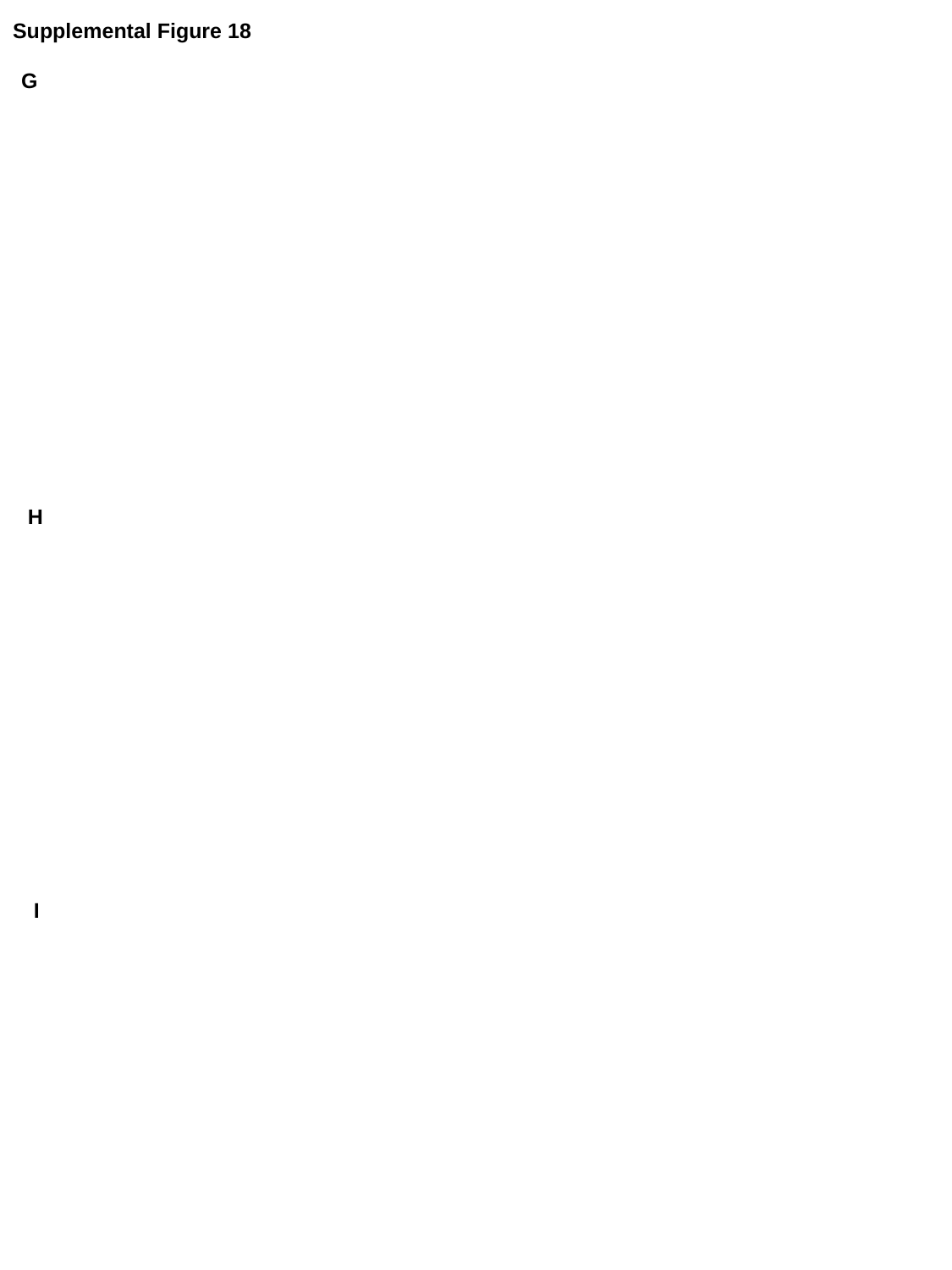

Supplemental Figure 18
G
H
I

### Slide 29
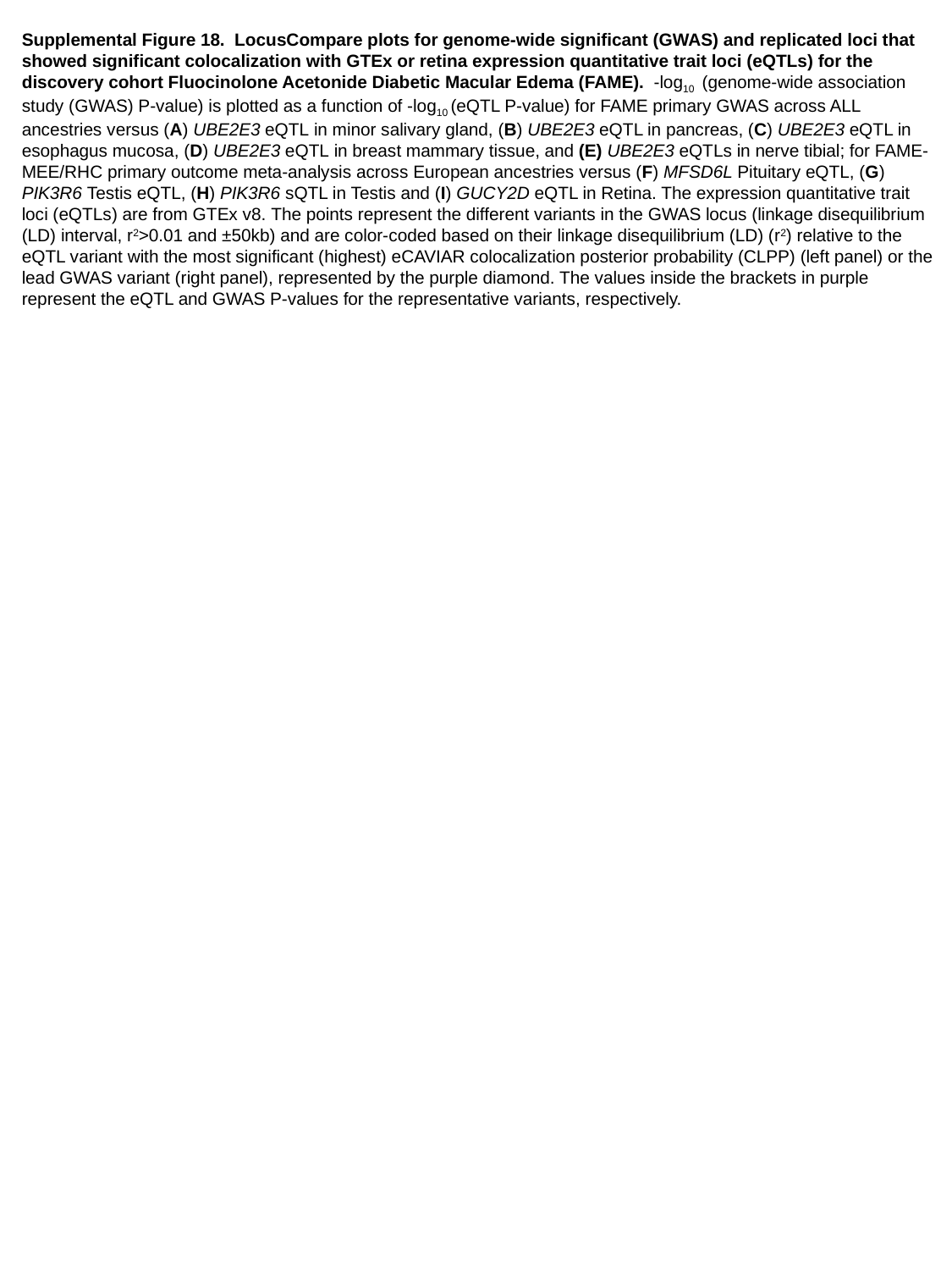

Supplemental Figure 18.  LocusCompare plots for genome-wide significant (GWAS) and replicated loci that showed significant colocalization with GTEx or retina expression quantitative trait loci (eQTLs) for the discovery cohort Fluocinolone Acetonide Diabetic Macular Edema (FAME).  -log_10_  (genome-wide association study (GWAS) P-value) is plotted as a function of -log_10_ (eQTL P-value) for FAME primary GWAS across ALL ancestries versus (A) UBE2E3 eQTL in minor salivary gland, (B) UBE2E3 eQTL in pancreas, (C) UBE2E3 eQTL in esophagus mucosa, (D) UBE2E3 eQTL in breast mammary tissue, and (E) UBE2E3 eQTLs in nerve tibial; for FAME-MEE/RHC primary outcome meta-analysis across European ancestries versus (F) MFSD6L Pituitary eQTL, (G) PIK3R6 Testis eQTL, (H) PIK3R6 sQTL in Testis and (I) GUCY2D eQTL in Retina. The expression quantitative trait loci (eQTLs) are from GTEx v8. The points represent the different variants in the GWAS locus (linkage disequilibrium (LD) interval, r^2^>0.01 and ±50kb) and are color-coded based on their linkage disequilibrium (LD) (r^2^) relative to the eQTL variant with the most significant (highest) eCAVIAR colocalization posterior probability (CLPP) (left panel) or the lead GWAS variant (right panel), represented by the purple diamond. The values inside the brackets in purple represent the eQTL and GWAS P-values for the representative variants, respectively.

### Slide 30
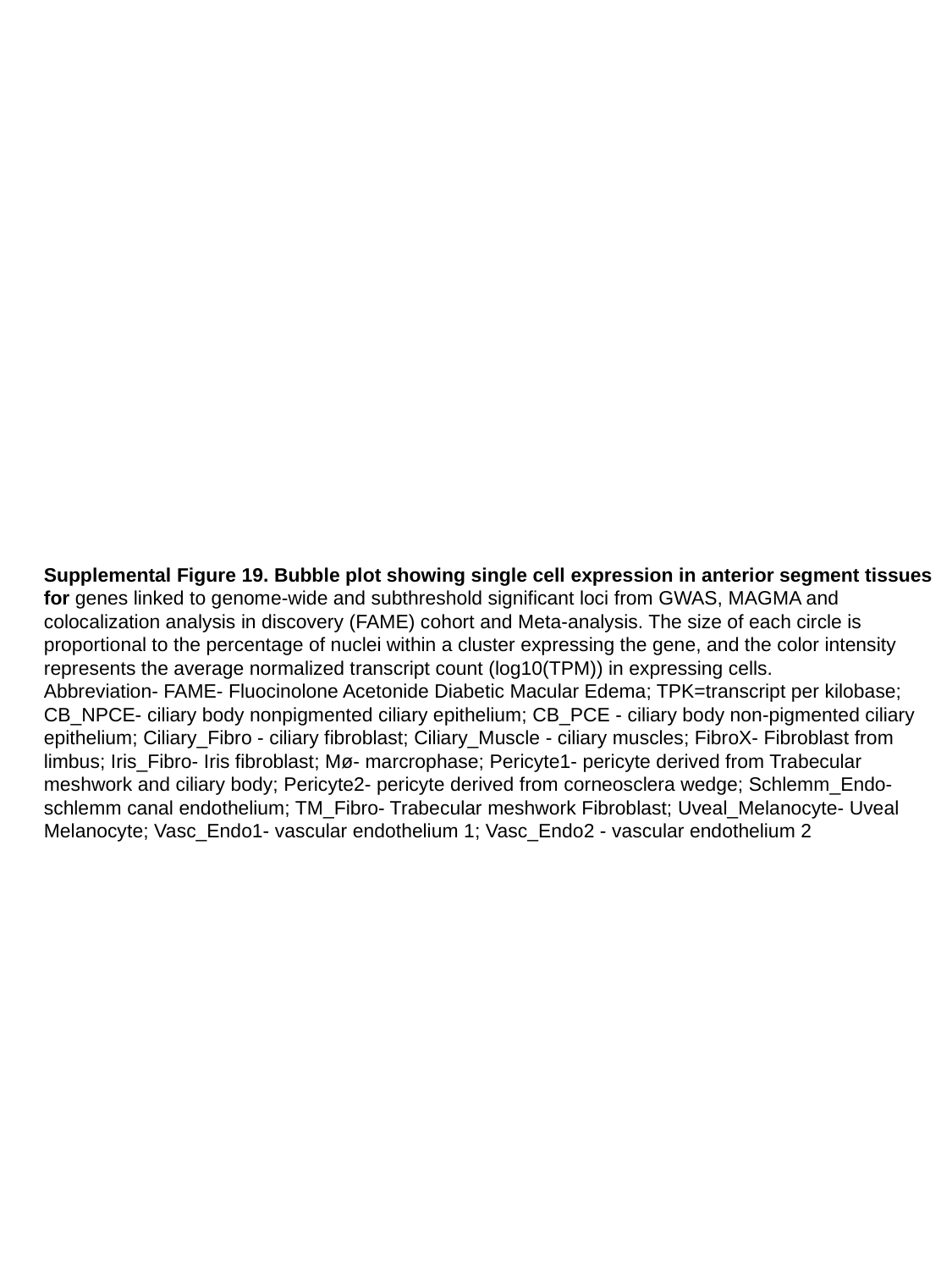

Supplemental Figure 19. Bubble plot showing single cell expression in anterior segment tissues for genes linked to genome-wide and subthreshold significant loci from GWAS, MAGMA and colocalization analysis in discovery (FAME) cohort and Meta-analysis. The size of each circle is proportional to the percentage of nuclei within a cluster expressing the gene, and the color intensity represents the average normalized transcript count (log10(TPM)) in expressing cells.
Abbreviation- FAME- Fluocinolone Acetonide Diabetic Macular Edema; TPK=transcript per kilobase; CB_NPCE- ciliary body nonpigmented ciliary epithelium; CB_PCE - ciliary body non-pigmented ciliary epithelium; Ciliary_Fibro - ciliary fibroblast; Ciliary_Muscle - ciliary muscles; FibroX- Fibroblast from limbus; Iris_Fibro- Iris fibroblast; Mø- marcrophase; Pericyte1- pericyte derived from Trabecular meshwork and ciliary body; Pericyte2- pericyte derived from corneosclera wedge; Schlemm_Endo- schlemm canal endothelium; TM_Fibro- Trabecular meshwork Fibroblast; Uveal_Melanocyte- Uveal Melanocyte; Vasc_Endo1- vascular endothelium 1; Vasc_Endo2 - vascular endothelium 2

### Slide 31
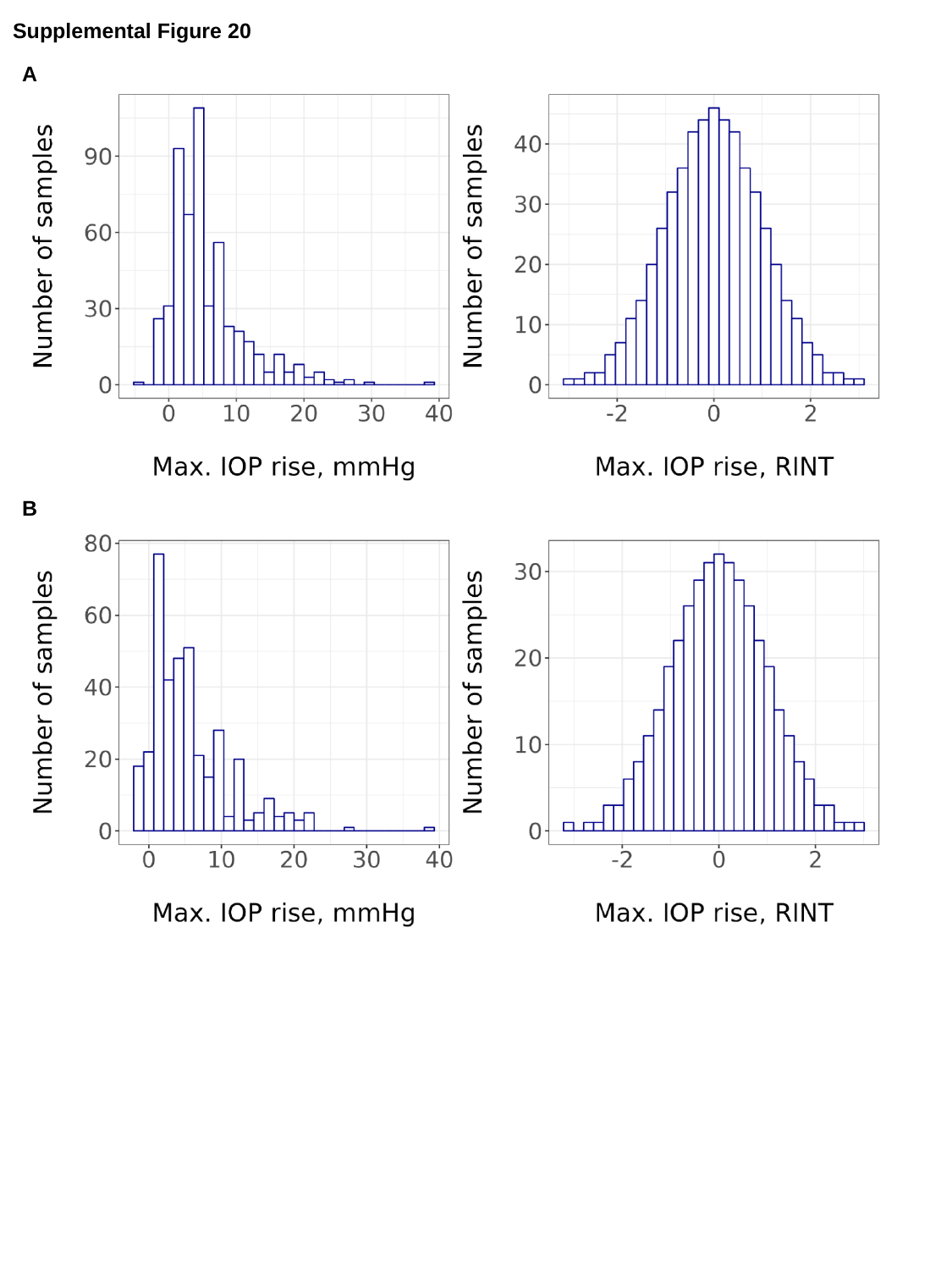

Supplemental Figure 20
A
B

### Slide 32
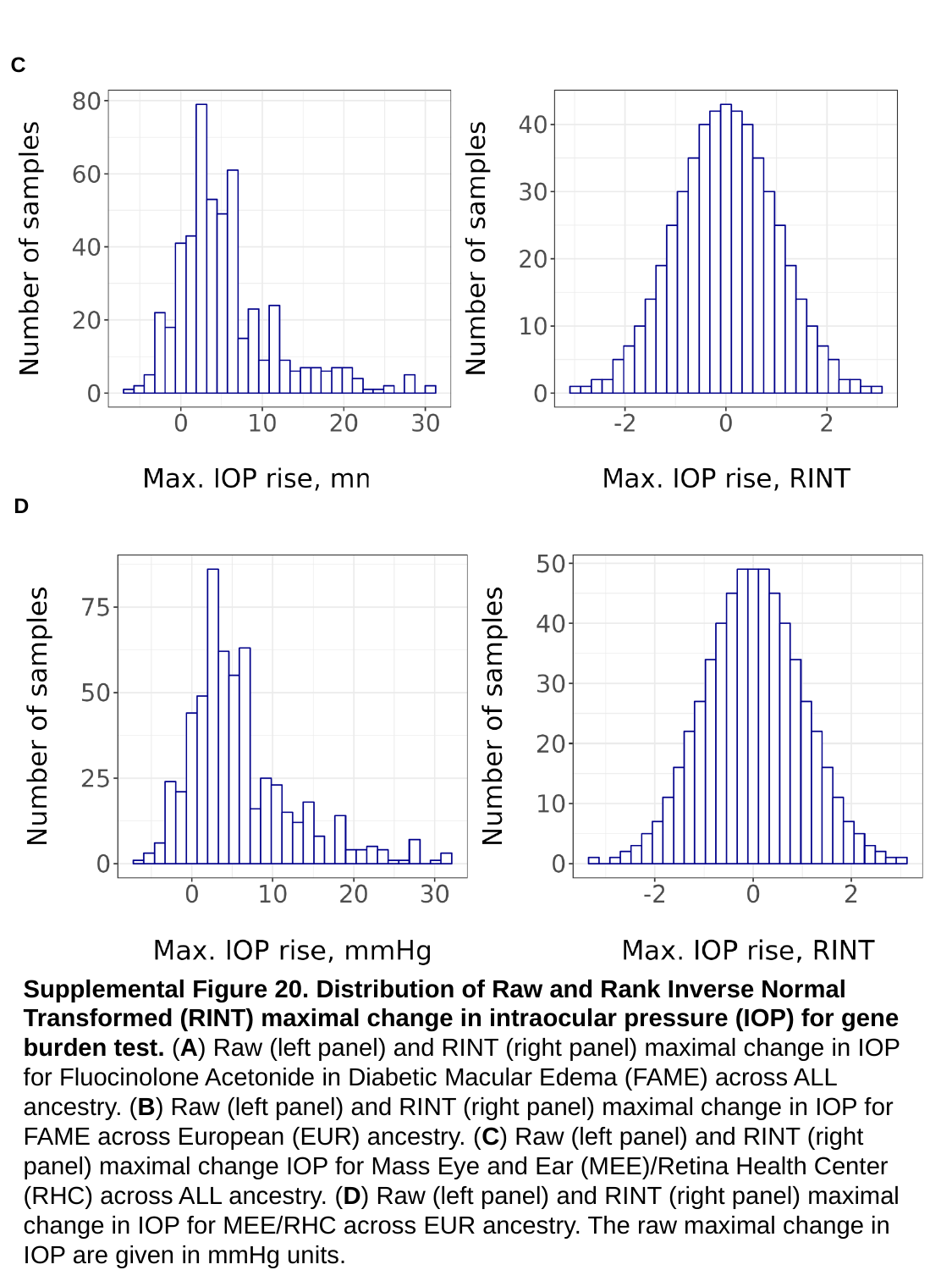

C
D
Supplemental Figure 20. Distribution of Raw and Rank Inverse Normal Transformed (RINT) maximal change in intraocular pressure (IOP) for gene burden test. (A) Raw (left panel) and RINT (right panel) maximal change in IOP for Fluocinolone Acetonide in Diabetic Macular Edema (FAME) across ALL ancestry. (B) Raw (left panel) and RINT (right panel) maximal change in IOP for FAME across European (EUR) ancestry. (C) Raw (left panel) and RINT (right panel) maximal change IOP for Mass Eye and Ear (MEE)/Retina Health Center (RHC) across ALL ancestry. (D) Raw (left panel) and RINT (right panel) maximal change in IOP for MEE/RHC across EUR ancestry. The raw maximal change in IOP are given in mmHg units.

### Slide 33
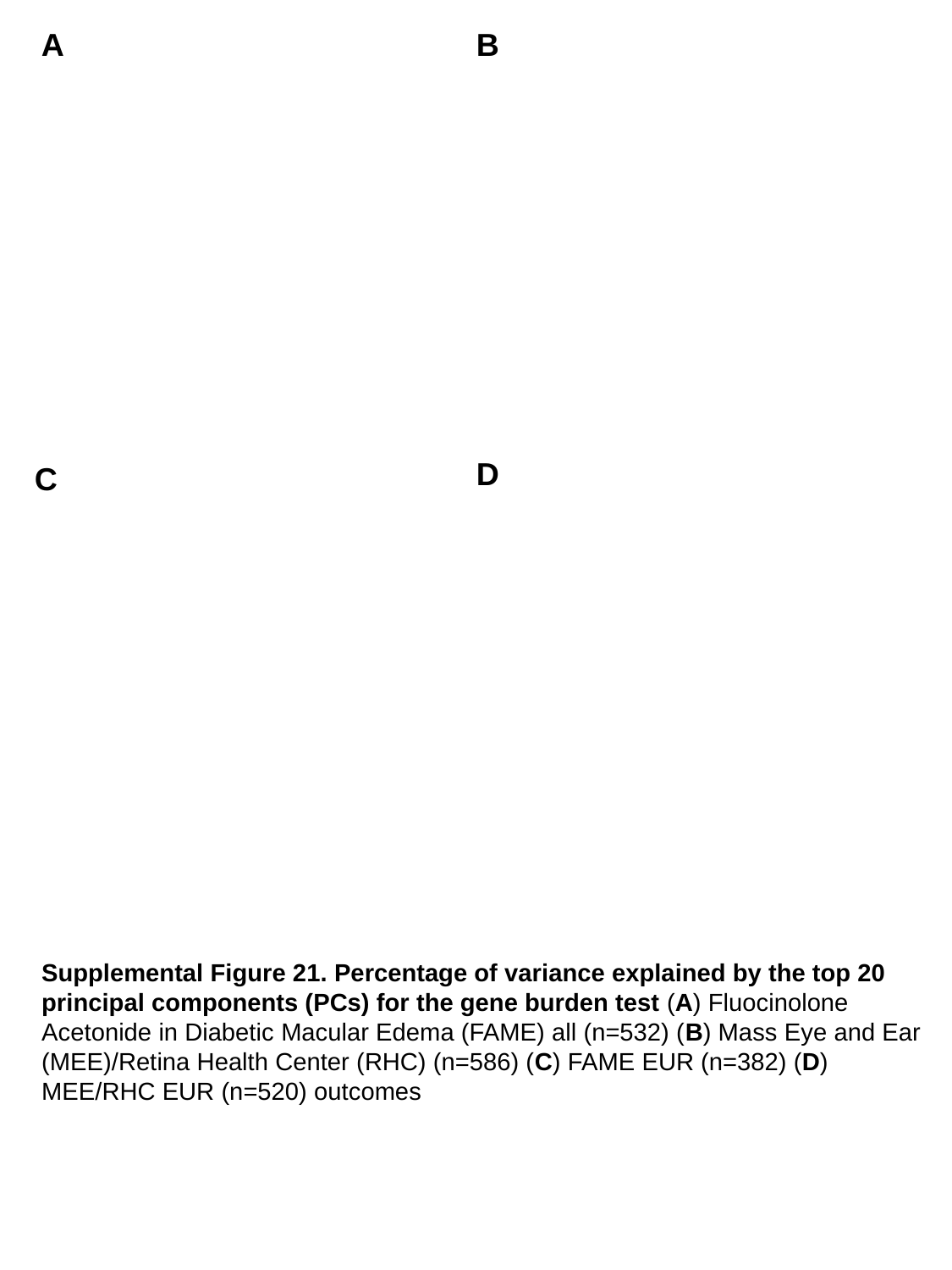

A
B
D
C
Supplemental Figure 21. Percentage of variance explained by the top 20 principal components (PCs) for the gene burden test (A) Fluocinolone Acetonide in Diabetic Macular Edema (FAME) all (n=532) (B) Mass Eye and Ear (MEE)/Retina Health Center (RHC) (n=586) (C) FAME EUR (n=382) (D) MEE/RHC EUR (n=520) outcomes

### Slide 34
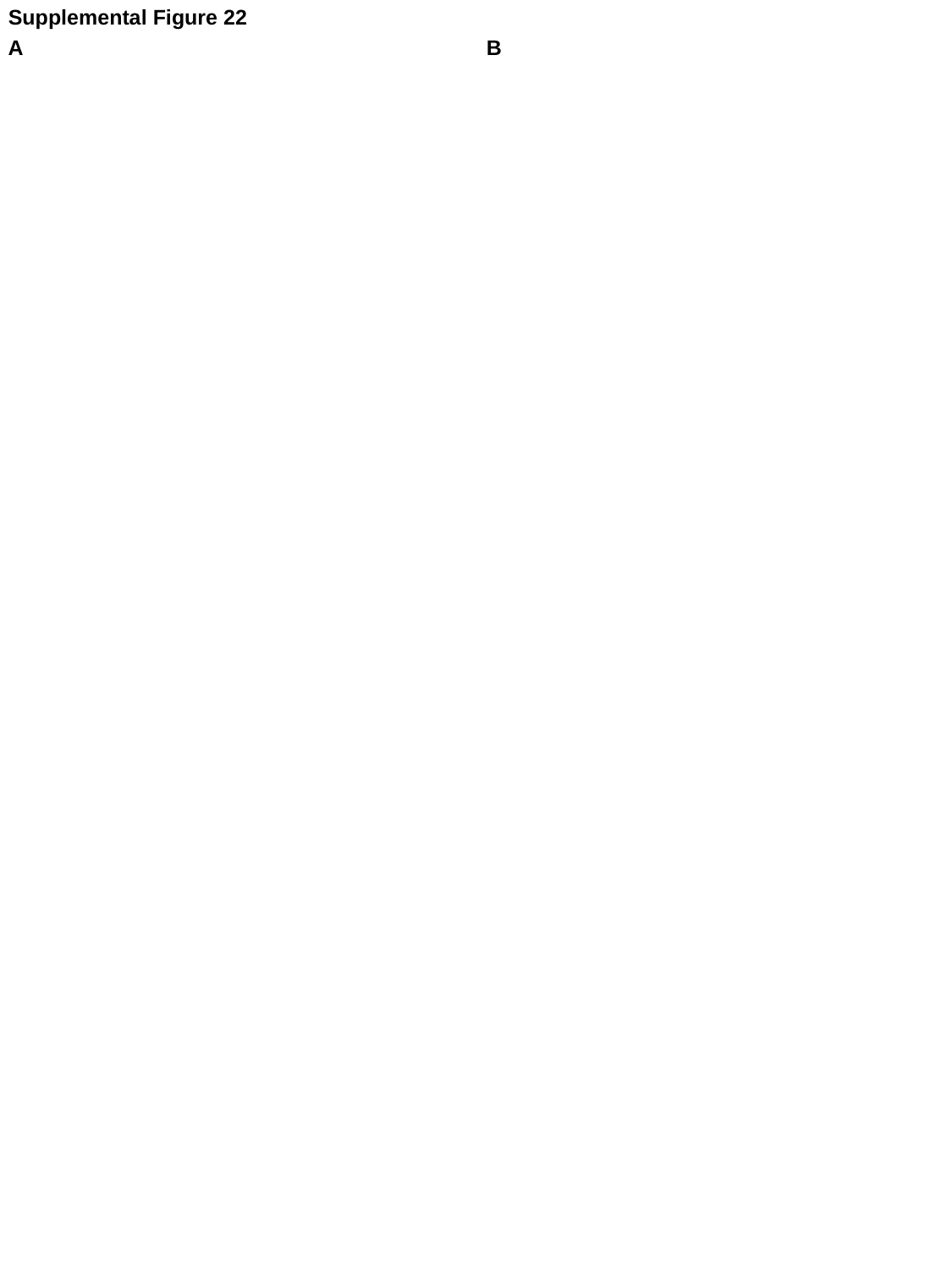

Supplemental Figure 22
A
B

### Slide 35
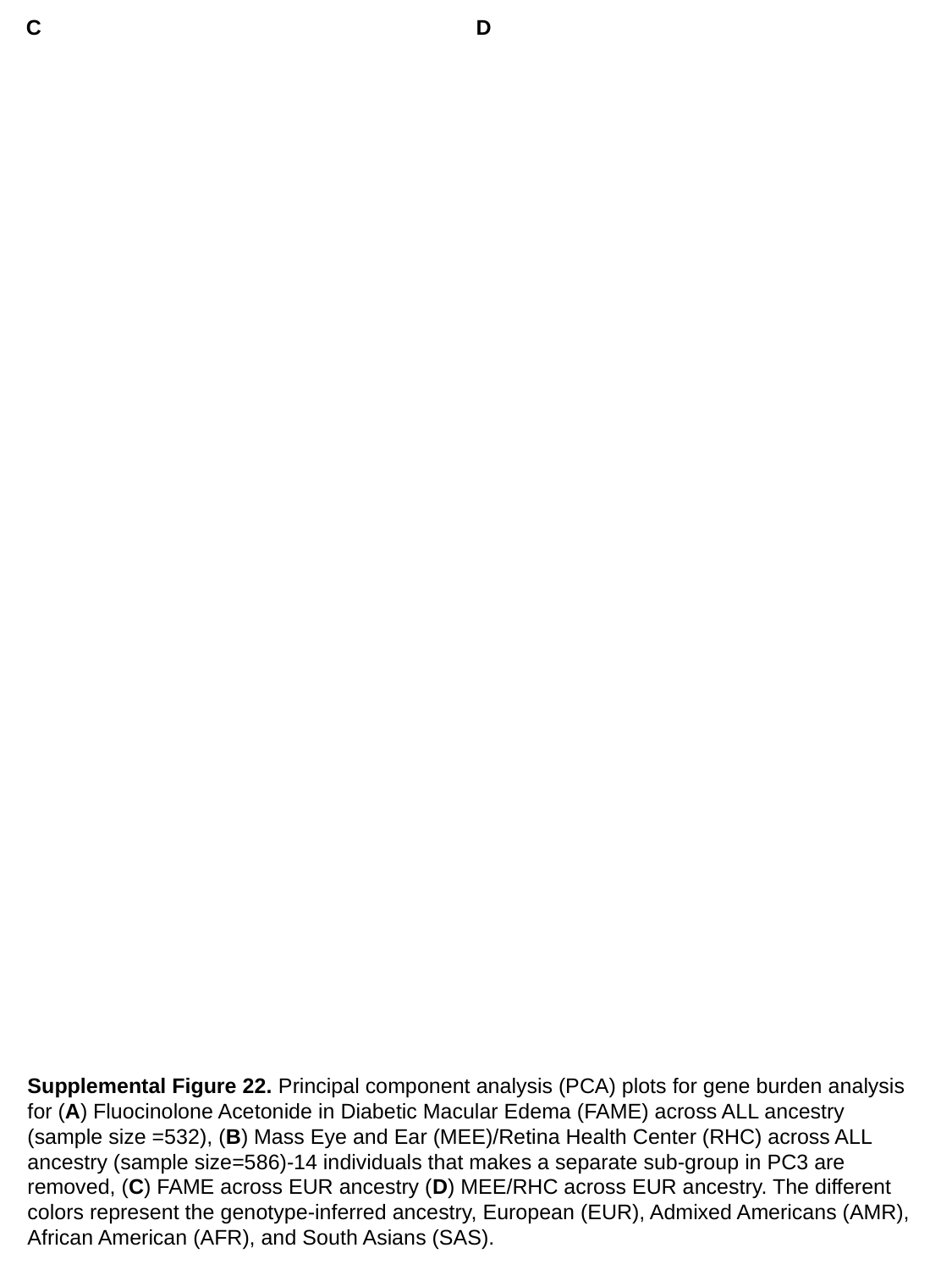

C
D
Supplemental Figure 22. Principal component analysis (PCA) plots for gene burden analysis for (A) Fluocinolone Acetonide in Diabetic Macular Edema (FAME) across ALL ancestry (sample size =532), (B) Mass Eye and Ear (MEE)/Retina Health Center (RHC) across ALL ancestry (sample size=586)-14 individuals that makes a separate sub-group in PC3 are removed, (C) FAME across EUR ancestry (D) MEE/RHC across EUR ancestry. The different colors represent the genotype-inferred ancestry, European (EUR), Admixed Americans (AMR), African American (AFR), and South Asians (SAS).

### Slide 36
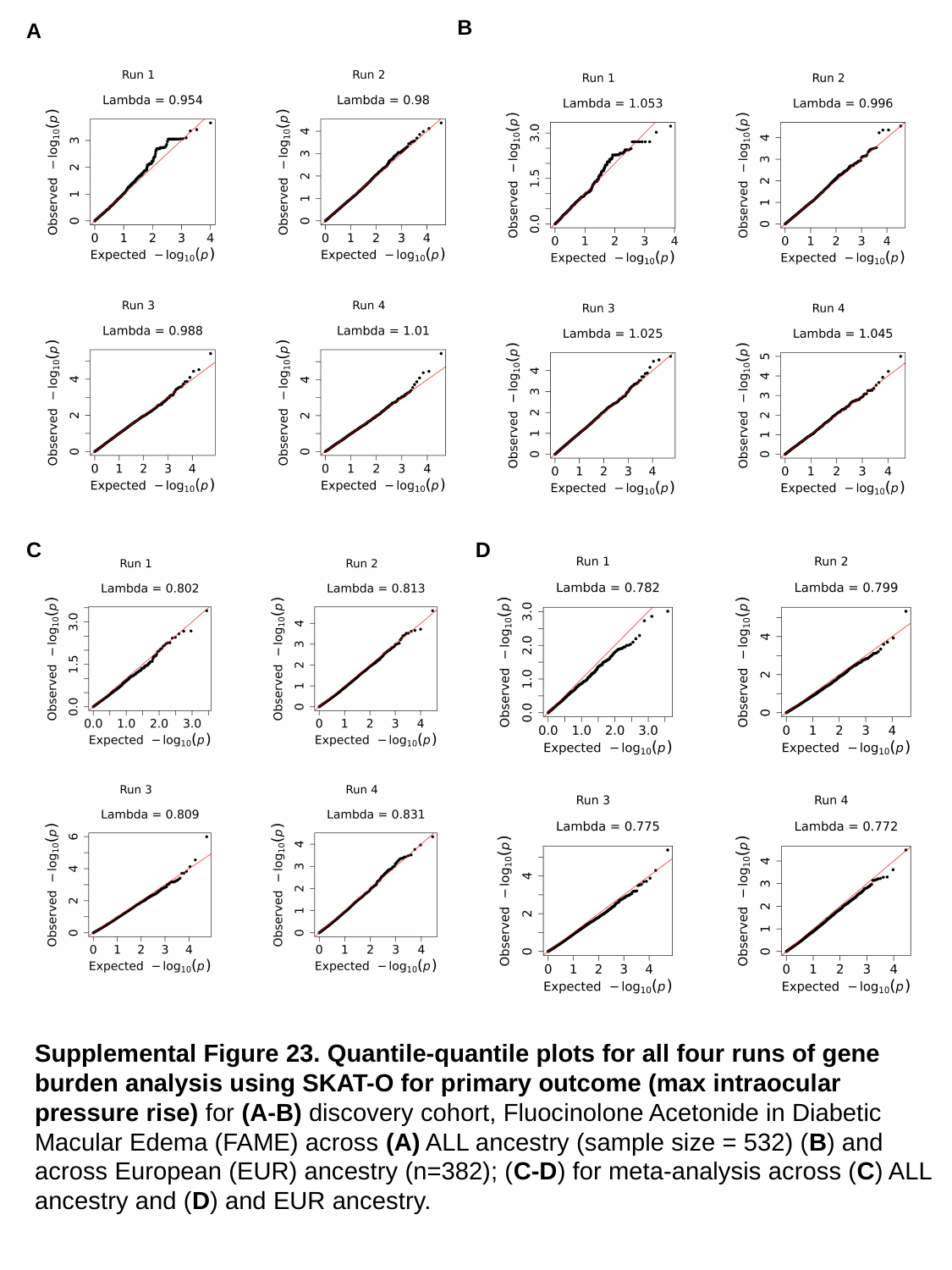

B
A
C
D
Supplemental Figure 23. Quantile-quantile plots for all four runs of gene burden analysis using SKAT-O for primary outcome (max intraocular pressure rise) for (A-B) discovery cohort, Fluocinolone Acetonide in Diabetic Macular Edema (FAME) across (A) ALL ancestry (sample size = 532) (B) and across European (EUR) ancestry (n=382); (C-D) for meta-analysis across (C) ALL ancestry and (D) and EUR ancestry.

### Slide 37
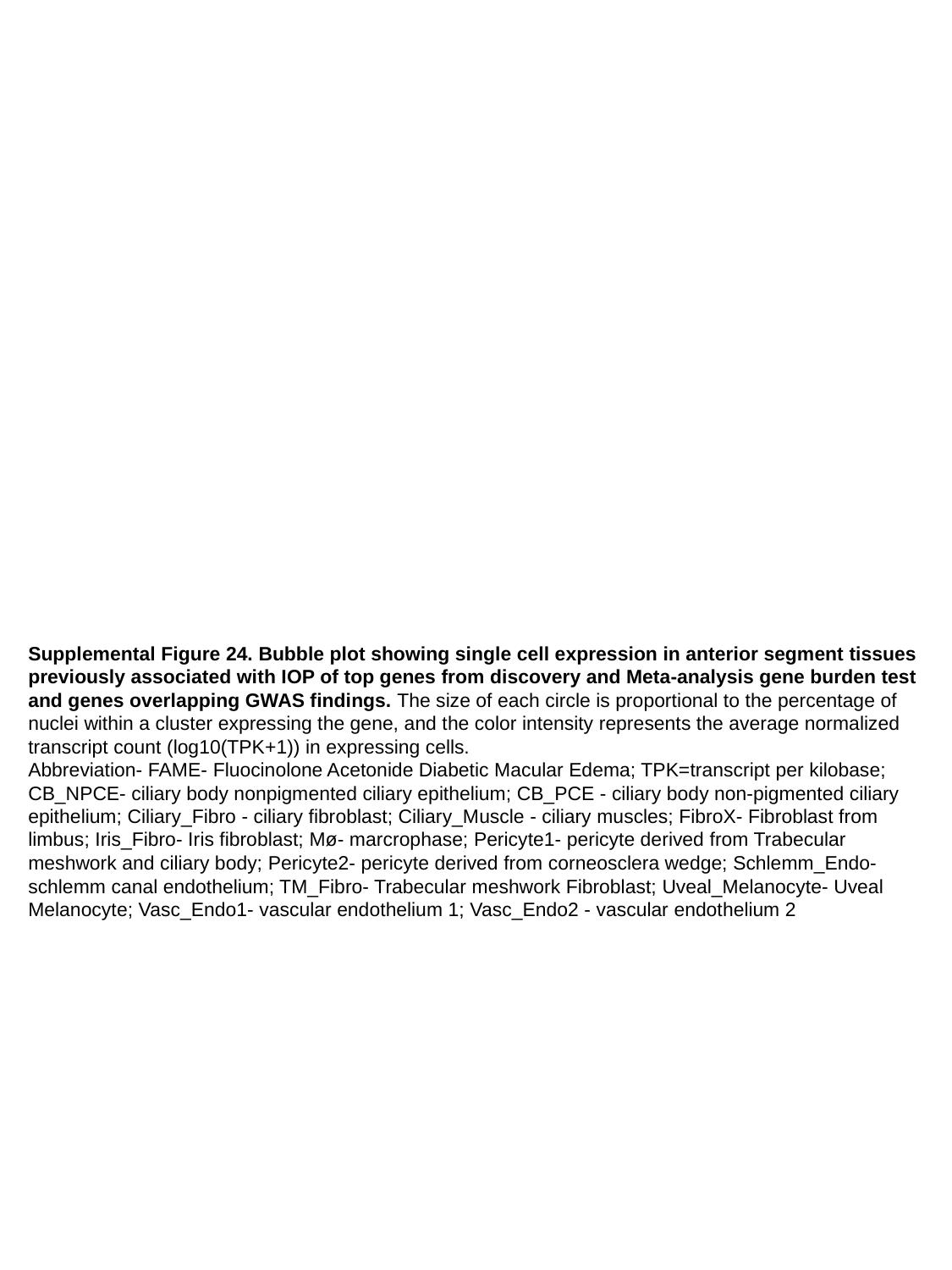

Supplemental Figure 24. Bubble plot showing single cell expression in anterior segment tissues previously associated with IOP of top genes from discovery and Meta-analysis gene burden test and genes overlapping GWAS findings. The size of each circle is proportional to the percentage of nuclei within a cluster expressing the gene, and the color intensity represents the average normalized transcript count (log10(TPK+1)) in expressing cells.
Abbreviation- FAME- Fluocinolone Acetonide Diabetic Macular Edema; TPK=transcript per kilobase; CB_NPCE- ciliary body nonpigmented ciliary epithelium; CB_PCE - ciliary body non-pigmented ciliary epithelium; Ciliary_Fibro - ciliary fibroblast; Ciliary_Muscle - ciliary muscles; FibroX- Fibroblast from limbus; Iris_Fibro- Iris fibroblast; Mø- marcrophase; Pericyte1- pericyte derived from Trabecular meshwork and ciliary body; Pericyte2- pericyte derived from corneosclera wedge; Schlemm_Endo- schlemm canal endothelium; TM_Fibro- Trabecular meshwork Fibroblast; Uveal_Melanocyte- Uveal Melanocyte; Vasc_Endo1- vascular endothelium 1; Vasc_Endo2 - vascular endothelium 2
