## Supplementary Tables for "Genome-Wide Association Study for Glucocorticoid-Induced Ocular Hypertension"

**Supplemental Table 1. Genome-wide genotyping: Pre-Imputation Sample and Variant Quality Control (QC)**

| QC steps | Cutoff | Number (#) of removed samples | # of retained samples | # of variants removed | # of retained variants |
| --- | --- | --- | --- | --- | --- |
| Original data (1,376 IDs) | - | - | 1376 | - | 729,900 |
| Exclude multiallelic variants | - | - | 1376 | 96 | 729,804 |
| Pre-Imputation Sample QC | | | | | |
| Exclude samples with missingness (>2%) Minimum call rate ~0.9 | (>2%) | 0 | 1376 | - | 729,804 |
| Heterozygosity test^1^ | +/-5std | 10 | 1376 | - | 729,804 |
| Sex check | F=0.2 | 5 | 1362* | - | 729,804 |
| Ancestry check |  | 5 | 1357 | - | 729,804 |
| Genome identity-by-descent check^2^ | >0.1875 | 6 | 1351 | - | 729,804 |
| Total sample QC failed | - | 25 | 1351 | - | - |
| Pre-Imputation Variant QC | | | | | |
| Mitochondria variant removed | - | - | 1351 | 1,245 | 728,559 |
| Remove large Indels | >50 bp | - | 1351 | 129 | 728,430 |
| Exclude heterozygous haploid genotypes on sex chromosome in males | Non-pseudoautosomal region (PAR) | - | 1351 | 0 | 728,430 |
| Exclude variant genotyping call rate | >2% | - | 1351 | 12,086 | 716,344 |
| Exclude monomorphic variants  1/(2*N)=1/(2*1352) | 0.00037 | - | 1351 | 124,713 | 591,631 |
| Testing Hardy-Weinberg Equilibrium  (only on variants with minor allele frequency > 0.01) | P=10^-6^ | - | 1351 | 1,674 | 589,957 |
| Remove strand ambiguous variants (A/T,C/G) | - | - | 1351 | 6,412 | 583,545 |
| Remove sample duplicates | - | 0 | 1351 | - | 583,545 |
| Remove variants with multiple same copies | - | - | 1351 | 4,521 | 579,024 |
| Fix reference/alternate flipped variants | - | - | 1351 | 84,629 | 579,024 |
| Remove multiallelic variants | - | - | 1351 | 31 | 578,993 |
| Total remaining pre-imputation data (663 FAME + 688 MEE/RHC=1,351 sample IDs) | - | - | 1351 | - | 578,993 |

^1^Heterozygosity test was performed separately for each of the 5 ancestry groups (AMR, AFR, EUR, SAS, EAS)

^2^Among IBD failed: 5 pairs were duplicates with pi-hat>0.9, 1 pair were first degree relatives with pi-hat= 0.2693

*One sample failed both sex check and heterozygosity test

Abbreviations: QC-quality control, #-number

### **Supplemental Table 2. Whole exome sequencing data: Exome sample Quality Control**

| **Filter Criteria** | **Number of samples that failed QC step** |
| --- | --- |
| Original sample (1,376 IDs) | - |
| Sex mismatch | 3 |
| Remove related individuals (Identity-by-descent PI_HAT > 0.1875) | 5 |
| Ancestry mismatch | 7 |
| het/hom outlier (above or below 4 std) | 6 |
| No. of singleton outlier (above or below 4 std) | 1 |
| Percentage chimera outlier (above or below 4std) | 1 |
| Percentage contamination outlier (above or below 4std) | 2 |
| genotype quality outlier (above or below 4std) | 7 |
| Transition/Transversion outlier (above or below 8 std) | 1 |
| No. of SNPs outlier (above or below 4std) | 10 |
| No. of Deletions outlier (above or below 4std) | 6 |
| No. of Insertions outlier (above or below 4std) | 5 |
| *Total Samples that failed | 31 |
| Remaining Samples | 1,345 |

*Many samples failed one or more of the above filter criteria used for sample QC

**Supplemental Table 3. Whole exome sequencing data: Exome variant Quality Control**

| Filtering criteria | Total sites | Single nucleotide polymorphism | InDel | Multiallelic sites  (Split) | Minor allele frequency (MAF)>0.01 | MAF>0.05 |
| --- | --- | --- | --- | --- | --- | --- |
| Initial variant calls (1345 IDs) | 3,269,192 | 2,798,015 | 471,177 | 409,955 | 537,826 | 311,986 |
| In low complexity region | 304,227 | 76,365 | 227,862 | - | 121,761 | 75,437 |
| Variant quality score recalibration | 247,926 | 166,151 | 81,775 | - | 46,823 | 26,053 |
| Low Quality | - | - | - | - | - | - |
| Excess Heterozygosity >54.69 | 92,846 | 22,162 | 70,684 | - | 63,729 | 44,931 |
| Monomorphic | 362,944 | 279,341 | 83,603 | - | - | - |
| Missingness (AB,DP,GQ)** | 1,310,038 | 983,949 | 326,089 | - | 395,570 | 252,376 |
| Polymerase chain reaction (PCR) batch association (P<10-8) | - | - | - | - | - | - |
| SEQ | - | - | - | - | - | - |
| LCSET | - | - | - | - | - | - |
| Non-random missingness test (HMISS) failed Plink non-random missingness test (P<5X10^-8^) | 4,046 | 3,708 | 338 | - | 639 | 377 |
| Hardy-Weinberg Equilibrium (HWE) test for AFR *** | - | - | - | - | - | - |
| HWE test for AMR | - | - | - | - | - | - |
| HWE test for EAS | - | - | - | - | - | - |
| HWE test for EUR | 44,501 | 23,324 | 21,177 | - | 44,285 | 43,308 |
| HWE test for SAS | 3,982 | 2,095 | 1,887 | - | 3,980 | 3,970 |
| HWE for AFR in Female | - | - | - | - | - | - |
| HWE for AMR in Female | - | - | - | - | - | - |
| HWE for EAS in Female | - | - | - | - | - | - |
| HWE for EUR in Female | 84 | 60 | 24 | - | 84 | 84 |
| HWE for SAS in Female | - | - | - | - | - | - |
| HWE for AFR in Male | - | - | - | - | - | - |
| HWE for AMR in Male | - | - | - | - | - | - |
| HWE for EAS in Male | - | - | - | - | - | - |
| HWE for EUR in Male | 82 | 57 | 25 | - | 82 | 81 |
| HWE for SAS in Male | - | - | - | - | - | - |
| PASS | 1,814,244 | 1,717,915 | 96,329 | - | 244,709 | 151,795 |
| *VQSR cutoff were set to 99.8% for SNPs and 99.95% for InDels | | | | | | |
| **Allelic balance (AB) <=0.2 and >= 0.8; Missingness 2%; genotype quality (GQ)=20; read depth (DP)=10; | | | | | | |
| ***HWE ran on population with >=100 samples; HWE test performed on all alleles with MAF >0.01 filter (P<10-8) | | | | | | |

Abbreviations: InDel-insertion/deletion, MAF-minor allele frequency, HWE-Hardy-Weinberg Equilibrium

**Supplemental Table 4. Genome-wide genotyping: Distribution of imputation quality INFO score across three minor allele frequency (MAF) groups**

| MAF <0.01 | |  | MAF 0.01-0.05 | |  | MAF >0.05 | |
| --- | --- | --- | --- | --- | --- | --- | --- |
| Statistic | INFO, r^2^ |  | Statistic | INFO, r^2^ |  | Statistic | INFO, r^2^ |
| Mean | 0.73 |  | Mean | 0.92 |  | Mean | 0.96 |
| Median | 0.88 |  | Median | 0.95 |  | Median | 0.97 |
| Max | 1 |  | Max | 1.00 |  | Max | 1.00 |
| Min | 0.1 |  | Min | 0.00 |  | Min | 0.01 |

Abbreviations: MAF-minor allele frequency

### **Supplemental Table 5. Post-imputation genotype variant quality control (QC) of discovery cohort Fluocinolone Acetonide in Diabetic Macular Edema (FAME) and replication cohort, Mass Eye and Ear/Retina Health Center (MEE/RHC).**

| QC steps | Cutoff | Number (#) of sample IDs | # of variants removed | # of variants kept |
| --- | --- | --- | --- | --- |
| Post-imputed data (1,351 FAME+MEE sample IDs) | - | 1,351 | - | 444,721,145 |
| Removed 185 IDs which failed meeting the final study inclusion criteria* | - | 1,166 | - | - |
| FAME | | | | |
| INFO Score (R^2^) filter | >=0.6 | 532 | - | 46,623,344 |
| Remove large Indels | >50 bp | 532 | - | 46,623,344 |
| Exclude heterozygous haploid genotypes on sex chromosome in males | - | 532 | - | 46,623,344 |
| Exclude variant genotyping call rate | >2% | 532 | 4,533 | 46,623,344 |
| Minor allele frequency | >0.01 | 532 | 37,174,379 | 9,448,975 |
| Hardy Weinberg Equilibrium test | 1x10^-8^ | 532 | 105 | 9,448,884 |
| Total Remaining | - | 532 | - | 9,448,884 |
| MEE/RHC | | | | |
| InfoScore (R^2^) filter | >=0.6 | 634** | - | 46,623,344 |
| Remove large Indels | >50 bp | 634 | 0 | 46,623,344 |
| Exclude heterozygous haploid genotypes on sex chromosome in males | - | 634 | 0 | 46,623,344 |
| Exclude variant genotyping call rate | >2% | 634 | 2,204 | 46,621,140 |
| Minor allele frequency | >0.01 | 634 | 37,050,968 | 9,572,386 |
| Hardy Weinberg Equilibrium test | 1x10^-8^ | 634 | 181 | 9,572,205 |
| Total Remaining | - | 634 | - | 9,572,205 |

Abbreviations: QC-quality control, FAME-Fluocinolone Acetonide in Diabetic Macular Edema, MEE/RHC-Mass Eye and Ear/Retina Health Center, #-number

* ”excluded from further analysis” flag consists of samples with perioperative steroid use within 3 months, pre-existing steroid use by first visit, glaucoma procedure prior to steroid use, missing follow-up intraocular pressures due to loss to follow-up, pre-existing hypotony, history of endophthalmitis, or history of trauma.

**The 634 samples in MEE/RHC include 31 samples with missing covariates and 15 samples (8 EAS and 7 SAS) that were later excluded from GWAS analysis due to small population sample size. Therefore, the final number of samples used in GWAS was 588.

**Supplemental Table 6. Phenotypes tested in genome-wide association study (GWAS) and number of genome-wide significant or subthreshold genomic loci for discovery cohort, Fluocinolone Acetonide in Diabetic Macular Edema (FAME), replication cohorts, Mass Eye and Ear/Retina Health Center (MEE/RHC), and meta-analysis combining discovery and replication cohorts (FAME-MEE/RHC).**The table shows the summary of all phenotypes/traits and populations tested in GWAS using PLINK and lists the number of samples, genomic control (GC) inflation statistic (lambda), and the total linkage disequilibrium (LD)-independent variants that passed genome wide significance and subthreshold significance (P<1x10^-5^).

| **Discovery cohort (FAME)** | | | | | | |
| --- | --- | --- | --- | --- | --- | --- |
| **GWAS Analysis** | **Trait** | **Population** | **Number**  **of samples** | **GC lambda** | **Number of significant linkage disequilibrium (LD)-independent variants (P<5E-8)** | **LD-independent variants at P<1E-5** |
| Primary Outcome | Max Intraocular Pressure (IOP) Change | All | 530 | 1.01 | 1 | 40 |
| Primary Outcome | Max IOP Change | EUR | 372 | 1.00 | 0 | 24 |
| Secondary Outcome | Responders | All | 530 | 1.03 | 0 | 16 |
| Secondary Outcome | Extreme Responders | All | 244 | 1.05 | 0 | 2 |
| **Replication cohort (MEE/RHC)** | | | | | | |
| Primary Outcome | Max IOP Change | All | 577 | 1.04 | 2 | 62 |
| Primary Outcome, Sensitivity Analysis of Intravitreal Injections Only | Max IOP Change | All | 437 | 1.01 | 0 | 40 |
| EUR Primary | Max IOP Change | EUR | 495 | 0.99 | 0 | 26 |
| EUR Primary Outcome, Sensitivity Analysis of Intravitreal Injections Only | Max IOP Change | EUR | 385 | 1.00 | 1 | 31 |
| Secondary | Responders | All | 588 | 1.03 | 0 | 10 |
| Secondary | Extreme Responders | All | 306 | 1.14 | 0 | 8 |
| **FAME-MEE/RHC Meta-analysis** | | | | | | |
| Primary Outcome | Max IOP Change | All | 1107 | 1.01 | 1 | 44 |
| Primary Outcome, Sensitivity Analysis of Intravitreal Injections Only | Max IOP Change | All | 967 | 1.00 | 0 | 33 |
| EUR Primary Outcome | Max IOP Change | EUR | 867 | 1.00 | 0 | 20 |
| EUR Primary Outcome, Sensitivity Analysis of Intravitreal Injections Only | Max IOP Change | EUR | 757 | 1.01 | 0 | 24 |
| Secondary Outcome | Responders | All | 832 | 1.00 | 0 | 9 |
| Secondary Outcome | Extreme Responders | All | 550 | 1.06 | 0 | 1 |

Abbreviations: GWAS-Genome-wide association study, FAME-Fluocinolone Acetonide in Diabetic Macular Edema, MEE/RHC-Mass Eye and Ear/Retina Health Center, LD-linkage disequilibrium, IOP-intraocular pressure, Max-maximal, EUR-European

**Supplemental Table 7. Rare variant functional effect categories for gene burden tests**

| Gene burden mask/runs | Categories | Type of mutation |
| --- | --- | --- |
| Run 1 | Loss of Function1 (LoF1) | Stop gained, splice acceptor, splice donor, frameshift, transcript ablation |
| Run 2 | LoF1 | Stop gained, splice acceptor, splice donor, frameshift, transcript ablation |
|  | Loss of Function2 (LoF2) | Stop lost, start lost, transcription amplification, feature elongation, feature truncation |
|  | Missense | Missense variants |
| Run 3 | LoF1 | Stop gained, splice acceptor, splice donor, frameshift, transcript ablation |
|  | LoF2 | Stop lost, start lost, transcription amplification, feature elongation, feature truncation |
|  | Missense | Missense variants |
|  | Moderate | In-frame insertion, in-frame deletion, protein altering |
|  | Low | Splice region, splice donor 5^th^ base variant, splice donor variant, splice polypyrimidine tract variant, incomplete terminal codon |
|  | Modifier | 3' untranslated region (UTR) variant, 5'UTR variant, transcription factor (TF) binding site varaint, intergenic, intron variant, mature microRNA, non-coding transcript, non-coding transcript exon variant, regulatory region ablation and amplification, upstream and downstream, TF binding sites ablation and amplification, coding sequence variant, regulatory region ablation and amplification, intergenic variant, sequence variant, Nonsense-mediated decay (NMD) transcript varaint |
| Run 4 | Synonymous (control group) | Synonymous, start retrained, stop retrained |

Abbreviations: LoF1-loss of function1, LoF2-loss of function2, UTR-untranslated region, TF-transcription factor

**Supplemental Table 8. List of number of rare variants used in gene burden test**

|  | Dichotomous Outcome (Responders) | | | | Quantitative Outcome (Maximal IOP Change) | | | | Dichotomous (Extreme Responders) | | | |
| --- | --- | --- | --- | --- | --- | --- | --- | --- | --- | --- | --- | --- |
|  | FAME (N=532) | FAME EUR (N=382) | MEE/RHC (N=586) | MEE/RHC EUR (N=520) | FAME (N=532) | FAME EUR (N=382) | MEE/RHC (N=575) | MEE/RHC EUR (509) | FAME (N=228) | FAME EUR (N=188) | MEE/RHC (N=297) | MEE/RHC EUR (264) |
| Total (1345 sample IDs) | 1,814,246 | 1,814,246 | 1,814,246 | 1,814,246 | 1,814,246 | 1,814,246 | 1,814,246 | 1,814,246 | 1,814,246 | 1,814,246 | 1,814,246 | 1,814,246 |
| Removed mitochondrial variants (1345 sample IDs) | 1,812,940 | 1,812,940 | 1,812,940 | 1,812,940 | 1,812,940 | 1,812,940 | 1,812,940 | 1,812,940 | 1,812,940 | 1,812,940 | 1,812,940 | 1,812,940 |
| Keep* |  | | | | | | | | | | | |
| Removed off-target variants | 1,812,940 | 1,812,940 | 1,812,940 | 1,812,940 | 1,812,940 | 1,812,940 | 1,812,940 | 1,812,940 | 1,812,940 | 1,812,940 | 1,812,940 | 1,812,940 |
| Missingness (>2%) | 1,809,494 | 1,808,080 | 1,790,294 | 1,790,587 | 1,809,494 | 1,808,080 | 1,791,663 | 1,792,060 | 1,804,971 | 1,784,610 | 1,775,387 | 1,800,792 |
| Exclude monomorphic variants | 1,108,449 | 704,327 | 1,084,963 | 828,355 | 1,108,449 | 704,327 | 1,078,884 | 514,510 | 609,690 | 510,356 | 502,603 | 510,356 |
| Rare variants used in Gene Burden (MAF<0.01) | 866,551 | 481,522 | 840,135 | 609,179 | 866,551 | 481,522 | 834,769 | 296,184 | 382,775 | 284,235 | 246,092 | 284,234 |
| % of rare variants | 78% | 68% | 77% | 74% | 78% | 68% | 77% | 58% | 62% | 56% | 49% | 56% |

*Only keeping samples used in gene burden test separately for each cohort. The number of samples kept are in the column names represented by N.

Abbreviations: IOP-intraocular pressure, FAME-Fluocinolone Acetonide in Diabetic Macular Edema, MEE/RHC-Mass Eye and Ear/Retina Health Center, MAF-Minor allele frequency

**Supplemental Table 9. Top variants associated with glucocorticoid-induced intraocular pressure rise (IOP) GWAS using REGENIE.** The table shows all the variants that were genome-wide significant (P< 5x10^-8^) and/or replicated in both cohorts- Fluocinolone Acetonide in Diabetic Macular Edema (FAME) discovery cohort and Mass Eye and Ear/Retina Health Center (MEE/RHC) replication cohort. Listed are the summary statistics for GWAS in discovery FAME cohort, replication MEE/RHC cohort and the FAME-MEE/RHC meta-analysis sorted by their P-values within their trait. The Effect Allele (A1), Effect Allele’s frequency, P-values, direction of effect, heterozygosity P-value, replication Bonferroni cutoff and genotype imputation quality (R^2^, INFO score) are provided. The table also shows RINT’ed and raw effect size (Beta) and standard error (SE) from GWAS using rank inverse normal transformed (RINT) and pre-transformed maximal IOP rise in original unit scale (mmHg), respectively. Minor allele frequency (MAF) of the variant within each ancestry was calculated using the study population in which the variant was genome-wide significant or passed replication Bonferroni cutoff. The nearest gene and most deleterious consequences were annotated using Ensembl’s Variant Effect Predictor (VEP).

| Variant ID  hg38 | Trait | **FAME Primary** | | | | | **MEE/RHC Primary** | | | | **Meta-Analysis Primary** | | | | | | | | R^2^ (Info Score) | Nearest Gene | Deleterious  Consequences |
| --- | --- | --- | --- | --- | --- | --- | --- | --- | --- | --- | --- | --- | --- | --- | --- | --- | --- | --- | --- | --- | --- |
|  |  | Effect Allele (A1) | A1 Freq | BETA (RINT) | SE (RINT) | P-value | BETA (RINT) | SE (RINT) | P-value | Bonf. adj  P-value | Effect (RINT) | SE (RINT) | P-value | Bonf. adj P-value | Direction | Het P-value | Effect Raw  (mmHg) | SE Raw  (mmHg) |  |  |  |
| chr17:8838263:C:T | Primary ALL | T | 0.03 | -0.55 | 0.13 | 4.8E-05 | -0.56 | 0.16 | 2.5E-04 | 0.01 | -0.56 | 0.1 | 4.5E-08** | 2.2E-06 | -- | 0.82 | -2.9 | 0.58 | 0.88 | *PI3KR6* | MANE intron variant |
| chr2:181021297:G:A | Primary ALL | A | 0.05 | -0.55 | 0.11 | 3.0E-07 | -0.25 | 0.12 | 0.04 | 2.07 | -0.42 | 0.08 | 2.0E-07 | 1.0E-05* | -- | 0.07 | -2.01 | 0.46 | 0.91 | *UBE2E3* | MANE intron variant |
| chr2:181019696:A:C | Primary ALL | C | 0.05 | -0.55 | 0.11 | 3.1E-07 | -0.25 | 0.12 | 0.04 | 1.90 | -0.42 | 0.08 | 2.0E-07 | 9.9E-06* | -- | 0.06 | -2.00 | 0.46 | 0.91 | *UBE2E3* | MANE intron variant |
| chr1:111045611:C:T | Primary ALL | T | 0.01 | -1.08 | 0.23 | 3.6E-06 | -0.55 | 0.22 | 0.01 | 0.65 | -0.81 | 0.16 | 6.0E-07 | 3.0E-05* | -- | 0.10 | -3.42 | 0.90 | 0.95 | *LRIF1* | Downstream gene variant, regulatory region variant |
| chr1:111046239:C:A | Primary ALL | A | 0.98 | 1.14 | 0.25 | 3.8E-06 | 0.57 | 0.29 | 0.05 | 2.21 | 0.90 | 0.19 | 1.5E-06 | 7.6E-05* | ++ | 0.13 | 3.87 | 1.05 | 0.66 | *LRIF1* | Downstream gene variant |
| chr6:148187527:T:G | Primary ALL | G | 0.03 | -0.61 | 0.13 | 6.0E-06 | -0.27 | 0.14 | 0.04 | 2.19 | -0.44 | 0.10 | 3.6E-06 | 1.8E-04* | -- | 0.08 | -2.15 | 0.55 | 0.98 | *SASH1* | Intergenic variant |
| chr6:148233400:C:T | Primary ALL | T | 0.03 | -0.62 | 0.14 | 6.0E-06 | -0.27 | 0.13 | 0.04 | 1.98 | -0.44 | 0.10 | 3.8E-06 | 1.9E-04* | -- | 0.07 | -2.20 | 0.55 | 0.99 | *SASH1* | Downstream gene variant |
| chr17:33083254:T:C | Primary ALL | C | 0.18 | 0.27 | 0.06 | 8.7E-06 | 0.11 | 0.06 | 0.05 | 2.40 | 0.19 | 0.04 | 7.3E-06 | 3.6E-04* | ++ | 0.06 | 1.06 | 0.24 | 0.99 | *SPACA3* | MANE intron variant |
| chr17:33083893:A:T | Primary ALL | T | 0.18 | 0.28 | 0.06 | 9.1E-06 | 0.11 | 0.06 | 0.05 | 2.41 | 0.19 | 0.04 | 7.7E-06 | 3.8E-04* | ++ | 0.06 | 1.06 | 0.24 | 0.99 | *SPACA3* | MANE intron variant |
| chr17:33083254:T:C | Primary Injections ALL | C | 0.18 | 0.27 | 0.06 | 8.7E-06 | 0.13 | 0.06 | 0.02 | 1.05 | 0.20 | 0.04 | 2.5E-06 | 1.2E-04* | ++ | 0.09 | 0.98 | 0.24 | 0.99 | *SPACA3* | MANE intron variant |
| chr17:33083893:A:T | Primary Injections ALL | T | 0.18 | 0.28 | 0.06 | 9.1E-06 | 0.13 | 0.06 | 0.02 | 1.00 | 0.20 | 0.04 | 2.4E-06 | 1.2E-04* | ++ | 0.09 | 0.98 | 0.24 | 0.99 | *SPACA3* | MANE intron variant |
| chr17:33083893:A:T | Primary Injections EUR | T | 0.19 | 0.27 | 0.06 | 1.6E-06 | 0.14 | 0.06 | 0.02 | 0.79 | 0.21 | 0.04 | 2.7E-07 | 1.2E-05* | ++ | 0.1327 | 1.19 | 0.26 | 0.99 | *SPACA3* | MANE intron variant |
| chr17:33083254:T:C | Primary Injections EUR | C | 0.19 | 0.27 | 0.06 | 1.6E-06 | 0.14 | 0.06 | 0.02 | 0.78 | 0.21 | 0.04 | 2.7E-07 | 1.2E-05* | ++ | 0.1345 | 1.19 | 0.26 | 0.99 | *SPACA3* | MANE intron variant |
| chr4:8737331:A:C | Primary Injections EUR | C | 0.06 | -0.44 | 0.1 | 7.8E-06 | -0.24 | 0.11 | 0.04 | 1.75 | -0.35 | 0.07 | 2.2E-06 | 9.6E-05* | -- | 0.1799 | -1.61 | 0.46 | 0.82 | *HMX1* | Intergenic variant |

**P-value passes genome-wide significance (P<5E-08); *P-value passes replication Bonferroni cutoff.

Abbreviations: GWAS- Genome-Wide Association Study, FAME- Fluocinolone Acetonide in Diabetic Macular Edema, MEE/RHC- Mass Eye and Ear/Retina Health Center, A1- effect allele, RINT- Rank Inverse Normal Transformation, SE-Standard Error, Raw Beta- effect size from GWAS using original unit scale (mmHg rise or fall of IOP), Raw SE- standard error from GWAS using original unit scale (mmHg rise or fall of IOP), INFO score- genotype imputation quality score. EUR- European, ALL includes EUR, AFR- African, AMR-Admixed American, and SAS- South Asian. MANE- Matched Annotation from NCBI and EMBL-EBI.

**Supplemental Table 10. Top genome-wide association study hits for Fluocinolone Acetonide in Diabetic Macular Edema (FAME) samples across ALL and EUR populations with primary outcome (Maximal intraocular pressure rise) (P<10^-5^) (n=530 and 372).** The table shows the summary statistics for linkage disequilibrium (LD)-independent variants (LD clumping at r^2^ > 0.1 in 500kb sliding window) that passed P<5x10^-7^. Listed are the Effect Allele (A1), Effect Allele’s (A1) frequency, effect size of the A1 allele (Beta) for the rank inverse normal transformed (RINT) maximal intraocular pressure (IOP) rise phenotype, Standard Error (SE), P-value, and genotype imputation quality (R^2^, INFO score). The table also shows effect size (Raw Beta) and standard errors (Raw SE) for raw (pre-transformed) maximal IOP rise at the original mmHg change per month units, and minor allele counts (MAC) for four ancestral groups: European (EUR), Admixed American (AMR), African American (AFR), and South Asian (SAS). The nearest gene and most deleterious consequences were annotated using Ensembl’s Variant Effect Predictor (VEP) and the Matched Annotation from NCBI and EMBL-EBI (MANE) transcripts.

| Variant ID (hg38) | Rs ID | Effect Allele (A1) | A1 Frequency | Beta (RINT) | Standard Error (SE) | P-value | Raw Beta | Raw SE | INFO(R^2^) | Nearest Gene | Deleterious Consequences | EUR Minor Allele Count (MAC)* | AMR MAC* | AFR MAC* | SAS MAC* |
| --- | --- | --- | --- | --- | --- | --- | --- | --- | --- | --- | --- | --- | --- | --- | --- |
| **FAME Primary ALL** | | | | | | | | | | | | | | | |
| chr2:181004816:G:C | rs13425173 | C | 0.056 | -0.66 | 0.12 | 2.9E-08 | -2.4 | 0.62 | 0.92 | *UBE2E3* | MANE_intron_variant | 23 | 2 | 12 | 23 |
| chr16:19391115:C:T | rs140119109 | T | 0.024 | -0.91 | 0.17 | 1.5E-07 | -4.3 | 0.90 | 0.97 | *TMC5* | non_coding_transcript_variant,intron_variant,downstream_gene_variant | 19 | 1 | 0 | 5 |
| chr7:139570283:G:A | rs112212342 | A | 0.063 | -0.57 | 0.11 | 4.2E-07 | -2.2 | 0.59 | 0.91 | *HIPK2* | MANE_3_prime_untranslated_region_variant | 48 | 5 | 2 | 12 |
| **FAME Primary EUR** | | | | | | | | | | | | | | | |
| chr7:4108958:C:G | rs2342502 | G | 0.496 | -0.30 | 0.06 | 4.5E-07 | -1.6 | 0.30 | 0.97 | *SDK1* | MANE_intron_variant | 369 | - | - | - |

*Minor allele counts (MACs) were calculated using imputed genotype dosages.

**Supplemental Table 11. Top associated loci from meta-analysis from Fluocinolone Acetonide in Diabetic Macular Edema and Mass Eye and Ear/Retina Health Center (FAME-MEE/RHC) samples across ALL and EUR ancestry with a primary outcome (Max intraocular pressure rise) (P<10^-5^). T**he table shows the summary statistics for linkage disequilibrium (LD)-independent SNPs (LD clumping at r^2^ > 0.1 in 500 kb sliding window) that passed P<5X10^-7^. Listed are the Effect Allele, Effect size (Effect) for rank inverse normal transformed Max IOP rise phenotype, Standard Error (SE), and P-value, imputed genotype quality (R^2^, Info Score). The table also shows effect size (Raw Beta) and standard errors (Raw SE) for raw Max IOP rise phenotype, and minor allele counts (MAC) for four ancestry, European (EUR), Admixed American (AMR), African American (AFR), and South Asians (SAS). Minor allele counts were calculated using imputed genotype dosages. The nearest gene and most deleterious consequences were annotated using Ensembl’s Variant Effect Predictor (VEP). Column Direction represents the direction of effect from two studies in the meta-analysis, ‘+’ indicates positive effect size and ‘-’ indicating negative effect size of the SNP and column Het P-value represents heterogeneity P-value.

| Variant ID (hg38) | rsID | Effect Allele | Effect | Standard Error (SE) | P-value | Raw Beta | RawSE | Direction | Het P-value | R^2^ (InfoScore) | Nearest Gene | Deleterious Consequences | EUR minor allele count (MAC)* | AMR MAC* | AFR MAC* | SAS MAC* |
| --- | --- | --- | --- | --- | --- | --- | --- | --- | --- | --- | --- | --- | --- | --- | --- | --- |
| **FAME-MEE/RHC meta-analysis Primary ALL** | | | | | | | | | | | | | | | | |
| chr2:181021297:G:A | rs1040227 | a | -0.52 | 0.09 | 1.2E-08 | -2.3 | 0.89 | -- | 0.08 | 0.91 | *UBE2E3* | MANE_intron_variant | 53 | 5 | 27 | 23 |
| chr18:11313797:C:A | rs118113189 | a | -0.78 | 0.15 | 4.1E-07 | -4.3 | 1.38 | -- | 0.26 | 0.70 | *PIEZO2* | intergenic_variant | 39 | 1 | 0 | 4 |
| **FAME-MEE/RHC meta-analysis Primary EUR** | | | | | | | | | | | | | | | | |
| chr1:38092698:T:C | rs12734080 | t | 0.87 | 0.17 | 1.3E-07 | 4.4 | 0.94 | ++ | 0.66 | 0.86 | *POU3F1* | downstream_gene_variant, non_coding_transcript_variant, intron_variant, regulatory_region_variant | 30 | - | - | - |
| chr17:8838263:C:T | rs12952302 | t | -0.73 | 0.15 | 4.9E-07 | -3.5 | 0.82 | -- | 0.16 | 0.88 | *PIK3R6* | MANE_intron_variant | 37 | - | - | - |
| **FAME-MEE/RHC Injections-only meta-analysis Primary ALL** | | | | | | | | | | | | | | | | |
| chr2:181021297:G:A | rs1040227 | a | -0.51 | 0.09 | 6.1E-08 | -2.1 | 0.52 | -- | 0.05 | 0.91 | *UBE2E3* | MANE_intron_variant | 48 | 4 | 19 | 23 |
| chr8:72086409:C:T | rs34846538 | t | -0.38 | 0.07 | 1.8E-07 | -1.7 | 0.40 | -- | 0.93 | 0.98 | *TRPA1* | intron_variant, non_coding_transcript_variant | 104 | 14 | 23 | 9 |
| chr17:33095158:C:T | rs3928996 | t | 0.20 | 0.04 | 2.0E-07 | 1.0 | 0.21 | ++ | 0.96 | 0.98 | *SPACA3* | MANE_intron_variant | 612 | 57 | 52 | 80 |

*MACs were calculated using imputed genotype dosages

Abbreviations: SE-standard error, MAC-minor allele count, MANE-Matched Annotation from NCBI and EMBL-EBI

**Supplemental Table 12. Top genome-wide association study loci for Mass Eye and Ear/ Retina Health Center (MEE/RHC) samples across ALL and EUR ancestry with primary outcome (Max intraocular pressure rise) (P<10^-5^).** The table shows the summary statistics for linkage disequilibrium (LD)-independent variants (LD clumping at r^2^ > 0.1 in 500kb sliding window) that passed P<1x10^-5^. Listed are the Effect Allele (A1), Effect Allele’s (A1) frequency, effect size of the A1 allele (Beta) for the rank inverse normal transformed (RINT) maximal intraocular pressure (IOP) rise phenotype, Standard Error (SE), P-value, and genotype imputation quality (R^2^, INFO score). The table also shows effect size (Raw Beta) and standard errors (Raw SE) for raw (pre-transformed) maximal IOP rise at the original mmHg change per month units, and minor allele counts (MAC) for four ancestral groups: European (EUR), Admixed American (AMR), African American (AFR), and South Asian (SAS). The nearest gene and most deleterious consequences were annotated using Ensembl’s Variant Effect Predictor (VEP) and the Matched Annotation from NCBI and EMBL-EBI (MANE) transcripts.

| Variant ID (hg38) | rsID | Effect Allele (A1) | A1  Frequency | Beta | Standard Error (SE) | P-value | Raw Beta | Raw SE | R^2^ | Nearest Gene | Deleterious Consequences | EUR minor allele count (MAC)* | AMRMAC* | AFR MAC* |
| --- | --- | --- | --- | --- | --- | --- | --- | --- | --- | --- | --- | --- | --- | --- |
| **MEE/RHC Primary ALL (N=577)** | | | | | | | | | | | | | | |
| chr6:39168761:C:T | rs9470989 | T | 0.010 | -1.64 | 0.27 | 1.4E-09 | -9.8 | 1.69 | 0.98 | *KCNK5* | intergenic_variant | 0 | 1 | 11 |
| chr12:71680955:G:T | rs10879312 | T | 0.116 | 0.46 | 0.08 | 2.8E-08 | 2.8 | 0.51 | 0.95 | *TMEM19* | MANE_downstream_gene_variant | 92 | 14 | 28 |
| chr2:56602923:C:T | rs112926739 | T | 0.016 | -1.10 | 0.21 | 1.4E-07 | -6.0 | 1.31 | 0.97 | *CCDC85A* | intergenic_variant | 0 | 3 | 16 |
| chr22:18054275:G:A | rs9605514 | A | 0.046 | -0.66 | 0.13 | 2.8E-07 | -3.5 | 0.81 | 0.89 | *PEX26* | regulatory_region_variant,intergenic_variant | 38 | 2 | 13 |
| chr12:109140921:G:A | rs115670846 | A | 0.014 | -1.11 | 0.22 | 4.2E-07 | -6.2 | 1.37 | 0.95 | *ACACB* | MANE_intron_variant | 2 | 0 | 14 |
| chr18:11287802:C:T | rs117698025 | T | 0.018 | -1.14 | 0.22 | 4.7E-07 | -5.5 | 1.42 | 0.71 | *PIEZO2* | intergenic_variant | 20 | 1 | 0 |
| **MEE/RHC Primary EUR (N=495)** | | | | | | | | | | | | | | |
| chr18:11287802:C:T | rs117698025 | T | 0.02 | -1.17 | 0.22 | 2.7E-07 | -5.7 | 1.38 | 0.71 | *PIEZO2* | intergenic_variant | 20 | - | - |
| **MEE/RHC injections-only Primary ALL (N=437)** | | | | | | | | | | | | | | |
| chr17:71963822:T:C | rs7214582 | C | 0.953 | 0.76 | 0.14 | 7.1E-08 | 3.5 | 0.80 | 0.89 | *SOX9* | intergenic_variant | 40 | 1 | 0 |
| chr7:13785499:C:T | rs28451894 | T | 0.119 | -0.48 | 0.09 | 3.1E-07 | -2.3 | 0.53 | 0.86 | *ETV1* | intergenic_variant | 93 | 6 | 5 |
| **MEE/RHC injections-only Primary EUR (N=385)** | | | | | | | | | | | | | | |
| chr17:71963822:T:C | rs7214582 | C | 0.95 | 0.78 | 0.14 | 3.1E-08 | 3.6 | 0.79 | 0.89 | *SOX9* | intergenic_variant | 40 | - | - |
| chr7:13785499:C:T | rs28451894 | T | 0.12 | -0.52 | 0.09 | 7.4E-08 | -2.4 | 0.54 | 0.86 | *ETV1* | intergenic_variant | 93 | - | - |
| chr1:82306279:G:A | rs12741991 | A | 0.02 | -1.29 | 0.24 | 1.1E-07 | -6.1 | 1.37 | 1.00 | *ADGRL2* | non_coding_transcript_variant,intron_variant | 12 | - | - |
| chr10:43803196:G:A | rs12357048 | A | 0.22 | 0.38 | 0.07 | 1.2E-07 | 2.0 | 0.40 | 1.00 | *ZNF32* | non_coding_transcript_variant,intron_variant | 167 | - | - |

* Minor allele counts (MACs) were calculated using imputed genotype dosages.

**Supplemental Table 13. eCAVIAR colocalization results for genome-wide significant and subthreshold loci from genome-wide association study (GWAS) for discovery cohort Fluocinolone Acetonide in Diabetic Macular Edema (FAME).** This table contains the significant eCAVIAR colocalization results from genome-wide significant and subthreshold GWAS variants by GWAS traits/analysis, target gene and tissue combinations tested across 49 Genotype-Tissue Expression (GTEx) tissues and peripheral retina. Significance was determined at a colocalization posterior probability (CLPP) > 0.01. The colocalization e/sVariant with the highest CLPP is provided in column e/sQTL variant ID and its e/sQTL p-value and GWAS p-value are provided in columns e/sQTL p-value nominal and e/sQTL GWAS p-value. To eliminate potential false positives, only cases where the GWAS p-value of colocalization e/sVariant was below or equal to 0.05 or whose e/sQTL p-values was below or equal to 10^-4^ and/or did pass false discovery rate (FDR) below 0.05 are shown. All e/sVariant within a linkage disequilibrium (LD) interval (r^2^>0.1 plus 50 kb on either side) around each lead GWAS variant (column: GWAS variant tested (hg38)) were tested in the colocalization analysis.

| GWAS Trait | GWAS variant tested (hg38) | Effect allele | GWAS Effect | GWAS OR | GWAS P | # variants tested in LD interval* | eQTL variant ID | Gene  ENS ID | Gene  symbol | Tissue | eQTL  p-value  nominal | eQTL GWAS p-value | eQTL  slope relativeALT | eQTL SE | eQTL  GWAS beta  relative  ALT | eQTL GWAS  SE | eQTL GWAS relative effect | ECAVIAR prob in causal set | CLPP |
| --- | --- | --- | --- | --- | --- | --- | --- | --- | --- | --- | --- | --- | --- | --- | --- | --- | --- | --- | --- |
| FAME Primary ALL | chr2:181004816:G:C | C | -0.66 | 0.52 | 2.9E-08** | 1169 | chr2_180992142_C_T_b38 | ENSG00000170035.15 | *UBE2E3* | Breast Mammary Tissue | 1.6E-05 | 5.2E-06 | 0.12 | 0.026 | -0.26 | 0.056 | OPPOSITE | 0.17 | 0.059 |
| FAME Primary ALL | chr2:181004816:G:C | C | -0.66 | 0.52 | 2.9E-08** | 1169 | chr2_181029486_T_A_b38 | ENSG00000170035.15 | *UBE2E3* | Breast Mammary Tissue | 1.6E-05 | 5.2E-06 | 0.12 | 0.026 | -0.26 | 0.056 | OPPOSITE | 0.15 | 0.054 |
| FAME Primary ALL | chr2:181004816:G:C | C | -0.66 | 0.52 | 2.9E-08** | 1169 | chr2_181102348_G_A_b38 | ENSG00000170035.15 | *UBE2E3* | Breast Mammary Tissue | 1.6E-05 | 4.3E-05 | -0.12 | 0.027 | 0.23 | 0.055 | OPPOSITE | 0.24 | 0.085 |
| FAME Primary ALL | chr2:181004816:G:C | C | -0.66 | 0.52 | 2.9E-08** | 1169 | chr2_181103423_A_T_b38 | ENSG00000170035.15 | *UBE2E3* | Breast Mammary Tissue | 1.6E-05 | 6.9E-05 | -0.12 | 0.027 | 0.22 | 0.056 | OPPOSITE | 0.12 | 0.041 |
| FAME Primary ALL | chr2:181004816:G:C | C | -0.66 | 0.52 | 2.9E-08** | 1169 | chr2_181092766_G_A_b38 | ENSG00000170035.15 | *UBE2E3* | Breast Mammary Tissue | 1.6E-05 | 7.7E-05 | -0.12 | 0.027 | 0.22 | 0.056 | OPPOSITE | 0.10 | 0.035 |
| FAME Primary ALL | chr2:181004816:G:C | C | -0.66 | 0.52 | 2.9E-08** | 1169 | chr2_181086700_C_G_b38 | ENSG00000170035.15 | *UBE2E3* | Breast Mammary Tissue | 1.6E-05 | 7.8E-05 | -0.12 | 0.027 | 0.22 | 0.056 | OPPOSITE | 0.09 | 0.031 |
| FAME Primary ALL | chr2:181004816:G:C | C | -0.66 | 0.52 | 2.9E-08** | 1169 | chr2_180968140_C_T_b38 | ENSG00000170035.15 | *UBE2E3* | Breast Mammary Tissue | 6.5E-06 | 7.9E-06 | 0.12 | 0.026 | -0.25 | 0.056 | OPPOSITE | 0.08 | 0.027 |
| FAME Primary ALL | chr2:181004816:G:C | C | -0.66 | 0.52 | 2.9E-08** | 1169 | chr2_180992142_C_T_b38 | ENSG00000170035.15 | *UBE2E3* | Minor Salivary Gland | 4.3E-08 | 5.2E-06 | 0.30 | 0.052 | -0.26 | 0.056 | OPPOSITE | 0.22 | 0.098 |
| FAME Primary ALL | chr2:181004816:G:C | C | -0.66 | 0.52 | 2.9E-08** | 1169 | chr2_181029486_T_A_b38 | ENSG00000170035.15 | *UBE2E3* | Minor Salivary Gland | 4.3E-08 | 5.2E-06 | 0.30 | 0.052 | -0.26 | 0.056 | OPPOSITE | 0.19 | 0.085 |
| FAME Primary ALL | chr2:181004816:G:C | C | -0.66 | 0.52 | 2.9E-08** | 1169 | chr2_181102348_G_A_b38 | ENSG00000170035.15 | *UBE2E3* | Minor Salivary Gland | 1.2E-07 | 4.3E-05 | -0.30 | 0.053 | 0.23 | 0.055 | OPPOSITE | 0.18 | 0.083 |
| FAME Primary ALL | chr2:181004816:G:C | C | -0.66 | 0.52 | 2.9E-08** | 1169 | chr2_181024335_G_C_b38 | ENSG00000170035.15 | *UBE2E3* | Minor Salivary Gland | 4.3E-08 | 5.3E-06 | 0.30 | 0.052 | -0.26 | 0.056 | OPPOSITE | 0.13 | 0.060 |
| FAME Primary ALL | chr2:181004816:G:C | C | -0.66 | 0.52 | 2.9E-08** | 1169 | chr2_181103423_A_T_b38 | ENSG00000170035.15 | *UBE2E3* | Minor Salivary Gland | 1.2E-07 | 6.9E-05 | -0.30 | 0.053 | 0.22 | 0.056 | OPPOSITE | 0.09 | 0.040 |
| FAME Primary ALL | chr2:181004816:G:C | C | -0.66 | 0.52 | 2.9E-08** | 1169 | chr2_181092766_G_A_b38 | ENSG00000170035.15 | *UBE2E3* | Minor Salivary Gland | 1.2E-07 | 7.7E-05 | -0.30 | 0.053 | 0.22 | 0.056 | OPPOSITE | 0.07 | 0.033 |
| FAME Primary ALL | chr2:181004816:G:C | C | -0.66 | 0.52 | 2.9E-08** | 1169 | chr2_181086700_C_G_b38 | ENSG00000170035.15 | *UBE2E3* | Minor Salivary Gland | 1.2E-07 | 7.8E-05 | -0.30 | 0.053 | 0.22 | 0.056 | OPPOSITE | 0.07 | 0.033 |
| FAME Primary ALL | chr2:181004816:G:C | C | -0.66 | 0.52 | 2.9E-08** | 1169 | chr2_181018152_G_A_b38 | ENSG00000170035.15 | *UBE2E3* | Minor Salivary Gland | 5.5E-08 | 9.4E-06 | 0.30 | 0.051 | -0.25 | 0.056 | OPPOSITE | 0.02 | 0.011 |
| FAME Primary ALL | chr2:181004816:G:C | C | -0.66 | 0.52 | 2.9E-08** | 1169 | chr2_180992142_C_T_b38 | ENSG00000170035.15 | *UBE2E3* | Pancreas | 8.5E-06 | 5.2E-06 | 0.18 | 0.040 | -0.26 | 0.056 | OPPOSITE | 0.10 | 0.030 |
| FAME Primary ALL | chr2:181004816:G:C | C | -0.66 | 0.52 | 2.9E-08** | 1169 | chr2_181029486_T_A_b38 | ENSG00000170035.15 | *UBE2E3* | Pancreas | 8.0E-06 | 5.2E-06 | 0.18 | 0.040 | -0.26 | 0.056 | OPPOSITE | 0.10 | 0.030 |
| FAME Primary ALL | chr2:181004816:G:C | C | -0.66 | 0.52 | 2.9E-08** | 1169 | chr2_181102348_G_A_b38 | ENSG00000170035.15 | *UBE2E3* | Pancreas | 4.5E-06 | 4.3E-05 | -0.19 | 0.040 | 0.23 | 0.055 | OPPOSITE | 0.24 | 0.075 |
| FAME Primary ALL | chr2:181004816:G:C | C | -0.66 | 0.52 | 2.9E-08** | 1169 | chr2_181103423_A_T_b38 | ENSG00000170035.15 | *UBE2E3* | Pancreas | 4.5E-06 | 6.9E-05 | -0.19 | 0.040 | 0.22 | 0.056 | OPPOSITE | 0.12 | 0.036 |
| FAME Primary ALL | chr2:181004816:G:C | C | -0.66 | 0.52 | 2.9E-08** | 1169 | chr2_181092766_G_A_b38 | ENSG00000170035.15 | *UBE2E3* | Pancreas | 4.5E-06 | 7.7E-05 | -0.19 | 0.040 | 0.22 | 0.056 | OPPOSITE | 0.10 | 0.030 |
| FAME Primary ALL | chr2:181004816:G:C | C | -0.66 | 0.52 | 2.9E-08** | 1169 | chr2_181086700_C_G_b38 | ENSG00000170035.15 | *UBE2E3* | Pancreas | 4.3E-06 | 7.8E-05 | -0.19 | 0.040 | 0.22 | 0.056 | OPPOSITE | 0.10 | 0.032 |
| FAME Primary ALL | chr2:181004816:G:C | C | -0.66 | 0.52 | 2.9E-08** | 1169 | chr2_180968140_C_T_b38 | ENSG00000170035.15 | *UBE2E3* | Pancreas | 1.5E-06 | 7.9E-06 | 0.19 | 0.039 | -0.25 | 0.056 | OPPOSITE | 0.16 | 0.049 |
| FAME Primary ALL | chr2:181004816:G:C | C | -0.66 | 0.52 | 2.9E-08** | 1169 | chr2_180992142_C_T_b38 | ENSG00000170035.15 | *UBE2E3* | Artery Aorta | 2.2E-05 | 5.2E-06 | 0.12 | 0.028 | -0.26 | 0.056 | OPPOSITE | 0.19 | 0.074 |
| FAME Primary ALL | chr2:181004816:G:C | C | -0.66 | 0.52 | 2.9E-08** | 1169 | chr2_181029486_T_A_b38 | ENSG00000170035.15 | *UBE2E3* | Artery Aorta | 3.1E-05 | 5.2E-06 | 0.12 | 0.028 | -0.26 | 0.056 | OPPOSITE | 0.10 | 0.038 |
| FAME Primary ALL | chr2:181004816:G:C | C | -0.66 | 0.52 | 2.9E-08** | 1169 | chr2_181102348_G_A_b38 | ENSG00000170035.15 | *UBE2E3* | Artery Aorta | 4.0E-06 | 4.3E-05 | -0.13 | 0.028 | 0.23 | 0.055 | OPPOSITE | 0.25 | 0.098 |
| FAME Primary ALL | chr2:181004816:G:C | C | -0.66 | 0.52 | 2.9E-08** | 1169 | chr2_181024335_G_C_b38 | ENSG00000170035.15 | *UBE2E3* | Artery Aorta | 3.3E-05 | 5.3E-06 | 0.12 | 0.028 | -0.26 | 0.056 | OPPOSITE | 0.06 | 0.025 |
| FAME Primary ALL | chr2:181004816:G:C | C | -0.66 | 0.52 | 2.9E-08** | 1169 | chr2_181103423_A_T_b38 | ENSG00000170035.15 | *UBE2E3* | Artery Aorta | 4.0E-06 | 6.9E-05 | -0.13 | 0.028 | 0.22 | 0.056 | OPPOSITE | 0.12 | 0.047 |
| FAME Primary ALL | chr2:181004816:G:C | C | -0.66 | 0.52 | 2.9E-08** | 1169 | chr2_181092766_G_A_b38 | ENSG00000170035.15 | *UBE2E3* | Artery Aorta | 4.0E-06 | 7.7E-05 | -0.13 | 0.028 | 0.22 | 0.056 | OPPOSITE | 0.10 | 0.040 |
| FAME Primary ALL | chr2:181004816:G:C | C | -0.66 | 0.52 | 2.9E-08** | 1169 | chr2_181086700_C_G_b38 | ENSG00000170035.15 | *UBE2E3* | Artery Aorta | 6.4E-06 | 7.8E-05 | -0.13 | 0.028 | 0.22 | 0.056 | OPPOSITE | 0.05 | 0.019 |
| FAME Primary ALL | chr2:181004816:G:C | C | -0.66 | 0.52 | 2.9E-08** | 1169 | chr2_181018152_G_A_b38 | ENSG00000170035.15 | *UBE2E3* | Artery Aorta | 1.3E-05 | 9.4E-06 | 0.12 | 0.028 | -0.25 | 0.056 | OPPOSITE | 0.09 | 0.037 |
| FAME Primary ALL | chr2:181004816:G:C | C | -0.66 | 0.52 | 2.9E-08** | 1169 | chr2_180992142_C_T_b38 | ENSG00000170035.15 | *UBE2E3* | Esophagus Mucosa | 1.4E-08 | 5.2E-06 | 0.17 | 0.029 | -0.26 | 0.056 | OPPOSITE | 0.21 | 0.016 |
| FAME Primary ALL | chr2:181004816:G:C | C | -0.66 | 0.52 | 2.9E-08** | 1169 | chr2_181029486_T_A_b38 | ENSG00000170035.15 | *UBE2E3* | Esophagus Mucosa | 1.0E-08 | 5.2E-06 | 0.17 | 0.029 | -0.26 | 0.056 | OPPOSITE | 0.32 | 0.025 |
| FAME Primary ALL | chr2:181004816:G:C | C | -0.66 | 0.52 | 2.9E-08** | 1169 | chr2_181102348_G_A_b38 | ENSG00000170035.15 | *UBE2E3* | Esophagus Mucosa | 2.4E-08 | 4.3E-05 | -0.17 | 0.029 | 0.23 | 0.055 | OPPOSITE | 0.18 | 0.014 |
| FAME Primary ALL | chr2:181004816:G:C | C | -0.66 | 0.52 | 2.9E-08** | 1169 | chr2_180992142_C_T_b38 | ENSG00000170035.15 | *UBE2E3* | Nerve Tibial | 3.2E-06 | 5.2E-06 | 0.10 | 0.022 | -0.26 | 0.056 | OPPOSITE | 0.12 | 0.030 |
| FAME Primary ALL | chr2:181004816:G:C | C | -0.66 | 0.52 | 2.9E-08** | 1169 | chr2_181029486_T_A_b38 | ENSG00000170035.15 | *UBE2E3* | Nerve Tibial | 1.9E-06 | 5.2E-06 | 0.11 | 0.022 | -0.26 | 0.056 | OPPOSITE | 0.24 | 0.062 |
| FAME Primary ALL | chr2:181004816:G:C | C | -0.66 | 0.52 | 2.9E-08** | 1169 | chr2_181102348_G_A_b38 | ENSG00000170035.15 | *UBE2E3* | Nerve Tibial | 1.6E-06 | 4.3E-05 | -0.11 | 0.022 | 0.23 | 0.055 | OPPOSITE | 0.21 | 0.054 |
| FAME Primary ALL | chr2:181004816:G:C | C | -0.66 | 0.52 | 2.9E-08** | 1169 | chr2_181024335_G_C_b38 | ENSG00000170035.15 | *UBE2E3* | Nerve Tibial | 2.6E-06 | 5.3E-06 | 0.10 | 0.022 | -0.26 | 0.056 | OPPOSITE | 0.11 | 0.028 |
| FAME Primary ALL | chr2:181004816:G:C | C | -0.66 | 0.52 | 2.9E-08** | 1169 | chr2_181103423_A_T_b38 | ENSG00000170035.15 | *UBE2E3* | Nerve Tibial | 1.6E-06 | 6.9E-05 | -0.11 | 0.022 | 0.22 | 0.056 | OPPOSITE | 0.10 | 0.026 |
| FAME Primary ALL | chr2:181004816:G:C | C | -0.66 | 0.52 | 2.9E-08** | 1169 | chr2_181092766_G_A_b38 | ENSG00000170035.15 | *UBE2E3* | Nerve Tibial | 1.6E-06 | 7.7E-05 | -0.11 | 0.022 | 0.22 | 0.056 | OPPOSITE | 0.08 | 0.022 |
| FAME Primary ALL | chr2:181004816:G:C | C | -0.66 | 0.52 | 2.9E-08** | 1169 | chr2_181086700_C_G_b38 | ENSG00000170035.15 | *UBE2E3* | Nerve Tibial | 1.8E-06 | 7.8E-05 | -0.11 | 0.022 | 0.22 | 0.056 | OPPOSITE | 0.07 | 0.019 |
| FAME Primary ALL | chr2:181004816:G:C | C | -0.66 | 0.52 | 2.9E-08** | 1169 | chr2_180968140_C_T_b38 | ENSG00000170035.15 | *UBE2E3* | Nerve Tibial | 1.3E-06 | 7.9E-06 | 0.11 | 0.022 | -0.25 | 0.056 | OPPOSITE | 0.05 | 0.012 |
| FAME Primary ALL | chr2:181004816:G:C | C | -0.66 | 0.52 | 2.9E-08** | 1169 | chr2_180992142_C_T_b38 | ENSG00000170035.15 | *UBE2E3* | Breast Mammary Tissue | 1.6E-05 | 5.2E-06 | 0.12 | 0.026 | -0.26 | 0.056 | OPPOSITE | 0.17 | 0.059 |
| FAME Primary ALL | chr2:181004816:G:C | C | -0.66 | 0.52 | 2.9E-08** | 1169 | chr2_181029486_T_A_b38 | ENSG00000170035.15 | *UBE2E3* | Breast Mammary Tissue | 1.6E-05 | 5.2E-06 | 0.12 | 0.026 | -0.26 | 0.056 | OPPOSITE | 0.15 | 0.054 |
| FAME Primary ALL | chr2:181004816:G:C | C | -0.66 | 0.52 | 2.9E-08** | 1169 | chr2_181102348_G_A_b38 | ENSG00000170035.15 | *UBE2E3* | Breast Mammary Tissue | 1.6E-05 | 4.3E-05 | -0.12 | 0.027 | 0.23 | 0.055 | OPPOSITE | 0.24 | 0.085 |
| FAME Primary ALL | chr2:181004816:G:C | C | -0.66 | 0.52 | 2.9E-08** | 1169 | chr2_181103423_A_T_b38 | ENSG00000170035.15 | *UBE2E3* | Breast Mammary Tissue | 1.6E-05 | 6.9E-05 | -0.12 | 0.027 | 0.22 | 0.056 | OPPOSITE | 0.12 | 0.041 |
| FAME Primary ALL | chr2:181004816:G:C | C | -0.66 | 0.52 | 2.9E-08** | 1169 | chr2_181092766_G_A_b38 | ENSG00000170035.15 | *UBE2E3* | Breast Mammary Tissue | 1.6E-05 | 7.7E-05 | -0.12 | 0.027 | 0.22 | 0.056 | OPPOSITE | 0.10 | 0.035 |
| FAME Primary ALL | chr2:181004816:G:C | C | -0.66 | 0.52 | 2.9E-08** | 1169 | chr2_181086700_C_G_b38 | ENSG00000170035.15 | *UBE2E3* | Breast Mammary Tissue | 1.6E-05 | 7.8E-05 | -0.12 | 0.027 | 0.22 | 0.056 | OPPOSITE | 0.09 | 0.031 |
| FAME Primary ALL | chr2:181004816:G:C | C | -0.66 | 0.52 | 2.9E-08** | 1169 | chr2_180968140_C_T_b38 | ENSG00000170035.15 | *UBE2E3* | Breast Mammary Tissue | 6.5E-06 | 7.9E-06 | 0.12 | 0.026 | -0.25 | 0.056 | OPPOSITE | 0.08 | 0.027 |
| FAME Primary ALL | chr2:181004816:G:C | C | -0.66 | 0.52 | 2.9E-08** | 1169 | chr2_180992142_C_T_b38 | ENSG00000170035.15 | *UBE2E3* | Minor Salivary Gland | 4.3E-08 | 5.2E-06 | 0.30 | 0.052 | -0.26 | 0.056 | OPPOSITE | 0.22 | 0.098 |
| FAME Primary ALL | chr2:181004816:G:C | C | -0.66 | 0.52 | 2.9E-08** | 1169 | chr2_181029486_T_A_b38 | ENSG00000170035.15 | *UBE2E3* | Minor Salivary Gland | 4.3E-08 | 5.2E-06 | 0.30 | 0.052 | -0.26 | 0.056 | OPPOSITE | 0.19 | 0.085 |
| FAME Primary ALL | chr2:181004816:G:C | C | -0.66 | 0.52 | 2.9E-08** | 1169 | chr2_181102348_G_A_b38 | ENSG00000170035.15 | *UBE2E3* | Minor Salivary Gland | 1.2E-07 | 4.3E-05 | -0.30 | 0.053 | 0.23 | 0.055 | OPPOSITE | 0.18 | 0.083 |
| FAME Primary ALL | chr2:181004816:G:C | C | -0.66 | 0.52 | 2.9E-08** | 1169 | chr2_181024335_G_C_b38 | ENSG00000170035.15 | *UBE2E3* | Minor Salivary Gland | 4.3E-08 | 5.3E-06 | 0.30 | 0.052 | -0.26 | 0.056 | OPPOSITE | 0.13 | 0.060 |
| FAME Primary ALL | chr2:181004816:G:C | C | -0.66 | 0.52 | 2.9E-08** | 1169 | chr2_181103423_A_T_b38 | ENSG00000170035.15 | *UBE2E3* | Minor Salivary Gland | 1.2E-07 | 6.9E-05 | -0.30 | 0.053 | 0.22 | 0.056 | OPPOSITE | 0.09 | 0.040 |
| FAME Primary ALL | chr2:181004816:G:C | C | -0.66 | 0.52 | 2.9E-08** | 1169 | chr2_181092766_G_A_b38 | ENSG00000170035.15 | *UBE2E3* | Minor Salivary Gland | 1.2E-07 | 7.7E-05 | -0.30 | 0.053 | 0.22 | 0.056 | OPPOSITE | 0.07 | 0.033 |
| FAME Primary ALL | chr2:181004816:G:C | C | -0.66 | 0.52 | 2.9E-08** | 1169 | chr2_181086700_C_G_b38 | ENSG00000170035.15 | *UBE2E3* | Minor Salivary Gland | 1.2E-07 | 7.8E-05 | -0.30 | 0.053 | 0.22 | 0.056 | OPPOSITE | 0.07 | 0.033 |
| FAME Primary ALL | chr2:181004816:G:C | C | -0.66 | 0.52 | 2.9E-08** | 1169 | chr2_181018152_G_A_b38 | ENSG00000170035.15 | *UBE2E3* | Minor Salivary Gland | 5.5E-08 | 9.4E-06 | 0.30 | 0.051 | -0.25 | 0.056 | OPPOSITE | 0.02 | 0.011 |
| FAME Primary ALL | chr2:181004816:G:C | C | -0.66 | 0.52 | 2.9E-08** | 1169 | chr2_180992142_C_T_b38 | ENSG00000170035.15 | *UBE2E3* | Pancreas | 8.5E-06 | 5.2E-06 | 0.18 | 0.040 | -0.26 | 0.056 | OPPOSITE | 0.10 | 0.030 |
| FAME Primary ALL | chr2:181004816:G:C | C | -0.66 | 0.52 | 2.9E-08** | 1169 | chr2_181029486_T_A_b38 | ENSG00000170035.15 | *UBE2E3* | Pancreas | 8.0E-06 | 5.2E-06 | 0.18 | 0.040 | -0.26 | 0.056 | OPPOSITE | 0.10 | 0.030 |
| FAME Primary ALL | chr2:181004816:G:C | C | -0.66 | 0.52 | 2.9E-08** | 1169 | chr2_181102348_G_A_b38 | ENSG00000170035.15 | *UBE2E3* | Pancreas | 4.5E-06 | 4.3E-05 | -0.19 | 0.040 | 0.23 | 0.055 | OPPOSITE | 0.24 | 0.075 |
| FAME Primary ALL | chr2:181004816:G:C | C | -0.66 | 0.52 | 2.9E-08** | 1169 | chr2_181103423_A_T_b38 | ENSG00000170035.15 | *UBE2E3* | Pancreas | 4.5E-06 | 6.9E-05 | -0.19 | 0.040 | 0.22 | 0.056 | OPPOSITE | 0.12 | 0.036 |
| FAME Primary ALL | chr2:181004816:G:C | C | -0.66 | 0.52 | 2.9E-08** | 1169 | chr2_181092766_G_A_b38 | ENSG00000170035.15 | *UBE2E3* | Pancreas | 4.5E-06 | 7.7E-05 | -0.19 | 0.040 | 0.22 | 0.056 | OPPOSITE | 0.10 | 0.030 |
| FAME Primary ALL | chr2:181004816:G:C | C | -0.66 | 0.52 | 2.9E-08** | 1169 | chr2_181086700_C_G_b38 | ENSG00000170035.15 | *UBE2E3* | Pancreas | 4.3E-06 | 7.8E-05 | -0.19 | 0.040 | 0.22 | 0.056 | OPPOSITE | 0.10 | 0.032 |
| FAME Primary ALL | chr2:181004816:G:C | C | -0.66 | 0.52 | 2.9E-08** | 1169 | chr2_180968140_C_T_b38 | ENSG00000170035.15 | *UBE2E3* | Pancreas | 1.5E-06 | 7.9E-06 | 0.19 | 0.039 | -0.25 | 0.056 | OPPOSITE | 0.16 | 0.049 |
| FAME Primary ALL | chr2:181004816:G:C | C | -0.66 | 0.52 | 2.9E-08** | 1169 | chr2_180992142_C_T_b38 | ENSG00000170035.15 | *UBE2E3* | Artery Aorta | 2.2E-05 | 5.2E-06 | 0.12 | 0.028 | -0.26 | 0.056 | OPPOSITE | 0.19 | 0.074 |
| FAME Primary ALL | chr2:181004816:G:C | C | -0.66 | 0.52 | 2.9E-08** | 1169 | chr2_181029486_T_A_b38 | ENSG00000170035.15 | *UBE2E3* | Artery Aorta | 3.1E-05 | 5.2E-06 | 0.12 | 0.028 | -0.26 | 0.056 | OPPOSITE | 0.10 | 0.038 |
| FAME Primary ALL | chr2:181004816:G:C | C | -0.66 | 0.52 | 2.9E-08** | 1169 | chr2_181102348_G_A_b38 | ENSG00000170035.15 | *UBE2E3* | Artery Aorta | 4.0E-06 | 4.3E-05 | -0.13 | 0.028 | 0.23 | 0.055 | OPPOSITE | 0.25 | 0.098 |
| FAME Primary ALL | chr2:181004816:G:C | C | -0.66 | 0.52 | 2.9E-08** | 1169 | chr2_181024335_G_C_b38 | ENSG00000170035.15 | *UBE2E3* | Artery Aorta | 3.3E-05 | 5.3E-06 | 0.12 | 0.028 | -0.26 | 0.056 | OPPOSITE | 0.06 | 0.025 |
| FAME Primary ALL | chr2:181004816:G:C | C | -0.66 | 0.52 | 2.9E-08** | 1169 | chr2_181103423_A_T_b38 | ENSG00000170035.15 | *UBE2E3* | Artery Aorta | 4.0E-06 | 6.9E-05 | -0.13 | 0.028 | 0.22 | 0.056 | OPPOSITE | 0.12 | 0.047 |
| FAME Primary ALL | chr2:181004816:G:C | C | -0.66 | 0.52 | 2.9E-08** | 1169 | chr2_181092766_G_A_b38 | ENSG00000170035.15 | *UBE2E3* | Artery Aorta | 4.0E-06 | 7.7E-05 | -0.13 | 0.028 | 0.22 | 0.056 | OPPOSITE | 0.10 | 0.040 |
| FAME Primary ALL | chr2:181004816:G:C | C | -0.66 | 0.52 | 2.9E-08** | 1169 | chr2_181086700_C_G_b38 | ENSG00000170035.15 | *UBE2E3* | Artery Aorta | 6.4E-06 | 7.8E-05 | -0.13 | 0.028 | 0.22 | 0.056 | OPPOSITE | 0.05 | 0.019 |
| FAME Primary ALL | chr2:181004816:G:C | C | -0.66 | 0.52 | 2.9E-08** | 1169 | chr2_181018152_G_A_b38 | ENSG00000170035.15 | *UBE2E3* | Artery Aorta | 1.3E-05 | 9.4E-06 | 0.12 | 0.028 | -0.25 | 0.056 | OPPOSITE | 0.09 | 0.037 |
| FAME Primary ALL | chr2:181004816:G:C | C | -0.66 | 0.52 | 2.9E-08** | 1169 | chr2_180992142_C_T_b38 | ENSG00000170035.15 | *UBE2E3* | Esophagus Mucosa | 1.4E-08 | 5.2E-06 | 0.17 | 0.029 | -0.26 | 0.056 | OPPOSITE | 0.21 | 0.016 |
| FAME Primary ALL | chr2:181004816:G:C | C | -0.66 | 0.52 | 2.9E-08** | 1169 | chr2_181029486_T_A_b38 | ENSG00000170035.15 | *UBE2E3* | Esophagus Mucosa | 1.0E-08 | 5.2E-06 | 0.17 | 0.029 | -0.26 | 0.056 | OPPOSITE | 0.32 | 0.025 |
| FAME Primary ALL | chr2:181004816:G:C | C | -0.66 | 0.52 | 2.9E-08** | 1169 | chr2_181102348_G_A_b38 | ENSG00000170035.15 | *UBE2E3* | Esophagus Mucosa | 2.4E-08 | 4.3E-05 | -0.17 | 0.029 | 0.23 | 0.055 | OPPOSITE | 0.18 | 0.014 |
| FAME Primary ALL | chr2:181004816:G:C | C | -0.66 | 0.52 | 2.9E-08** | 1169 | chr2_180992142_C_T_b38 | ENSG00000170035.15 | *UBE2E3* | Nerve Tibial | 3.2E-06 | 5.2E-06 | 0.10 | 0.022 | -0.26 | 0.056 | OPPOSITE | 0.12 | 0.030 |
| FAME Primary ALL | chr2:181004816:G:C | C | -0.66 | 0.52 | 2.9E-08** | 1169 | chr2_181029486_T_A_b38 | ENSG00000170035.15 | *UBE2E3* | Nerve Tibial | 1.9E-06 | 5.2E-06 | 0.11 | 0.022 | -0.26 | 0.056 | OPPOSITE | 0.24 | 0.062 |
| FAME Primary ALL | chr2:181004816:G:C | C | -0.66 | 0.52 | 2.9E-08** | 1169 | chr2_181102348_G_A_b38 | ENSG00000170035.15 | *UBE2E3* | Nerve Tibial | 1.6E-06 | 4.3E-05 | -0.11 | 0.022 | 0.23 | 0.055 | OPPOSITE | 0.21 | 0.054 |
| FAME Primary ALL | chr2:181004816:G:C | C | -0.66 | 0.52 | 2.9E-08** | 1169 | chr2_181024335_G_C_b38 | ENSG00000170035.15 | *UBE2E3* | Nerve Tibial | 2.6E-06 | 5.3E-06 | 0.10 | 0.022 | -0.26 | 0.056 | OPPOSITE | 0.11 | 0.028 |
| FAME Primary ALL | chr2:181004816:G:C | C | -0.66 | 0.52 | 2.9E-08** | 1169 | chr2_181103423_A_T_b38 | ENSG00000170035.15 | *UBE2E3* | Nerve Tibial | 1.6E-06 | 6.9E-05 | -0.11 | 0.022 | 0.22 | 0.056 | OPPOSITE | 0.10 | 0.026 |
| FAME Primary ALL | chr2:181004816:G:C | C | -0.66 | 0.52 | 2.9E-08** | 1169 | chr2_181092766_G_A_b38 | ENSG00000170035.15 | *UBE2E3* | Nerve Tibial | 1.6E-06 | 7.7E-05 | -0.11 | 0.022 | 0.22 | 0.056 | OPPOSITE | 0.08 | 0.022 |
| FAME Primary ALL | chr2:181004816:G:C | C | -0.66 | 0.52 | 2.9E-08** | 1169 | chr2_181086700_C_G_b38 | ENSG00000170035.15 | *UBE2E3* | Nerve Tibial | 1.8E-06 | 7.8E-05 | -0.11 | 0.022 | 0.22 | 0.056 | OPPOSITE | 0.07 | 0.019 |
| FAME Primary ALL | chr2:181004816:G:C | C | -0.66 | 0.52 | 2.9E-08** | 1169 | chr2_180968140_C_T_b38 | ENSG00000170035.15 | *UBE2E3* | Nerve Tibial | 1.3E-06 | 7.9E-06 | 0.11 | 0.022 | -0.25 | 0.056 | OPPOSITE | 0.05 | 0.012 |
| FAME Primary EUR | chr7:4108958:C:G | G | -0.30 | 0.74 | 4.5E-07 | 6679 | chr7_4111282_T_C_b38 | ENSG00000283991.1 | *RP11-42B7.1* | Liver | 6.2E-06 | 1.1E-06 | 0.38 | 0.082 | -0.29 | 0.057 | OPPOSITE | 0.08 | 0.013 |
| FAME Primary EUR | chr7:4108958:C:G | G | -0.30 | 0.74 | 4.5E-07 | 6679 | chr7_4131845_CAT_C_b38 | ENSG00000283991.1 | *RP11-42B7.1* | Liver | 5.4E-06 | 7.2E-05 | -0.38 | 0.081 | 0.24 | 0.059 | OPPOSITE | 0.53 | 0.082 |
| FAME Primary EUR | chr7:4108958:C:G | G | -0.30 | 0.74 | 4.5E-07 | 6679 | chr7_4134757_G_C_b38 | ENSG00000283991.1 | *RP11-42B7.1* | Liver | 5.4E-06 | 6.4E-05 | -0.38 | 0.081 | 0.24 | 0.059 | OPPOSITE | 0.36 | 0.055 |
| FAME Primary EUR | chr7:4108958:C:G | G | -0.30 | 0.74 | 4.5E-07 | 6680 | chr7_4111282_T_C_b38 | ENSG00000283991.1 | *RP11-42B7.1* | Ovary | 1.1E-06 | 1.1E-06 | 0.41 | 0.080 | -0.29 | 0.057 | OPPOSITE | 0.53 | 0.550 |
| FAME Primary EUR | chr7:4108958:C:G | G | -0.30 | 0.74 | 4.5E-07 | 6680 | chr7_4131845_CAT_C_b38 | ENSG00000283991.1 | *RP11-42B7.1* | Ovary | 7.8E-09 | 7.2E-05 | -0.49 | 0.079 | 0.24 | 0.059 | OPPOSITE | 0.41 | 0.425 |
| FAME Primary EUR | chr7:4108958:C:G | G | -0.30 | 0.74 | 4.5E-07 | 6680 | chr7_4134757_G_C_b38 | ENSG00000283991.1 | *RP11-42B7.1* | Ovary | 1.4E-08 | 6.4E-05 | -0.48 | 0.079 | 0.24 | 0.059 | OPPOSITE | 0.06 | 0.062 |
| FAME Primary ALL | chr16:19391115:C:T | G | -0.90 | 0.4 | 1.5e-07 | 26 | chr16_19442036_G_C_b38 | ENSG00000259925 | *CTA-363E6.2* | Retina | 1.85E-19 | 0.047 | -0.50 | 0.054 | 0.13 | 0.067 | OPPOSITE | 0.88 | 0.04 |
| FAME Primary ALL | chr16:19391115:C:T | G | -0.90 | 0.4 | 1.5e-07 | 26 | chr16_19442036_G_C_b38 | ENSG00000280265 | *CTA-363E6.3* | Retina | 2.48E-20 | 0.047 | -0.49 | 0.050 | 0.13 | 0.067 | OPPOSITE | 0.88 | 0.04 |

* r2 0.1 ± 50kb

**P passes the genome-wide significant threshold (P<5E-08)

Abbreviations: FAME-Fluocinolone Acetonide in Diabetic Macular Edema, GWAS-genome-wide association study, GTEx-Genotype-Tissue Expression, CLPP- colocalization posterior probability, eQTL-expression quantitative trait loci, sQTL- splicing Quantitative Trait Loci, FDR-false discovery rate, LD-linkage disequilibrium, OR-odds ratio, SE-standard error, Gene ENS ID- Ensembl gene ID, EUR-European

**Supplemental Table 14. eCAVIAR colocalization results for genome-wide significant and subthreshold loci from the Fluocinolone Acetonide in Diabetic Macular Edema (FAME)-Mass Eye and Ear/Retina Health Center (MEE/RHC) meta-analysis and the loci replicated in both cohorts.** This table contains the significant eCAVIAR colocalization results from genome-wide significant and sub-threshold GWAS variants by trait/analysis, target gene and tissue combinations tested across 49 GTEx tissues and peripheral retina. Significance was determined at a colocalization posterior probability (CLPP) > 0.01. The colocalization e/sVariant with the highest CLPP is provided in column e/sQTL variant ID and its e/sQTL p-value and GWAS p-value are provided in columns e/sQTL p-value nominal and e/sQTL GWAS p-value. To eliminate potential false positives, only cases where the GWAS p-value of colocalization e/sVariant was below or equal to 0.05 or whose e/sQTL p-values was below or equal to 10^-4^ and/or did passed FDR below 0.05 are shown. All e/sVariant within an LD interval (r^2^>0.1 plus 50 kb on either side) around each lead GWAS variant (column: GWAS variant tested (hg38)) were tested in the colocalization analysis.

| Meta-analysis Trait | GWAS variant tested | Effect allele | GWAS EFFECT | GWAS OR | GWAS P | # variants tested in LD interval* | e/sQTL Variant ID | Gene ENS ID | Gene symbol | Tissue | e/sQTL  p-value nominal | e/sQTL GWAS  p-value | e/sQTL slope relative ALT | e/sQTL SE | e/sQTL GWAS beta relative ALT | e/sQTL GWAS SE | e/sQTL GWAS relative effect | eCAVIAR prob in causal set | CLPP |
| --- | --- | --- | --- | --- | --- | --- | --- | --- | --- | --- | --- | --- | --- | --- | --- | --- | --- | --- | --- |
| FAME-MEE Primary ALL | chr2:181021297:G:A | a | -0.52 | 0.59 | 1.23E-08** | 1119 | chr2:181021297:G:A | ENSG00000138434.16 | *SSFA2* | Nerve_Tibial | 4.40E-05 | 1.23E-08 | -0.17 | 0.04 | -0.52 | 0.09 | SAME | 0.44 | 0.012 |
| FAME-MEE Primary ALL | chr2:181021296:A:C | a | -0.51 | 1.67 | 1.3E-08** | 1119 | chr2:181021297:G:A | ENSG00000138434.16 | *SSFA2* | Nerve_Tibial | 4.40E-05 | 1.23E-08 | -0.17 | 0.04 | -0.52 | 0.09 | SAME | 0.44 | 0.012 |
| FAME-MEE/RHC Injections Primary ALL | chr17:33095158:C:T | t | 0.20 | 1.22 | 2.04E-07 | 390 | chr17:33093712:C:T | ENSG00000280020.1 | *CTD-2095E4.4* | Artery_Tibial | 6.64E-05 | 2.29E-07 | 0.20 | 0.05 | 0.20 | 0.04 | SAME | 0.10 | 0.04 |
| FAME-MEE/RHC Injections Primary ALL | chr17:33095158:C:T | t | 0.20 | 1.22 | 2.04E-07 | 390 | chr17:33095158:C:T | ENSG00000280020.1 | *CTD-2095E4.4* | Artery_Tibial | 8.79E-05 | 2.04E-07 | 0.20 | 0.05 | 0.20 | 0.04 | SAME | 0.10 | 0.04 |
| FAME-MEE/RHC Injections Primary ALL | chr17:33095158:C:T | t | 0.20 | 1.22 | 2.04E-07 | 390 | chr17:33083436:C:T | ENSG00000280020.1 | *CTD-2095E4.4* | Artery_Tibial | 5.38E-05 | 2.31E-07 | 0.20 | 0.05 | 0.20 | 0.04 | SAME | 0.06 | 0.02 |
| FAME-MEE/RHC Injections Primary ALL | chr17:33095158:C:T | t | 0.20 | 1.22 | 2.04E-07 | 390 | chr17:33084797:A:C | ENSG00000280020.1 | *CTD-2095E4.4* | Artery_Tibial | 4.75E-05 | 3.13E-07 | 0.20 | 0.05 | -0.20 | 0.04 | OPPOSITE | 0.03 | 0.01 |
| FAME-MEE/RHC Injections Primary ALL | chr2:181021297:G:A | a | -0.51 | 0.60 | 6.08E-08 | 1118 | chr2:181021297:G:A | ENSG00000138434.16 | *SSFA2* | Nerve_Tibial | 4.40E-05 | 6.08E-08 | -0.17 | 0.04 | -0.51 | 0.09 | SAME | 0.45 | 0.012 |
| FAME-MEE/RHC Injections Primary ALL | chr2:181021297:G:A | a | -0.51 | 0.60 | 6.08E-08 | 1118 | chr2:181019696:A:C | ENSG00000138434.16 | *SSFA2* | Nerve_Tibial | 5.98E-05 | 6.16E-08 | -0.16 | 0.04 | 0.51 | 0.09 | OPPOSITE | 0.36 | 0.01 |
| FAME-MEE Primary ALL | chr2:181093334:C:T | t | -0.45 | 0.63 | 1.14E-06 | 1119 | chr2:181021297:G:A | ENSG00000138434.16 | *SSFA2* | Nerve_Tibial | 4.40E-05 | 1.23E-08 | -0.17 | 0.04 | -0.52 | 0.09 | SAME | 0.44 | 0.012 |
| FAME-MEE Primary EUR | chr17:8838263:C:T | t | -0.73 | 0.48 | 4.89E-07 | 35 | chr17: 8080839:A:C (eQTL) | ENSG00000132518 | *GUCY2D* | Retina | 5.6E-05 | 0.017 | 0.057 | 0.01 | -0.14 | 0.06 | OPPOSITE | 0.59 | 0.04 |
| FAME-MEE Primary EUR | chr17:8838263:C:T | t | -0.73 | 0.48 | 4.89E-07 | 436 | chr17:8851364:T:C (eQTL) | ENSG00000276231.4 | *PIK3R6* | Testis | 8.62E-18 | 0.013 | 0.40 | 0.04 | 0.11 | 0.04 | SAME | 0.92 | 0.02 |
| FAME-MEE Primary EUR | chr17:8838263:C:T | t | -0.73 | 0.48 | 4.89E-07 | 436 | chr17_8778482:G:A | ENSG00000185156.5 | *MFSD6L* | Pituitary | 2.30E-06 | 0.029 | 0.23 | 0.05 | -0.09 | 0.04 | OPPOSITE | 0.95 | 0.02 |
| FAME-MEE Primary EUR | chr17:8838263:C:T | t | -0.73 | 0.48 | 4.89E-07 | 436 | chr17_8851364:T:C (sQTL) | ENSG00000276231.4 | *PIK3R6* | Testis | 2.29E-06 | 0.013 | -0.38 | 0.08 | 0.11 | 0.04 | OPPOSITE | 0.91 | 0.02 |

* r2 > 0.1 ± 50kb

** P passes the genome-wide significant threshold (P<5E-08)

Abbreviations: FAME-Fluocinolone Acetonide in Diabetic Macular Edema, MEE/RHC-Mass Eye and Ear/Retina Health Center, GWAS-genome-wide association study, GTEx-Genotype-Tissue Expression, CLPP- colocalization posterior probability, eQTL-expression quantitative trait loci, sQTL- splicing Quantitative Trait Loci, FDR-false discovery rate, LD-linkage disequilibrium, OR-odds ratio, SE-standard error, Gene ENS ID- Ensembl gene ID, EUR-European

**Supplemental Table 15. Biological gene sets significantly enriched for glucocorticoid-induced intraocular pressure (IOP) gene associations using MAGMA for discovery cohort, Fluocinolone Acetonide in Diabetic Macular Edema (FAME)**. The table shows significantly enriched gene-sets or pathways from four databases downloaded from MSigDB using MAGMA. The databases include Gene Ontology (GO) with three domains: biological processes (BP), molecular function (MF), and cellular components (CC), Reactome, HALLMARK, and Kyoto Encyclopedia of Genes and Genomes (KEGG) shown in column ‘Resource’. Only Gene-sets that meet the Bonferroni-corrected p-value threshold (P ≤ Alpha/Number of tests per resource; Alpha=0.05) are reported. The number of independent gene sets tested per resource are shown in column ‘Number of Tests’. The number and name of the top genes with P<0.05 within the set are shown in column ‘Top Genes in Set (P<0.05)’.

| Resource | Gene-set/Pathway | # Genes in Set | Set P-Value | # Significant Genes in Set P≤ 0.05 | Top Genes in Set (P<0.05) | log10(P) | Set | Alpha | Number of Tests |
| --- | --- | --- | --- | --- | --- | --- | --- | --- | --- |
| **FAME ALL Extreme Responders** | | | | | | | | | |
| GO_BP | POSTSYNAPTIC_MODULATION_OF_CHEMICAL_SYNAPTIC_TRANSMISSION | 14 | 2.23E-08 | 1 | *CDH1* | 7.65 | SET1 | 0.05 | 7472 |
| REACTOME | SYNTHESIS_OF_PROSTAGLANDINS_PG_AND_THROMBOXANES_TX_ | 14 | 2.64E-05 | 1 | *AKR1C3* | 4.58 | SET1 | 0.05 | 1553 |
| **FAME EUR Max IOP Rise** | | | | | | | | | |
| GO_BP | PROTEIN_SIDE_CHAIN_DEGLUTAMYLATION | 5 | 2.23E-07 | 1 | *AGBL4* | 6.65 | SET1 | 0.05 | 7472 |
| GO_BP | PROTEIN_DEGLUTAMYLATION | 7 | 4.68E-06 | 1 | *AGBL4* | 5.33 | SET2 | 0.05 | 7472 |
| **FAME ALL Responders** | | | | | | | | | |
| GO_CC | SPERM_HEAD_PLASMA_MEMBRANE | 6 | 1.97E-06 | 1 | *RHO* | 5.71 | SET1 | 0.05 | 989 |
| GO_BP | POSITIVE_REGULATION_OF_VIRAL_GENOME_REPLICATION | 31 | 3.04E-06 | 4 | *TOP2B,CCL5,PPIE,NR5A2* | 5.52 | SET1 | 0.05 | 7472 |
| GO_MF | RNA_POLYMERASE_ACTIVITY | 37 | 8.07E-06 | 2 | *POLR2L,TRNT1* | 5.09 | SET1 | 0.05 | 1705 |
| REACTOME | CHK1_CHK2_CDS1_MEDIATED_INACTIVATION_OF_CYCLIN_B_CDK1_COMPLEX | 13 | 1.13E-05 | 1 | *YWHAZ* | 4.95 | SET2 | 0.05 | 1553 |
| GO_MF | NUCLEOTIDYLTRANSFERASE_ACTIVITY | 107 | 2.61E-05 | 5 | *YRDC,TRNT1,CDS1,POLR2L,PAPOLA* | 4.58 | SET2 | 0.05 | 1705 |
| GO_CC | SPERM_HEAD | 16 | 3.05E-05 | 2 | *DDX6,RHO* | 4.52 | SET2 | 0.05 | 989 |
| KEGG | ABC_TRANSPORTERS | 44 | 8.61E-05 | 1 | *CFTR* | 4.07 | SET1 | 0.05 | 186 |
| KEGG | RNA_POLYMERASE | 28 | 2.68E-04 | 1 | *POLR2L* | 3.57 | SET2 | 0.05 | 186 |
| **FAME ALL Max IOP Rise** | | | | | | | | | |
| GO_MF | POLYOL_TRANSMEMBRANE_TRANSPORTER_ACTIVITY | 11 | 3.07E-06 | 3 | *SLC5A1,SLC2A13,AQP3* | 5.51 | SET1 | 0.05 | 1705 |
| GO_BP | NLRP3_INFLAMMASOME_COMPLEX_ASSEMBLY | 15 | 4.85E-06 | 3 | *GBP5,TLR4,NLRC3* | 5.31 | SET1 | 0.05 | 7472 |
| GO_CC | EUKARYOTIC_TRANSLATION_INITIATION_FACTOR_4F_COMPLEX | 13 | 2.37E-05 | 2 | *EIF4G3,EIF4G1* | 4.62 | SET1 | 0.05 | 989 |

*All the above gene-sets passed the Bonferroni-corrected threshold of 0.05/N, where N is the no. of genes within each set.

Abbreviations: IOP-intraocular pressure, FAME-Fluocinolone Acetonide in Diabetic Macular Edema, GO**-**Gene Ontology, BP-biological processes, MF-molecular function, CC-cellular components, KEGG- Kyoto Encyclopedia of Genes and Genomes, EUR-European

**Supplemental Table 16. Exome Data: Lambda values for gene burden and SKAT-O tests.**

| Gene Burden Analysis | Trait | Population | Method | Run 1 | Run 2 | Run 3 | Run 4 |
| --- | --- | --- | --- | --- | --- | --- | --- |
| **Discovery cohort: Fluocinolone Acetonide in Diabetic Macular Edema (FAME)** | | | | | | | |
| FAME Primary | Max. IOP Rise | ALL | Burden test | 0.95 | 0.98 | 0.99 | 1.01 |
| FAME Primary EUR | Max. IOP Rise | EUR | Burden test | 1.05 | 1 | 1.03 | 1.04 |
| FAME Primary | Max. IOP Rise | ALL | SKAT-O | 0.95 | 0.98 | 0.99 | 1.01 |
| FAME Primary EUR | Max. IOP Rise | EUR | SKAT-O | 1.05 | 1.00 | 1.03 | 1.04 |
| FAME Secondary | Responders | ALL | Burden test | 0.91 | 1.04 | 1.02 | 1.04 |
| FAME Secondary EUR | Responders | EUR | Burden test | 0.96 | 1.09 | 1.10 | 1.12 |
| FAME Secondary | Extreme Responders | ALL | Burden test | 0.37 | 0.7 | 0.94 | 0.59 |
| FAME Secondary EUR | Extreme Responders | EUR | Burden test | 0.34 | 0.68 | 1.02 | 0.58 |
| **Replication cohort: Mass Eye and Ear (MEE)/Retina Health Center (RHC)** | | | | | | | |
| MEE Primary | Max. IOP Rise | ALL | Burden test | 1.03 | 1.15 | 1.11 | 1.19 |
| MEE Primary EUR | Max. IOP Rise | EUR | Burden test | 0.95 | 0.97 | 1.01 | 0.96 |
| MEE Primary | Max. IOP Rise | ALL | SKAT-O | 1.03 | 1.15 | 1.11 | 1.19 |
| MEE Primary EUR | Max. IOP Rise | EUR | SKAT-O | 0.95 | 0.97 | 1.01 | 0.96 |
| MEE Secondary | Responders | ALL | Burden test | 1.00 | 1.03 | 1.06 | 1.01 |
| MEE Secondary EUR | Responders | EUR | Burden test | 1.01 | 1.08 | 1.11 | 1.08 |
| MEE Secondary | Extreme Responders | ALL | Burden test | 0.53 | 0.99 | 1.19 | 1.00 |
| MEE Secondary EUR | Extreme Responders | EUR | Burden test | 0.41 | 0.84 | 1.1 | 0.74 |
| **FAME-MEE/RHC Meta-Analysis** | | | | | | | |
| Primary | Max. IOP Rise | ALL | Burden test | 0.8 | 0.81 | 0.81 | 0.83 |
| Primary EUR | Max. IOP Rise | EUR | Burden test | 0.78 | 0.8 | 0.77 | 0.77 |
| Primary | Max. IOP Rise | ALL | SKAT-O | 0.8 | 0.81 | 0.81 | 0.83 |
| Primary EUR | Max. IOP Rise | EUR | SKAT-O | 0.78 | 0.8 | 0.78 | 0.77 |
| Secondary | Responders | ALL | Burden test | 0.83 | 0.81 | 0.78 | 0.8 |
| Secondary EUR | Responders | EUR | Burden test | 0.85 | 0.84 | 0.83 | 0.86 |
| Secondary | Extreme Responders | ALL | Burden test | 0.8 | 0.85 | 0.87 | 0.88 |
| Secondary EUR | Extreme Responders | EUR | Burden test | 0.81 | 0.88 | 0.87 | 0.84 |

Abbreviations: FAME-Fluocinolone Acetonide in Diabetic Macular Edema, IOP-intraocular pressure, MEE/RHC-Mass Eye and Ear/Retina Health Center

**Supplemental Table 17. Exome Data: List of all phenotypes tested in gene burden test and number of significant findings.** The table shows number of samples (N), number of genes passing Bonferroni cutoff (P ≤ 0.05/Number of genes tested) or Benjamini-Hochberg FDR cutoff (FDR<0.15) in Run1-3 and the percentage of genes passing nominal significance (P<0.05) in the negative control, Run4 (synonymous mutations) by their analysis, trait, population and method of gene burden test (Burden or SKATO). If there are more than 5% of genes passing significance in Run4 (see column “% of significant genes in Run4 (P<0.05)”), the top genes in Run1-3 from the corresponding analysis/trait are not further considered.

| **Gene burden analysis** | **Trait** | **Population** | **Method** | **N** | **Number (#) of significant genes (Bonferroni cutoff)** | **# of significant genes (Benjamini-Hochberg FDR<0.2)** | **% of significant genes in Run4 (P<0.05)** |
| --- | --- | --- | --- | --- | --- | --- | --- |
| **Discovery cohort: Fluocinolone Acetonide in Diabetic Macular Edema (FAME)** | | | | | | |  |
| Primary | Max Intraocular Pressure (IOP) Rise | All | Burden/SKAT-O | 532 | 0 | 2 (same gene)** | 4.8 |
| EUR Primary | Max IOP Rise | EUR | Burden/SKAT-O | 382 | 0 | 0 | 4.9 |
| Secondary | Responders | All | Burden | 532 | 0 | 0 | 4.9 |
| EUR Secondary* | Responders | EUR | Burden | 382 | 0 | 71 | 6 |
| Secondary* | Extreme Responders | All | Burden | 228 | 0 | 2522 | 7.3 |
| EUR Secondary* | Extreme Responders | EUR | Burden | 188 | 0 | 1151 | 7.4 |
| **Total # of significant genes considered:** | | | |  | 0 | **2** |  |
| **Replication cohort: Mass Eye and Ear (MEE)/Retina Health Center (RHC)** | | | | | | |  |
| Primary* | Max IOP Rise | All | Burden/SKAT-O | 575 | 2 | 50 | 7.4 |
| EUR Primary | Max IOP Rise | EUR | Burden/SKAT-O | 509 | 0 | 14 | 4.5 |
| Secondary | Responders | All | Burden | 586 | 0 | 0 | 4.7 |
| EUR Secondary | Responders | EUR | Burden | 520 | 1 | 0 | 5.8 |
| Secondary* | Extreme Responders | All | Burden | 297 | 0 | 426 | 8.3 |
| EUR Secondary * | Extreme Responders | EUR | Burden | 264 | 0 | 352 | 7.2 |
| **Total # of significant genes considered:** | | | |  | **1** | **14** |  |
| **FAME-MEE/RHC Meta-Analysis** | | | | | | |  |
| Primary | Max IOP Rise | All | Burden/SKAT-O | 1,093 | 0 | 2 | 4.54 |
| EUR Primary | Max IOP Rise | EUR | Burden/SKAT-O | 889 | 0 | 4 | 3.72 |
| Secondary | Responders | All | Burden | 1,104 | 0 | 0 | 3.71 |
| EUR Secondary | Responders | EUR | Burden | 889 | 0 | 1 | 4.34 |
| Secondary | Extreme Responders | All | Burden | 525 | 0 | 3 | 4.18 |
| EUR Secondary | Extreme Responders | EUR | Burden | 452 | 0 | 3 | 4.01 |
| **Total # of significant genes considered:** | | | |  | 0 | **13** |  |

 *Includes one or more significant genes in Run1-3 that had nominally significant P-value in synonymous run (Run 4), which was later deemed as false positive and excluded from consideration

**same gene was significant using standard burden test and SKAT-O.

Abbreviations: FAME-Fluocinolone Acetonide in Diabetic Macular Edema, IOP-intraocular pressure, MEE/RHC-Mass Eye and Ear/Retina Health Center

**Supplemental Table 18. Top genes from gene burden test that passed Bonferroni correction and/or Benjamini-Hochberg False discovery rate (FDR) below 0.15 for discovery cohort, Fluocinolone Acetonide in Diabetic Macular Edema (FAME) max intraocular pressure rise (IOP).** *MSTO1* passed both Bonferroni and FDR cutoff in both the gene burden analysis using Regular and SKAT-O test.

| Pop | Gene | Method | Lambda | Run | Mutations | Type of mutation  for Run1 and Run2 | N | Beta | Standard Error | Chi^2 | P-value | FDR (BH) | P synonymous | Bonferroni cutoff | Passed Bonferroni | # genotype (GT) case; # GT control | Individuals case; Individuals control | Total GT case; Total GT control |
| --- | --- | --- | --- | --- | --- | --- | --- | --- | --- | --- | --- | --- | --- | --- | --- | --- | --- | --- |
| **Discovery cohort FAME (Max. IOP Rise)** | | | | | | | | | | | | | | | | | | |
| ALL | *MSTO1* | SKAT-O** | 0.99 | Run 3 | missense;modifier | missense; intron variant, NMD transcript variant, non-coding transcript exon variant, non-coding transcript variant, regulatory region variant, splice donor region variant, splice polypyrimidine tract variant, splice region variant | 532 | 0.41 | 0.09 | 21.35 | 3.8E-06 | 0.11 | 0.5 | 1.8E-06 | No | 18(missense) + 79(modifier); 19(missense)+88(modifier) | 15(missense) + 53(modifier) ;16(missense) + 56 (modifier) | 6560(missense) + 25816 ( modifier);10464 (missense) + 41178 (modifier) |

P.synonymous: P.value from Run4 (negative control) with only synonymous mutations

\# GT control: No. of Genotypes with specific mutation in controls

\# GT case: No. of Genotypes with specific mutation in cases

Individuals cases: No. of individuals with specific mutation in cases

Individuals control: No. of individuals with specific mutation in controls

Total GT case: Total no. of non-missing genotypes in cases

Total GT control: Total no. of non-missing genotypes in control

*Genes that have significant P-value in synonymous run (Run 4), which suggests these gene might be false positives

** The same gene was significant using regular gene-burden test.
Abbreviations- FDR-false discovery rate, BH- Benjamini-Hochberg, FAME- Fluocinolone Acetonide in Diabetic Macular Edema, IOP-intraocular pressure GT-genotype

**Supplemental Table 19. Top genes from gene burden test that passed Bonferroni correction and/or Benjamini-Hochberg (BH) false discovery rate (FDR) for meta-analysis of Fluocinolone Acetonide in Diabetic Macular Edema (FAME) and Mass Eye and Ear/Retina Health Center (MEE/RHC).**

| Pop | Gene | Method | Lambda | Run | Mutations | Type of mutation  for Run1 and Run2 | N | Beta | Standard Error | P-value | FDR (BH) | Het  p-value | Direction | P synonymous | Bonferroni cutoff | Passed Bonferroni | # genotype (GT) case; # GT control | Individuals case; Individuals control | | Total GT case; Total GT control |
| --- | --- | --- | --- | --- | --- | --- | --- | --- | --- | --- | --- | --- | --- | --- | --- | --- | --- | --- | --- | --- |
| **FAME-MEE/RHC Meta-analysis (Max. IOP Rise)** | | | | | | | | | | | | | | | | | | | | |
| ALL | *LDHAP5* | Regular, SKAT-O | 0.81 | 3 | modifier | Non coding transcript variant | 1099 | -1.3 | 0.3 | 1.0E-06 | 0.03 | 0.6 | -- | NA | 1.82E-06 | Passed | 2;8 | 2;8 | | 1694;2696 |
| EUR | *ZNF248* | Regular, SKAT-O | 0.8 | 2 | missense | missense | 891 | -1.2 | 0.3 | 4.7E-06 | 0.08 | 0.6 | -- | 0.94 | 3.02E-06 | No | 1;10 | 1;10 | | 3038;4854 |
| EUR | *CLDN19* | Regular, SKAT-O | 0.77, 0.78 | 3 | missense; modifier | missense; 3_prime_UTR_variant,downstream_gene_variant,intron_variant,splice_polypyrimidine_tract_variant | 891 | -0.6 | 0.1 | 4.3E-06 | 0.12 | 1 | -- | NA | 1.84E-06 | No | 1(missense) + 13(modifier) ; 2(missense) + 40(modifier) | 1(missense) + 13(modifier) ; 2(missense) + 40(modifier) | |  |
| **FAME-MEE/RHC Meta-analysis (Responder)** | | | | | | | | | | | | | | | | | | | | |
| EUR | *PLCL2* | Regular | 0.84 | 2 | LoF2; missense | Stop lost; missense | 902 | 3.8 | 0.9 | 9.3E-06 | 0.15 | 0.3 | ++ | 0.98 | 3.02E-06 | No | 1(LoF2) + 7 (missense) ; 1(LoF2) + 0 (missense) | 1(LoF2) + 7 (missense) ; 1(LoF2) + 0 (missense) | | 2394;3780 |
| **FAME-MEE/RHC Meta-analysis (Extreme Responder)** | | | | | | | | | | | | | | | | | | | | |
| ALL | *RFC3* | Regular | 0.8 | 1 | LoF1 | Stop gained | 525 | 8.9 | 2.3 | 8.1E-05 | 0.10 | 0.5 | ++ | NA | 4.05E-05 | No | 2;0 | 2;0 | | 402;648 |
| ALL | *NMNAT1* | Regular | 0.85 | 2 | missense | missense | 525 | 11.2 | 2.5 | 6.1E-06 | 0.05 | 0.9 | ++ | NA | 3.25E-06 | No | 2;1 | 2;1 | | 632;1012 |
| ALL | *SPAG4* | Regular | 0.85 | 1 | LoF1; missense | Frameshift variant; missense | 525 | 5.2 | 1.1 | 2.3E-06 | 0.04 | 0.5 | ++ | 0.26 | 3.25E-06 | Passed | 1(LoF1) + 7 (missense) ; 0(LoF1) + 2 (missense) | 1(LoF1) + 7 (missense) ; 0(LoF1) + 2 (missense) | | 976;1580 |
| EUR | *MYO10* | Regular | 0.81 | 1 | LoF1 | Frameshift variant | 452 | 10.0 | 2.4 | 2.5E-05 | 0.02 | 0.9 | -+ | 0.38 | 5.30E-05 | Passed | 2;1 | 2;1 | | 530;902 |
| EUR | *ZFAND4* | Regular | 0.81 | 1 | LoF1 | Splice donor variant | 452 | 5.9 | 1.6 | 1.7E-04 | 0.08 | 0.6 | ++ | 0.41 | 5.30E-05 | No | 3;2 | 3;2 | | 336;568 |
| EUR | *SLC52A3* | Regular | 0.88 | 2 | missense | missense | 452 | 16.1 | 2.9 | 4.1E-08 | 5.7E-04 | 0.4 | ++ | 0.27 | 3.56E-06 | Passed | 3;0 | 3;0 | | 478;802 |
| P.synonymous: P.value from Run4 (negative control) with only synonymous mutations | | | | | | | | | | | | | | | | | | |  |  |
| # GT control: No. of Genotypes with specific mutation in controls | | | | | | | | | | | | | | | | | | |  |  |
| # GT case: No. of Genotypes with specific mutation in cases | | | | | | | | | | | | | | | | | | |  |  |
| Individuals cases: No. of individuals with specific mutation in cases | | | | | | | | | | | | | | | | | | |  |  |
| Individuals control: No. of individuals with specific mutation in controls | | | | | | | | | | | | | | | | | | |  |  |
| Total GT case: Total no. of non-missing genotypes in cases | | | | | | | | | | | | | | | | | | |  |  |
| Total GT control: Total no. of non-missing genotypes in control | | | | | | | | | | | | | | | | | | |  |  |
| Excluding genes from studies/trait with many genes (n>5) passing bonferroni or FDR cutoff in Run4 | | | | | | | | | | | | | | | | | | |  |  |

**Supplemental Table 20. Significant gene set or pathway burden associations for glucocorticoid-induced intraocular pressure rise that passed Bonferroni correction for discovery cohort Fluocinolone Acetonide in Diabetic Macular Edema (FAME)**

| Trait | Ancestry | Phenotype | Type of Mutation/Run | Collection | Gene-set/Pathway | P-value* | Benjamini-Hochberg P-value |
| --- | --- | --- | --- | --- | --- | --- | --- |
| FAME Primary | ALL | Max IOP Rise | LOF1 | C7:IMMUNESIGDB | GSE17186_NAIVE_VS_CD21LOW_TRANSITIONAL_BCELL_CORD_BLOOD_DN | 4.2E-06 | 0.02 |
| FAME Responders | ALL | Responders | LOF1 | C7:IMMUNESIGDB | GSE7548_NAIVE_VS_DAY7_PCC_IMMUNIZATION_CD4_TCELL_UP | 4.7E-06 | 0.02 |
| FAME Responders | ALL | Responders | LOF1 | C5:GO:BP | GOBP_ESTABLISHMENT_OR_MAINTENANCE_OF_CELL_POLARITY | 7.4E-05 | 0.03 |
| FAME Primary | EUR | Max IOP Rise | Run3 | C5:GO:BP | GOBP_SECRETION | 7.84E-05 | 0.03 |
| FAME Primary | ALL | Max IOP Rise | LOF1 | C2:CP:REACTOME | REACTOME_SURFACTANT_METABOLISM | 1.6E-04 | 0.01 |
| FAME Extreme Responders | EUR | Extreme Responders | LOF1 | C7:IMMUNESIGDB | GSE9988_LPS_VS_VEHICLE_TREATED_MONOCYTE_DN | 3.7E-06 | 1.03E-05 |
| FAME Extreme Responders | EUR | Extreme Responders | LOF1 | C7:IMMUNESIGDB | GSE17721_LPS_VS_PAM3CSK4_0.5H_BMDC_DN | 5.6E-06 | 1.03E-05 |
| FAME Extreme Responders | EUR | Extreme Responders | Run2 | C7:IMMUNESIGDB | GSE3920_IFNA_VS_IFNG_TREATED_FIBROBLAST_DN | 6.0E-06 | 1.0E-05 |
| FAME Extreme Responders | EUR | Extreme Responders | Run1 | C7:IMMUNESIGDB | GSE9988_ANTI_TREM1_VS_VEHICLE_TREATED_MONOCYTES_DN | 7.9E-06 | 1.03E-05 |
| FAME Extreme Responders | EUR | Extreme Responders | LOF1 | C2:CP:KEGG_LEGACY | KEGG_MELANOMA | 1.5E-05 | 0.00034 |
| FAME Extreme Responders | EUR | Extreme Responders | LOF1 | C5:GO:CC | GOCC_CHROMOCENTER | 1.7E-05 | 0.00046 |
| FAME Extreme Responders | EUR | Extreme Responders | Run3 | C5:GO:CC | GOCC_GLIAL_CELL_PROJECTION | 8.0E-05 | 4.2E-04 |
| FAME Extreme Responders | EUR | Extreme Responders | Run2 | C5:GO:MF | GOMF_MISFOLDED_PROTEIN_BINDING | 2.0E-04 | 2.7E-04 |
| FAME Extreme Responders | EUR | Extreme Responders | Run2 | C2:CP:REACTOME | REACTOME_REPRODUCTION | 2.0E-04 | 6.8E-04 |
| FAME Extreme Responders | EUR | Extreme Responders | Run2 | C2:CP:KEGG_LEGACY | KEGG_MAPK_SIGNALING_PATHWAY | 2.7E-04 | 3.3E-04 |

Abbreviations: FAME-Fluocinolone Acetonide in Diabetic Macular Edema, LOF1-Loss of function 1

* Gene sets that pass Bonferroni correction.

**Supplemental Table 21. Overlapping genes from genome-wide association study (GWAS) and whole exome sequencing analysis in discovery cohort.** Following table shows the overlapping genes between Gene Burden findings (P<0.05) and GWAS top findings (nearest genes to the genome-wide and subthreshold (P<5E0-7) variants from discovery GWAS and Meta-analysis and findings from Magma gene-based and Colocalization analysis)

| Gene | GWAS trait | GWAS Analysis | GWAS_SNP | GWAS P-value | Raw Beta (Effect size) | Raw Standard Error | Gene Burden Trait | Mutation | Gene Burden (P-value, FDR, Lambda) | # of Individuals (case ; control) | # of GT (case ; control) |
| --- | --- | --- | --- | --- | --- | --- | --- | --- | --- | --- | --- |
| *TMC5* | FAME Primary ALL | GWAS | chr16:19391115:C:T | 1.50E-07 | -4.3 | 0.9 | Meta-analysis Extreme Responder EUR | LoF1, missense, modifier | P:0.048 \| FDR: 0.99 \| Lambda: 0.87 | 32;46 | 36;56 |
| *TRPA1* | Meta-analysis Injections Primary ALL, Meta-analysis Primary ALL, FAME-MEE/RHC Replication Primary ALL | Metal, Replication | chr8:72086409:C:T | 1.80E-07,  4.7E-06 | -1.7 | 0.4 | FAME Primary EUR | missense | P:0.0054\| FDR: 0.71\| Lambda: 1.00 | 4;14 | 4;14 |
| *SPACA3* | Meta-analysis Primary Injections EUR, MEE/RHC Primary Injections EUR | Metal | chr17:33095158:C:T | 2.0E-07 | 1 | 0.21 | FAME Primary ALL | missense | P:0.0277\| FDR: 1.00\| Lambda: 0.98 | 2;9 | 2;9 |
| *PIEZO2* | Meta-analysis Primary ALL, MEE/RHC Primary ALL | GWAS, Metal | chr18:11313797:C:A, chr18:11287802:C:T | 4.1E-07, | -4.3 ,-1.4 | 0.9 | FAME Responder ALL | missense, modifier, moderate | P:0.0244\| FDR: 0.95\| Lambda: 1.02 | 113;156 | 213;292 |
| *PIEZO2* | Meta-analysis Primary ALL, MEE/RHC Primary ALL | GWAS, Metal | chr18:11313797:C:A, chr18:11287802:C:T | 4.1E-07,  4.7E-07 | -4.3 ,-1.4 | 0.9 | Meta-analysis Responder EUR | LoF1, missense | P:0.012 \| FDR: 0.99 \| Lambda: 0.84 | 39;34 | 41;35 |
| *GUCY2D* | FAME-MEE/RHC Replication Primary EUR | Replication | chr17:8838263:C:T | 4.89E-07 | -0.73 | 0.15 | Meta-analysis Responder EUR | missense | P:0.011 \| FDR: 0.99 \| Lambda: 0.84 | 18;11 | 18;11 |
| None passed Bonferroni cutoff=1.28E-03 [0.05/39] in Gene Burden (P<Bonferroni cutoff); | | | | | | | | | | | |

**Supplemental Table 22. Top genes in genome-wide association study (GWAS) previously associated with intraocular pressure (IOP) or with primary open angle glaucoma** (**POAG).** The table shows an overlap of the top candidates from discovery GWAS, FAME-MEE/RHC meta-analysis, and replication GWAS with candidate genes previously associated with IOP, POAG or related phenotypes. The top candidates from our study were selected using the nearest genes from genome wide as well as nominally significant GWAS loci, top candidates from MAGMA and colocalization analysis.

| **Trait** | **Gene** | **GWAS Variant (hg38)** | **P** | **Association/Function** | **Reference (PMID)** |
| --- | --- | --- | --- | --- | --- |
| **Target of Steroid (triamcinolone acetonide (TA) and dexamethasone (DEX)) treatment** | | | | | |
| FAME primary ALL | *HIPK2* | chr7:139570283:G:A | 4.2E-07 | This gene is differentially upregulated in human trabecular meshwork (TM) cells after treatment with low dose (0.1 mg/ml) TA and DEX and has roles in cell growth. | Fan et al., *Invest Ophthalmol Vis Sci*. 2008 (PMID: 18436822) |
| FAME primary ALL | *TMC5* | chr16:19391115:C:T | 1.5E-07 | This gene is differentially expressed in TM cells after long term DEX treatment. | Rozsa et al., *Mol Vis.* 2006 (PMID: 16541013) |
| **Associated with IOP/POAG GWAS** | | | | | |
| **Trait** | **Gene** | **GWAS Variant (hg38)** | **P** | **Association/Function** | **Reference (PMID)** |
| Meta-analysis primary ALL & Mass Eye and Ear/Retina Health Center (MEE/RHC) primary | *CCDC85A* | chr2:56602923:C:T | 1.3E-06,  1.4E-07 | The gene is the nearest gene to the loci associated with IOP, glaucoma progression, primary glaucoma in dogs, and radial glial maintenance. | MacGregor et al., *Nat Genet*. 2018 (PMID: 30054594), Khawaja et al., *Nat Genet.* 2018 (PMID: 29785010) |
| Meta-analysis primary ALL & MEE/RHC primary | *PIEZO2* | chr18:11287802:C:T,  chr18:11313797:C:A | 4.1E-07,  4.7E-07 | A variant within this gene was predicted to be associated with High-tension glaucoma. | Liu et al., *Scientific reports.* 2023 (PMID: 37741866) |
| Fluocinolone Acetonide in Diabetic Macular Edema (FAME) primary EUR | *SDK1* | chr7:4108958:C:G | 4.5E-07 | A variant within this gene was associated with primary open angle glaucoma (POAG) in multi-cohort GWAS in African ancestry | Verma et al., *Cell.* 2024 (PMID:38242088) |
| Meta-analysis primary ALL & FAME-MEE/RHC replication analysis | *TRPA1* | chr8:71978897:T:C | 4.69E-06 | Common and rare variants within this gene was associated with High-tension glaucoma. | Liu et al., *Scientific reports.* 2023 (PMID: 37741866) |
| MEE/RHC Primary ALL, Meta-analysis Primary EUR | *ACTR5* | chr20:38746749:T:C | 2.4E-06,  2.1E-06 | *ACTR5* eQTL in Esophagus Mucosa tissue colocalizes with loci associated with POAG MTAG Cross ancestry study. | Han et al., *Nat Genet* 2023( PMID: 37386247) |
| Meta-analysis Primary ALL | *FOXQ1* | chr6:1288153:C:T | 5.3E-06 | *FOXQ1* eQTL in Whole blood colocalizes with GWAS loci associated with POAG multi-trait analysis (MTAG) in European ancestry | Han et al., *Nat Genet* 2023(PMID: 37386247) |
| Meta-analysis Injections-only Primary ALL | *GAS7* | chr17:10267851:G:A | 5.9E-06 | *GAS7* eQTL in retina, fibroblast cell culture, and artery tibial tissues colocalizes with GWAS loci associated with IOP. This gene is also associated with IOP using gene-based analysis conducted with Magma. | Han et al., *Nat Genet* 2023(PMID: 37386247), Gharahkhani et al., *Nat Comm* 2021(PMID:33627673), Hamel et al., *Nat Comm* 2024(PMID:38195602) |
| FAME Responder ALL | *ZNF516* | chr18 :76502182:A:G | 7.5E-06 | *ZNF516* eQTL and splicing QTL in Adipose Subcutaneous, Thyroid, Nerve tibial and Skin Not Sun Exposed Suprapubic tissues colocalizes with IOP GWAS loci and also with loci from POAG MTAG cross-ancestry analysis. | Hamel et al., *Nat Comm* 2024(PMID:38195602), Han et al., *Nat Genet* 2023(PMID: 37386247) |
| Meta-analysis Primary EUR | *CALD1* | chr7:134812954:C:G | 8.9E-06 | *CALD1* eQTLin Breast Mammary tissue colocalizes with POAG cross-ancestry GWAS loci | Gharahkhani et al., *Nat Comm* 2021(PMID:33627673), Hamel et al., *Nat Comm* 2024(PMID:38195602), Han et al., *Nat Genet* 2023 (PMID: 37386247) |
| Mass Eye and Ear (MEE)/Retina Health Center (RHC) Primary ALL | *KCNK5* | chr6:39168761:C:T | 1.4E-09 | This gene is the nearest gene to the loci associated with intraocular pressure (IOP) in 2 studies, also associated with platelet crit, optic nerve head parameters, vertical cup-disc ratio, and glaucoma. | Han et al., *Nat Genet* 2023(PMID: 37386247), Han et al., *JAMA Ophthalmol* 2020(PMID:32352494), Ghoussaini et al., Nucleic Acids Research 2021(PMID:33045747) |
| MEE/RHC Primary ALL | *GLP1R* | chr6:39168761:C:T | 1.0E-08 | This gene passed Bonferroni significance in MAGMA gene-based analysis in this (GC-induced IOP) study. This gene was also associated with Glaucoma/POAG risk and modestly associated with Intra-ocular pressure (IOP). | Vasu et al., Ophthalmology 2025(PMID:39978437); Hallaj et al., *Am J Ophthalmol* 2025(PMID:39237049) |
| MEE/RHC injections Primary EUR | *ETV1* | chr7:13785499:C:T | 7.4E-08 | The gene is the nearest gene to the loci associated with IOP in Chinese population. | Huang et al., *Sci China Life Sci*. (PMID: 30591961) |

Abbreviations: IOP-intraocular pressure, FAME-Fluocinolone Acetonide in Diabetic Macular Edema, MEE/RHC: Mass Eye and Ear/Retina Health Center

**Supplemental Table 23. Top genes from gene burden test with previous intraocular pressure-associated evidence**

| N | Gene | Trait | Ancestry | Type of Mutation | Association | Reference (PMID) |
| --- | --- | --- | --- | --- | --- | --- |
| 1 | *MYO10* | FAME-MEE/RHC Meta-analysis Extreme Responder | EUR | Loss of function 1  (Frameshift variant) | Plays a role in IOP regulation through its function in trabecular meshwork | Sun et al., *Invest Ophthalmol Vis Sci.* 2019 (PMID: 30807639) |
| 2 | *GLP1R* | FAME-MEE/RHC Meta-analysis Responder | EUR | missense, modifier | Is associated with Glaucoma/POAG risk and modestly associated with Intra-ocular pressure (IOP). | Vasu et al., Ophthalmology 2025(PMID:39978437); Hallaj et al., *Am J Ophthalmol* 2025 (PMID:39237049) |

Abbreviations: FAME-Fluocinolone Acetonide in Diabetic Macular Edema, MEE/RHC: Mass Eye and Ear/Retina Health Center
