## Supplementary Materials for "Genome-Wide Association Study for Glucocorticoid-Induced Ocular Hypertension"

**Supplemental Materials**

**Abbreviations:**

IOP – intraocular pressure

MEE/RHC - Mass Eye and Ear/Retina Health Center

QC – quality control

GSA – gene set analysis

PCA – principal component analysis

LD – linkage disequilibrium

MAF – minor allele frequency

PC – principal component

WES – whole exome sequencing

BAM – Binary Alignment/Map

VCF – Variant Call Format

EUR – European

AFR – African

AMR- Admixed American

EAS – East Asian

SAS – South Asian

GQ – genotype quality

SNPs – single nucleotide polymorphisms

MAD – median absolute deviation

GTEx - Genotype-Tissue Expression

Non-PAR - non-pseudoautosomal region

Indels – insertion-deletion

HWE – Hardy-Weinberg Equilibrium

AB – allelic balance

GWAS – genome-wide association study

FAME - Fluocinolone Acetonide in Diabetic Macular Edema

GC – glucocorticoid

eQTL – expression quantitative trait loci

eQTL – splicing quantitative trait loci

CLPP - colocalization posterior probability

FDR – false discovery rate

OHTN – ocular hypertension

RINT – rank-based inverse normal transformation

**Methods**

**Whole exome sequencing**

For sequencing, an aliquot of genomic DNA (125ng in 50μL) is used as the input into DNA fragmentation (aka shearing). Shearing is performed acoustically using a Covaris focused-ultrasonicator, targeting 385bp fragments. Following fragmentation, additional size selection is performed using a SPRI cleanup. Library preparation is performed using a commercially available kit provided by KAPA Biosystems (KAPA Hyper Prep with Library Amplification Primer Mix, product KK8504) and with palindromic forked adapters using unique 8-base index sequences embedded within the adapter (purchased from IDT). The libraries are then amplified by 10 cycles of PCR. Enzymatic clean-ups are performed using Beckman Coultier AMPure XP beads with elution volumes reduced to 30μL to maximize library concentration. Following library construction, library quantification is performed using the Invitrogen Quant-It broad range dsDNA quantification assay kit (Thermo Scientific Catalog: Q33130) with a 1:200 PicoGreen dilution. Following quantification, each library is normalized to a concentration of 25 ng/μL, using a 10 mM Tris HCl pH 8.0 solution. All steps performed during the library construction process and library quantification process are performed on the Agilent Bravo liquid handling system.

After library construction, hybridization and capture are performed using the relevant components of IDT’s XGen hybridization and wash kit and following the manufacturer’s suggested protocol, with several exceptions. A single pre-hybridization pool is created. The pre-hybridization pool comprised of 96 unique libraries is created by equivolume pooling of the normalized libraries as well as 5 uL of Human Cot-1 and 2 ul of IDT XGen blocking oligos. The pre-hybridization pool undergoes lyophilization using the Biotage SPE-DRY. Post lyophilization, 4 uL of custom exome bait (TWIST Biosciences) along with 13 uL of hybridization mastermix is added to the lyophilized pool prior to resuspension. An initial incubation is performed at 95ºC for 30 seconds, after which time the incubation temperature is lowered to 65ºC at which it remains overnight. Library normalization and hybridization setup are performed on a Hamilton Starlet liquid handling platform, while target capture is performed on the Agilent Bravo automated platform.

After post-capture enrichment, library pools are quantified using qPCR (automated assay on the Agilent Bravo), using a kit purchased from KAPA Biosystems with probes specific to the ends of the adapters. Based on qPCR quantification, pools are normalized using a Hamilton Starlet and sequenced on one lane of a NovaSeq S4 flow cell using the NovaSeq XP workflow with a read length of 2x151bp.

**Comparison of maximal IOP change by ancestry**

To compare the distribution of maximal intraocular pressure (IOP) change between ancestries we performed an analysis of variance for maximal IOP change as a response variable and ancestry as a fixed effect using the aov() function in R. A post-hoc Tukey honest significant difference test in R was applied to the maximal IOP change between population pairs in the replication cohort, Mass Eye and Ear (MEE)/Retina Health Center (RHC), to be able to detect any significant difference by ancestry.

**Sample and Variant Quality Control**

Sample and variant quality control (QC) procedures were performed using PLINK 1.9 (<https://www.cog-genomics.org/plink/>) and PLINK 2.0 (<https://www.cog-genomics.org/plink/2.0/>) before performing both the common and rare variant association analyses.^1^

*Sample QC*

For the Global Screening Array (GSA) genotyping data, we applied the sample QC steps summarized in **Supplemental Table 1**. Individuals with missing genotypes (>2%), ancestry outliers, failing heterozygosity test (>5 standard deviation from the mean within each population), and one of each pair of related individuals (detected based on PI_HAT> 0.1875 using identity-by-descent test representing individuals between first and second degree relatives) were excluded from further analysis. For sex check, F Statistics were calculated based on heterozygosity rate on chromosome X and were compared to self-reported sex (F-stat < 0.2 for women and > 0.8 for men). Principal Component Analysis (PCA) was performed on a set of linkage disequilibrium (LD)-independent variants (r^2^<0.1 in a 200kb window and step size of 100 variants) with a call rate of >99% and minor allele frequency (MAF) > 0.05. This method uses the k-nearest neighbors algorithm applied to the top principal component (PCs) to predict the ancestral background using ancestry labels from the 1000 Genomes Project Phase 3 reference panel.^2^

For whole exome sequencing (WES) data, we applied the sample QC steps as summarized in **Supplemental Table 2**. We computed Binary Alignment/Map (BAM) and Variant Call Format (VCF)-based statistics to detect technical outliers within each ancestry group [European (EUR), African (AFR), Admixed American (AMR), East Asian (EAS), and South Asian (SAS)]. The samples that failed the BAM-level QC metrics, including mean target coverage (<20), percentage of chimeric reads (>5%), and percentage of contamination rate (>2%) were excluded from further analysis. VCF-based QC metrics evaluated included average genotype quality (GQ), call rate, number of single nucleotide polymorphisms (SNPs), number of singletons, number of insertions, number of deletions, insertion to deletion ratio, heterozygous to homozygous ratio, and transition to transversion ratio. All samples had genotype quality above 30 and call rate above 99%. Samples with four median absolute deviations (MAD) from the median from the heterozygous to homozygous ratio and transition to transversion ratio were excluded from further analysis.

Outliers above or below four MADs from the median from any of the above QC metrics (except for number of singletons) were manually inspected. Singletons eight MADs away from the median were considered as outliers. Samples that showed deviation for multiple QC metrics were excluded from further analysis. Samples whose self-reported sex and ancestry did not match their genotype-inferred sex and ancestry were excluded. One sample among a pair of related individuals (detected based on PI_HAT> 0.1875 using identity-by-descent test) were excluded from further analysis. All the above sample QC steps for WES data were performed using custom pipeline developed in R, Python, and Hail v0.2 (Hail 0.2.120-f00f916faf78 <https://github.com/hail-is/hail/commit/f00f916faf78>) for the Genotype-Tissue Expression (GTEx) Program.^3^

*Variant QC*

For GSA data, we applied the variant QC steps summarized in **Supplemental Table 1** on 729,900 hard genotype calls before imputation. Heterozygous calls in the non-pseudoautosomal region (non-PAR) of chromosome X in males that are supposed to be haploids were set to missing. Insertions or deletions (indels) above 50 base pairs, variants (SNPs and indels) with >2% missing genotypes, variants that failed Hardy-Weinberg Equilibrium (HWE) testing (P < 10^-6^) performed on variants with MAF>0.01, as well as variants that are multiallelic or strand ambiguous (G/C, A/T) were excluded from further analysis.

For WES data, we applied the variant QC steps as summarized in **Supplemental Table 3.** Multi-allelic sites were split into biallelic sites using Hail v0.2 (https://hail.is/). Variants with the following characteristics were excluded from the analysis: Variant Quality Score Recalibration below 99.8% for SNPs or 99.95% for indels, lying in Low Complexity Region, having excess heterozygosity (>54.69), monomorphic, having low genotype quality (GQ<20), having calls with allelic imbalance [allelic balance (AB)>0.8 or AB<0.2], mitochondrial variants, having heterozygous calls in non-PAR region of chromosome X in males, missingness rate >2%, and failing HWE test (P<10^-8^) performed separately in each ancestry group with more than 100 samples. Common variants (MAF>0.01) were excluded from further analysis. All the above variant QC steps for WES data were performed using a custom pipeline developed in R, Python and Hail v0.2 (Hail 0.2.120-f00f916faf78 <https://github.com/hail-is/hail/commit/f00f916faf78>) for GTEx.^3^

*Genotype imputation and post-imputation QC*

QCed hard call genotypes of 1,351 participants were imputed by minimac4-1.8.0 using TOPMed r3 (hg38) as reference panel through the TOPMed imputation server (<https://imputation.biodatacatalyst.nhlbi.nih.gov/>).^4^ Genotyping was available on 1,351 participants (greater than the 1,118 total participants included in the genome-wide association study [GWAS]) because some participants who were genotyped subsequently failed QC metrics – both phenotypic and genotypic. The reference panel contains phased whole genome sequences of ~158,000 individuals of diverse ancestry groups. Phasing was performed by eagle-2.4. The distribution of the imputation quality INFO score across three MAF groups (MAF < 0.01, MAF 0.01-0.05, and MAF > 0.05) is shown in **Supplemental Figure 1** and **Supplemental Table 4**. Post-imputation variant QC was applied separately for the discovery, Fluocinolone Acetonide in Diabetic Macular Edema (FAME) trial and replication, MEE/RHC cohorts as listed in **Supplemental Table 5**. Common variants (MAF>0.01) with the following characteristics were excluded: monomorphic variants, variants with low imputation quality (INFO score R^2^ < 0.6), indels above 50 base pairs, and those failing HWE test (P<10^-6^). HWE tests were performed within each subpopulation with >100 samples. Heterozygous calls in the non-PAR of chromosome X in males were set to missing. All post-imputation QC steps were conducted using pfile format (from PLINK 2.0) that maintained genotype dosages from the imputation. After filtering for MAF>0.01, a total of 9,448,884 variants in FAME and 9,572,205 variants in MEE/RHC were taken forward for the genome-wide association analysis. PCA was rerun on the post-imputed, QCed, LD-pruned SNPs (r^2^<0.1 in 200kb windows, step size 100kb, MAF>0.05, call rate >99%) and the top 5 PCs were included as a covariate in the GWAS analysis to control the effects of population substructures. All genomic chromosome positions are in genome build GRCh38.

**Colocalization analysis of expression and splicing QTLs with associated genomic loci**

To identify the candidate causal genes and underlying regulatory mechanisms that may be driving the genetic associations with glucocorticoid (GC)-induced IOP change in the significant GWAS loci, we conducted colocalization analysis using eCAVIAR.^5^ This tool assesses the probability that the same causal variant contributes to both the GWAS and a co-occurring expression or splicing quantitative trait locus (eQTL or sQTL) signal. We tested whether loci (both genome-wide significant and genome-wide suggestive loci) colocalized with eQTLs in human peripheral retina^6^ or eQTL and sQTLs in 49 human tissues from GTEx Program v8,^7^ using a pipeline developed in Hamel *et al*.^8^ Only e/sQTLs with at least five significant variants in the LD interval (r^2^>0.1 plus 50kb on either side of lead variant) around the lead GWAS variants were tested. We used ALL population [European, African, Asian, and Admixed American] in the 1000 Genomes Project Phase 3 for LD interval calculation and only used the LD proxy variant (r^2^>0.8) or the nearest variant in GTEx whole genome sequencing v8 data consisting of ALL population [European, African, Asian, and Admixed American] when the variant was absent in 1000 Genomes. All variants within the computed LD interval were used in the analysis. Colocalizing GWAS-e/sQTL-tissue signals with colocalization posterior probability (CLPP) above 0.01 and with e/sQTL false discovery rate (FDR) < 0.05 and/or P value <10^-4^, and GWAS P value < 0.05 were considered significant. LocusCompare plots comparing the GWAS -log(P values) to the e/sQTL-log(P values) of all variants in the LD window tested were generated with color coding of the points based on the variants’ LD relative to the lead GWAS variant or the variant with the highest CLPP. The plots were generated using custom code in Python that runs the tool: <https://github.com/boxiangliu/locucomparer>.

**Gene and gene-set level association analysis of GWAS using MAGMA**

To identify genes and pathways or gene ontologies associated with GC-induced ocular hypertension (OHTN), we conducted gene-based and gene-set (pathway)-based analyses of genetic associations using MAGMA^9^ by inputting summary statistics from GWAS analyses for all three phenotypes and populations. Genes were scored based on the most significant variant P value within 100kb around the gene start and end sites. Gene sets were downloaded from MSigDB (<http://www.qsea-misgb.org/gsea/msigdb/collections.jsp>) for the following databases: Gene Ontology with three domains: biological processes, molecular function, and cellular components; HALLMARK; Reactome; and Kyoto Encyclopedia of Genes and Genomes. Bonferroni correction at 0.05 significance level was used to determine significant genes and pathways.

**Gene and pathway burden testing using whole exome sequencing**

To identify genes and pathways with a higher frequency of rare variants in cases compared to controls and vice versa, we performed gene- and pathway-based burden tests for rare variants with different levels of predicted functional effects (**Supplemental Table 7**). The test was conducted for both discovery (FAME) and replication (MEE/RHC) cohorts. After sub setting the cohorts from the full sample set of post-QC WES data, for the remaining 532 FAME and 572 MEE/RHC samples (**Supplemental Figure 2**), additional QC steps were performed that included removal of monomorphic variants and variants with missingness rate >2%. Rare variants (MAF<0.01) were extracted after which functional annotation was performed using Ensembl’s Variant Effect Predictor.^10^ The number of rare variants used in the gene burden test for each outcome (Maximum IOP Change, Responder vs. Nonresponder, Extreme Responder vs. Extreme Nonresponder) and population (ALL or EUR) are listed in **Supplemental Table 8.** Standard gene burden test^11, 12^ was conducted for dichotomous traits (Responders vs. Nonresponders, Extreme Responders vs Extreme Nonresponders), whereas both burden and SKAT-O^13^ were conducted for the primary quantitative outcome (Maximum IOP Change) using REGENIE.^14^ RINT was applied to the quantitative trait, Maximum IOP Change, due to its non-normal distribution. Variants were grouped into four different categories based on their level of functional impact to the gene as defined in **Supplemental Table 7**:

Category 1: includes loss of function 1 variants

Category 2: loss of function 1, loss of function 2, and missense variants

Category 3: loss of function 1, loss of function 2, and missense, moderate, low, and modifiers

Category 4: synonymous variants

The burden test was run separately within each of these four categories. Category 4 was used as a negative control group. Genes in Categories 1-3 were considered significant if the P value passed Bonferroni cutoff (P <0.05/no. of genes tested) or if FDR was <0.15. In Category 4 any genes with P<0.05 were treated as false positives and were not considered as candidate genes. To avoid potential bias due to population sub-structure or small sample size we additionally excluded genes from Categories 1-3 when we detected substantial enrichment of genes passing P<0.05 than expected by chance in category 4. Pathway-burden test was performed by aggregating genes into pathways using the Molecular Signatures database. We used a custom pipeline written in Python to perform all the above analyses and to run the REGENIE tool.

**Gene burden testing in the FAME/RHC/MEE Meta-analysis of Whole Exome Sequencing using METAL**

We used the metafor package in R^15^ using restricted maximum likelihood method to perform meta-analysis between FAME and MEE/RHC cohorts for all 3 traits (Maximum IOP Change, Responders vs. Nonresponders, and Extreme Responders vs. Extreme Nonresponders) across 2 populations (ALL and EUR).

**Results**

**Ancestry Distributions of the GWAS Participants**

The percentage of variance explained by the top 20 genotype PCs is shown in **Supplemental Figure 5** for the discovery (FAME) and replication (MEE/RHC) samples. Most of the population structure variance was captured by the first five PCs and thus were included in the GWAS model as described in the Methods. The ancestral distribution of the FAME and MEE/RHC individuals based on top three PCs is shown in **Supplemental Figure 6**. There was consistent separation of the ancestral groups along these first three PCs. In the FAME cohort, 70% of the participants were of EUR ancestry, followed by 17% SAS, 7% AMR, and 6% AFR ancestry, and in the MEE/RHC cohorts, 86% were EUR, 8% AFR, and 7% AMR (**Table 1**).

**Discovery: Secondary Outcome GWAS (Common Variants)**

The Q-Q and Manhattan plots for the two secondary outcomes: dichotomous outcomes: responders vs. non-responders and extreme responder vs. extreme non-responders in the ALL ancestry analysis in discovery cohort are shown in **Supplemental Figures 7 and 8**, respectively. None of the variants passed genome-wide significance for the secondary outcomes.

**Replication: GWAS (Common Variants)**

The Manhattan plot for all the outcomes and ancestries for replication cohort, MEE-RHC, are shown in **Supplemental Figure 16**. We found three variants that passed genome-wide significance [chr6:39168761:C:T (rs9470989) (associated with IOP), chr12:71680955:G:T (rs10879312), chr17:71963822:T:C (rs7214582)] in the replication cohort for the primary outcome, but none were found for the secondary outcomes (Supplemental Table 12). chr6:39168761:C:T and chr12:71680955:G:T reached genome-wide significance (P = 1.44 X 10^-9^ and P = 2.8 X 10^-8^, respectively) in the ALL ancestry GWAS. chr17:71963822:T:C achieved genome-wide significance (P = 3.12 x 10^-8^) in the EUR GWAS using participants who only received intravitreal GC. The variant on chromosome 6 is closest to the transcription end site of *KCNK5* and the nearest gene based on the transcription start site is *SAYSD1*, the SAYSVFN motif containing 1 gene. The T allele is more common in the AFR population (MAF=0.12) and quite rare in the other populations; because of this, it is not possible to confidently estimate the effect on change in IOP for this variant in our current sample size. This variant was absent in discovery FAME cohort. In addition, none of the variants in LD (r^2^>0.1) with it (LD proxy variants) were significant and had opposite direction of effect in FAME GWAS. The variant on chromosome 12 is closest to the gene *TMEM19*, transmembrane protein 19 . The T allele is most common in the AFR (MAF=0.31) and AMR (MAR=0.19) populations. For each copy of the T allele that a participant had, they had a 2.75 mmHg *higher* rise in IOP, on average. The closest protein-coding gene to the variant on chromosome 17 is *SOX9*, a transcription regulator, although there is a long non-coding ribonucleic acid (RNA) (*ROCR*) closer to the variant. The C (risk) allele is common (allele frequency [AF] = 0.95) in the EUR population, and in other populations (AMR AF=0.97, AFR AF=0.99). For each copy of the C allele that a EUR participant had, there was a 3.6 mmHg greater rise in IOP after GC intravitreal injection. Both chromosome 12 and 17 variants were not significant in discovery FAME GWAS and had opposite direction of effects. The results of all LD-independent variant associations at P<5 X 10^-7^ for all outcomes and ancestries are shown in **Supplemental Table 12**.

**Colocalization Analysis for GWAS loci (Common Variants)**

In addition to the genome-wide significant and the replicated loci, we also performed colocalization analysis of 9 subthreshold (P < 5 X 10^-7^) locus from the discovery GWAS and meta-analysis. Of these five subthreshold loci significantly colocalized with expression and splicing (e/sQTL) (**Supplemental Tables 13-14).** Only one locus chr16:19391115:C:T) was associated with eQTLs in retina. This locus from discovery primary outcome GWAS in ALL ancestry colocalized with eQTLs targeting long non-coding genes, *CTA-363E6.2 (LOC105371114)* and *CTA-363E6.3* (**Supplemental Table 13**).

**Single-cell expression in anterior segment of the eye (Common Variants)**

Three other genes (*HIPK2, PIEZO2,* and *SDK1*) associated with subthreshold GWAS loci from discovery GWAS and meta-analysis demonstrated high expression across multiple tissues associated with IOP (**Supplemental Figure 19**) in the single cell RNA-seq dataset from the anterior segment of the eye.^16^

*SSFA2*, another candidate gene suggested by the colocalization analysis, exhibited high expression in vascular endothelial tissues. Similarly, the *PSD3* gene, linked with replicated locus chr8:19145108:T:C in the EUR ancestry, displayed markedly high expression in non-pigmented ciliary epithelium and fibroblasts from TM, CB, and iris **(Supplemental Figure 19)**.

**Ancestry Distributions of the Participants Included in the Whole Exome Sequencing Analyses**

The percentage of variance explained by the top 20 PCs applied to the WES data used in the gene burden test are shown in **Supplemental Figure 21**. The top 10 PCs capture the majority of the population variance. The ancestral distribution of the FAME and MEE/RHC individuals based on the first three PCs are shown in **Supplemental Figure 22**. There was consistent separation of the ancestral groups along these first three PCs, as found with the genotype array PCs.

**Whole Exome Sequencing Analysis: Gene and Gene Set Burden Testing (Rare Variants)**

In the meta-analysis of the FAME and MEE/RHC cohorts, there were a total of 10 significant genes (*LDHAP5, ZNF248, CLDN19, PLCL2, RFC3, NMNAT1, SPAG4, MYO10,* and *SLC52A3*) among which four (*LDHAP5, SPAG4, MYO10,* and *SLC52A3*) passed both the Bonferroni and Benjamini-Hochberg (BH) FDR cutoff (FDR<0.2) (**Supplemental Table 19)**. For the primary outcome meta-analysis of maximal IOP change, *LDHAP5* was significantly associated across ALL ancestry and two genes, *ZNF248* and *CLDN19,* in EUR ancestry. *PLCL2* showed association with the secondary outcome (responder vs. non-responder). For the Extreme Responder outcome 3 genes, *RFC3, NMNAT1, SPAG4*, showed significant association in ALL ancestry and 3 other genes, *MYO10, ZFAND4, SLC52A3* in the EUR ancestry.

Pathway burden analysis from the discovery cohort revealed immune-related signatures associated with CD4^+^ T cell and B cell activation, cell polarity, secretion, surfactant metabolism, glial cell morphology and MAPK signaling **(Supplemental Table 20)**.

Five nominally significant genes (P<0.05), *SPACA3, PIEZO2, TRPA1, TMC5,* and *GUCY2D*, from discovery rare variant analyses overlapped with GWAS findings (genome-wide and subthreshold, P<5 X 10^-7^) (**Supplemental Tables 21**). **Supplemental Figure 24** shows the single cell expression of top WES genes in tissues from the anterior segment of the eye.

**Overlap with genes previously associated with POAG and IOP**

We found three top variants- *KCNK5, GLP1R,* and *ETV1-* from replication (MEE/RHC) GWAS residing in or near by genes that were previously associated with POAG or IOP (Supplemental Tables 22)One of the variants, which was genome-wide significant (P *=* 1.4 X 10^-9^) is nearby *KCNK5*, a gene which also had a variant, rs2815114, associated (P = 6.43 X 10^-11^) with POAG in a multi-trait GWAS meta-analysis^17^. The associated variant from our GC-induced OHTN GWAS, rs9470989, is not in high LD with the POAG-associated variant (r^2^=0.01). *GLP1R* passing Bonferroni significance in gene level test (P = 1 X 10^-8^) and nominally significant in the WES meta-analysis in EUR responders (P=0.02, lambda=0.83) has been previously associated with POAG risk (**Supplemental Table 23**). Agonists of GLP-1R, glucagon like peptide 1 receptor that stimulates glucose-induced insulin secretion has been associated with reduced ocular hypertension and POAG risk^18^ and unclear IOP reduction^19^
